# Circulating cardiovascular proteomic associations with genetics and disease

**DOI:** 10.1101/2024.10.18.24315790

**Authors:** Kathryn A. McGurk, Lara Curran, Arunashis Sau, Fu Siong Ng, Brian Halliday, James S. Ware, Declan P. O’Regan

## Abstract

**Background:** The analysis of the circulating proteome can identify translational modifiers and biomarkers of disease expressivity and severity at a given time point. Here we explore the relationships between protein measures implicated in cardiovascular disease and whether they mediate causal relationships between cardiovascular risk factors and disease development.

**Methods:** To understand the relationships between circulating biomarkers and genetic variants, medications, anthropometric traits, lifestyle factors, imaging-derived measures, and diagnoses of cardiovascular disease, we analysed measures of nine plasma proteins with *a priori* roles in genetic and structural cardiovascular disease or treatment pathways (ACE2, ACTA2, ACTN4, BAG3, BNP, CDKN1A, NOTCH1, NT-proBNP, and TNNI3) from the Pharma Proteomics Project of the UK Biobank cohort (over 45,000 participants sampled at recruitment).

**Results:** We identified significant variability in circulating proteins with age, sex, ancestry, alcohol intake, smoking, and medication intake. Phenome-wide association studies highlighted the range of cardiovascular clinical features with relationships to protein levels. Genome-wide genetic association studies identified variants near *GCKR*, *APOE*, and *SERPINA1*, that modified multiple circulating protein levels (BAG3, CDKN1A, and/or NOTCH1). NT-proBNP and BNP levels associated with variants in *BAG3*. ACE2 levels were increased with a diagnosis of hypertension or diabetes and were influenced by variants in genes associated with diabetes (*HNF1A, HNF4A*). Two-sample Mendelian randomisation identified ACE2 as protective for systolic blood pressure and Type-2 diabetes.

**Conclusions:** From a panel of circulating proteins, the results from this observational study provide evidence that ACE2 is causally associated with hypertension and diabetes. This suggests that ACE2 stimulation may provide additional protection from these cardiovascular diseases. This study provides an improved understanding of the circulating pathways depicting cardiovascular disease dynamics.

**Graphical abstract:** 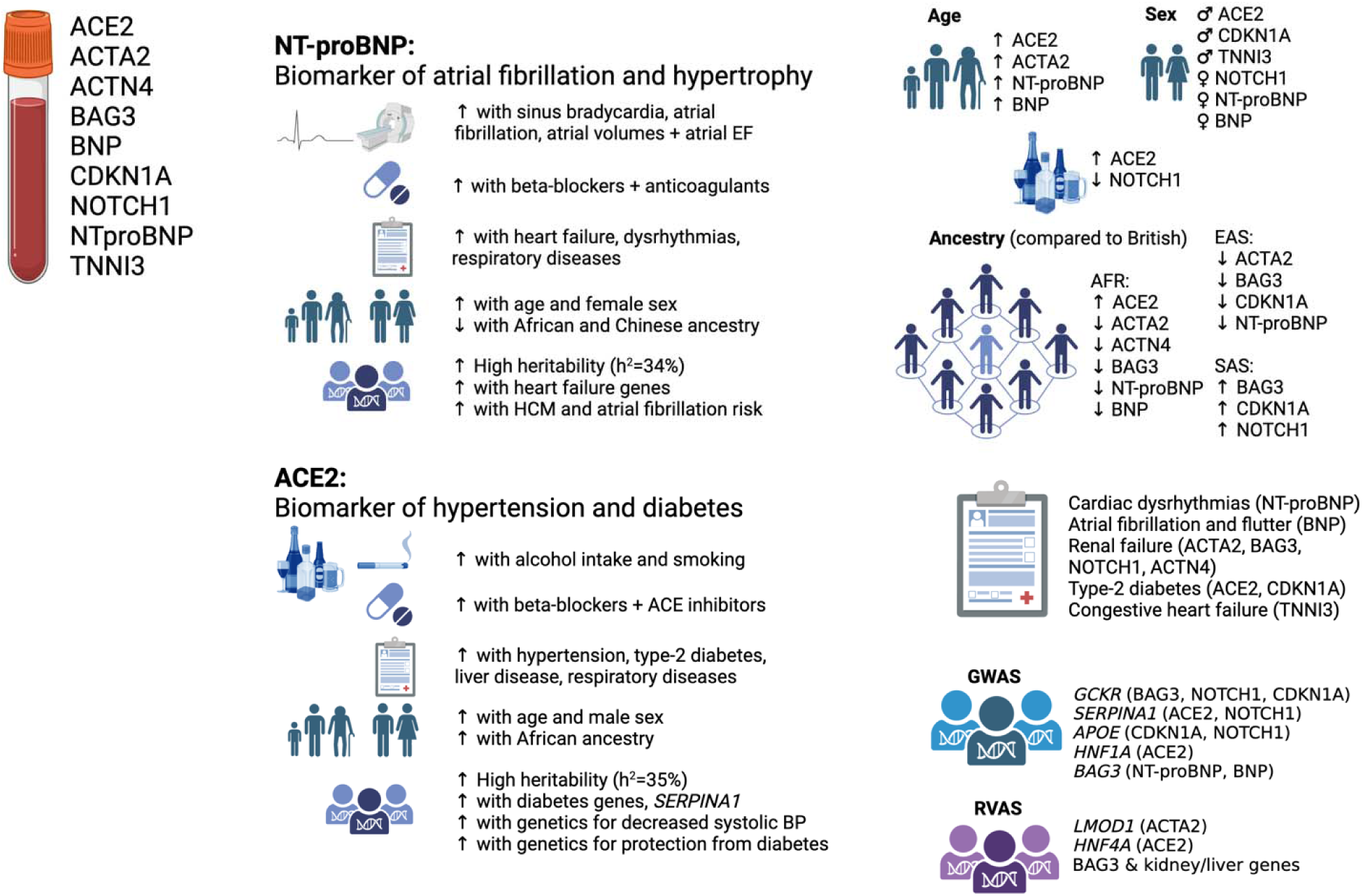

## Introduction

Circulating proteomics provides information on the landscape of biological function, metabolism, disease, and homeostasis. While genetic testing can provide a once-off invariable assessment from birth, proteome analyses can identify translational modifiers and biomarkers of disease expressivity and severity at a particular sampling time point. In addition to large-scale proteome-wide discovery and risk score analyses, in-depth assessments of selected proteins based on *a priori* implication in disease are required to fully identify and interpret the relationships between protein measures of interest and cardiovascular disease, and whether they mediate causal relationships between cardiovascular risk factors and disease development.

To understand these relationships, we undertook an in-depth assessment of the circulating levels of nine plasma proteins implicated through genetic studies for roles in structural cardiovascular disease or treatment pathways (aortopathies, cardiomyopathies, congenital heart disease, and heart failure^1^): ACE2 (*ACE2*; angiotensin-converting enzyme 2), ACTA2 (*ACTA2*; actin alpha 2, smooth muscle), ACTN4 (*ACTN4;* Actinin Alpha 4), BAG3 (*BAG3*; BAG cochaperone 3), BNP (*NPPB;* brain natriuretic peptide or B-type natriuretic peptide), CDKN1A (*CDKN1A;* cyclin-dependent kinase inhibitor 1A), NOTCH1 (*NOTCH1; Neurogenic locus notch homolog protein 1*), NT-proBNP (*NPPB*; N-terminal prohormone of brain natriuretic peptide), and TNNI3 (*TNNI3*; Troponin I).

ACE2, with functionally opposing roles of ACE1 (which is unmeasured here, is the target of ACE inhibitors, and is involved in the biosynthesis of the angiotensin II vasoconstrictor), is a part of the renin-angiotensin-aldosterone system that regulates blood pressure by catalysing the hydrolysis of vasoconstrictor angiotensin II to other vasodilator angiotensins. ACTA2 is an actin protein involved in the contraction of smooth muscle and genetic variants in *ACTA2* cause autosomal dominant familial thoracic aortic aneurysm and aortic dissection^2^. ACTN4 is an actin-binding protein that anchors the myofibrillar actin filaments in muscle cells. BAG3 is involved in chaperone-assisted selective autophagy of damaged cytoskeletal components and variants in the *BAG3* gene cause autosomal dominant dilated cardiomyopathy and myofibrillar myopathy^1,3^. The genetic locus of CDKN1A has been recently identified in case-control GWAS analyses of cardiomyopathies^4,5^ and it is a regulator of cardiomyocyte cell cycle arrest^6^. NOTCH1 controls cell fate decisions and variants at the locus have been previously associated with congenital heart disease^7^ and trabeculation^8^. PreproBNP and proBNP (both unmeasured here) are cleaved by a convertase to create BNP and NT-proBNP (inactive with a longer half-life than BNP); both are biomarkers and upregulated with heart failure and myocardial stretching. PreproBNP is encoded by *NPPB* which has unique expression in the heart, highest in the atrial appendage (GTEx)^9^. BNP has roles in natriuresis, diuresis and vasodilatation. TNNI3 is a mediator of relaxation in the sarcomeric thin filament of cardiac striated muscle and is exclusively expressed in the heart. Variants in *TNNI3* cause autosomal dominant hypertrophic cardiomyopathy^1,10^.

Exploration of the relationships between circulating proteins and upstream factors that may influence their levels (e.g., anthropometric traits and protein quantitative trait loci identifiable from genome-wide common and rare genetic factors) as well as downstream clinical endpoints they may predict or prevent (cardiovascular diagnoses, clinical features, and ECG and MRI-derived traits), would aid our understanding of these biomarkers of cardiovascular disease. Here, we use the results of the Pharma Proteomics Project of the UK Biobank cohort: Olink proteomic data from plasma samples collected at recruitment. We analysed the proteomics from over 45,000 participants to identify relationships with the full spectrum of available UK Biobank data and tested for causal relationships (**Graphical abstract**).

## Methods

### Study overview

The UK Biobank (UKB) cohort study recruited 500,000 participants aged 40 to 69 years old from across the United Kingdom between 2006 and 2010 (National Research Ethics Service, 11/NW/0382)^11^. This study was conducted under the terms of access of projects 47602 and 40616. All participants provided written informed consent.

Genotyping array data and exome sequencing data were available for over 450,000 participants. Sub-studies analysed baseline plasma proteomics in over 45,000 participants^12^ and recalled participants for electrocardiograms (ECG)^13^ and cardiac magnetic resonance (CMR) imaging^14,15^. Additional phenotypic and outcome data included hospital episode statistics, self-reported questionnaire data, alcohol intake^16^, and smoking status. Protein levels were assessed for association with genetic, phenotypic, and clinical outcome data (**Figure 1**). Two-sample Mendelian randomization was used to determine causality and risk prediction of Cox proportional hazards regression models were created to understand whether the biomarkers predicted incident diagnoses.

**Figure 1.**
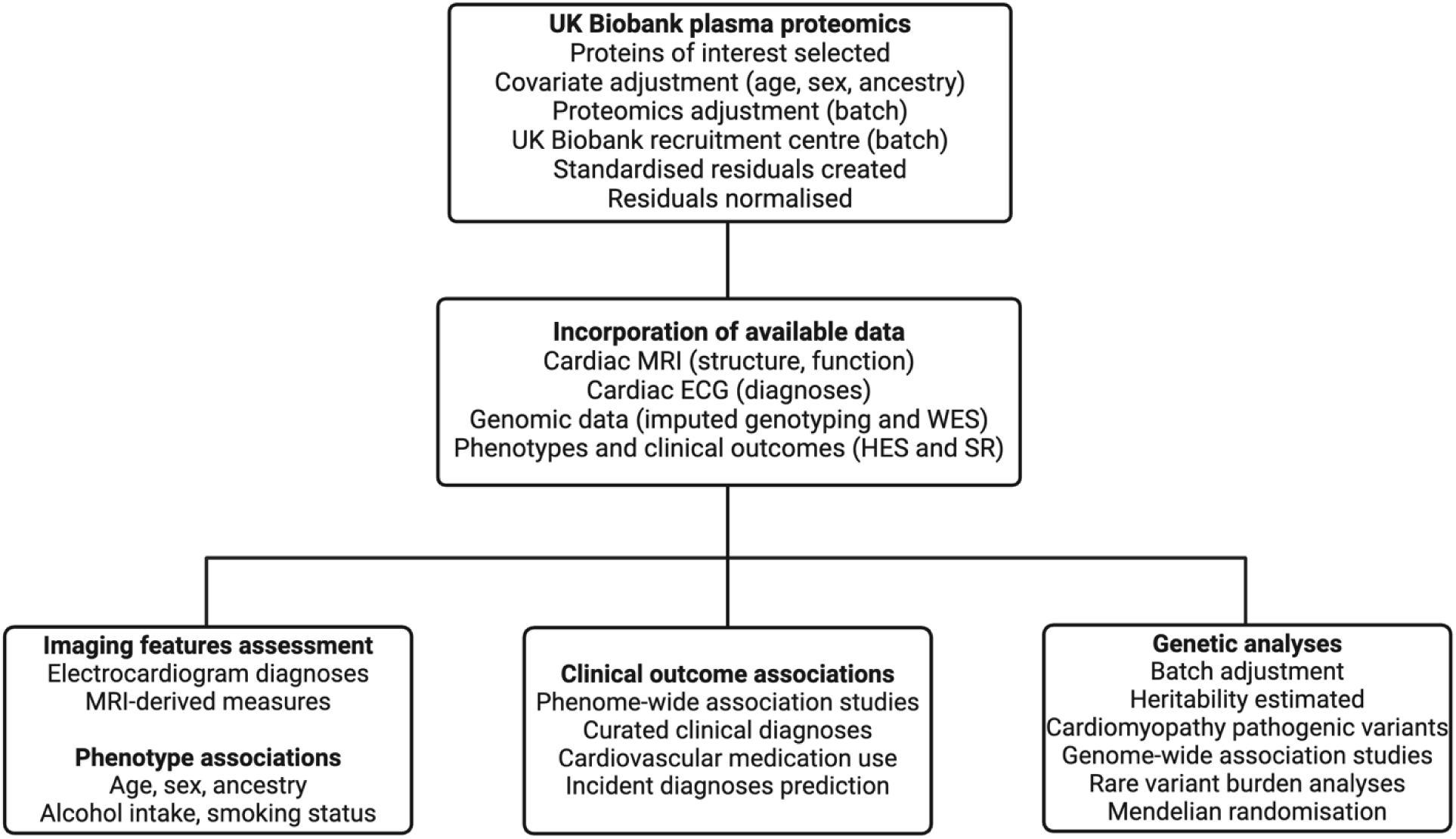
Study flow chart and protein summary. A summary of the main analysis steps and data available for the analysis of nine plasma proteins and the genetic and outcome associations. MRI, magnetic resonance imaging; CMR, cardiac MRI; WES, whole exome sequencing; HES, hospital episode statistics; SR, self-reported.

### Proteomics

Plasma from the initial UKB assessment visit (2006-2010) was collected. Details of participant randomisation, sample handling, Olink proteomics assay through the antibody-based Olink Explore 3072 PEA, data processing, and quality control are as detailed previously^12^. Briefly, proteomics was undertaken on samples of a randomly selected subset of 46,595 UKB participants at the baseline visit, of which 46,011 participants had complete measures of the nine proteins of interest with *a priori* implication in structural cardiovascular disease or treatment pathway through genetic studies (aortopathies, cardiomyopathies, congenital heart disease, and heart failure; ACE2, ACTA2, ACTN4, BAG3, BNP, CDKN1A, NOTCH1, NT-proBNP, and TNNI3).

Covariates were assessed (age at recruitment, UKB recruitment centre, genetically determined sex, and proteomics batch). The protein levels are provided in normalized protein eXpression (NPX); Olink’s arbitrary unit in log_2_ scale.

### Genetic analyses

Genotyping array data and exome sequencing data were available for over 450,000 participants. Genotype calling and exome sequencing were performed and imputed as described previously^11,17^. For genome-wide association studies (GWAS), the imputed UKB genotyping data was used, where a minor allele frequency of >0.001 in autosomes was included. Individuals with more than 5% missing genotypes and SNPs with more than 5% missingness were excluded. Participant sex discrepancies, heterozygosity, and relatedness were handled by keeping only genetically European individuals and participants included in the UKB principal components analysis^11^. SNPs deviating from Hardy-Weinberg equilibrium (1x10^-8^) and those with an imputation INFO score of <0.4 were excluded. Individuals with proteomics data were extracted and GWAS was undertaken using GCTA software (version 64)^18^. A sparse genetic relationship matrix (GRM) was created and FastGWA was undertaken with a mixed linear model, adjusting for the genotyping array batch. Genes of independent loci were prioritized through LocusZoom and eQTLs from GTEx (v8). Phenotype associations through GWAS catalog and PheWEB, were assessed. Heritability was estimated by creating a genetic relationship matrix in GCTA and using a restricted maximum likelihood analysis (REML) to estimate the variance explained by the SNPs that were used to estimate the GRM. Mendelian randomization was undertaken using GWAS summary statistics from published literature^4,5,19,20^ with NT-proBNP, BNP, and ACE2, GWAS results using the R package TwoSampleMR. Exposure variants were included if GWAS significant (P<5x10^-8^). Tests of pleiotropy, Steiger directionality, and heterogeneity were assessed.

Rare variant association studies (RVAS) were undertaken using Regenie software on the DNA Nexus Research Analysis Platform^21^. The genotyping data for step 1 of Regenie included SNPs in autosomes with a minor allele frequency <0.01, missingness of >0.01, a minor allele count of <20, deviations from Hardy-Weinberg equilibrium (5x10^-15^), and individuals with greater than 10% missingness, were excluded from the analysis. Interchromosome, SNPs in linkage disequilibrium (indep-pairwise 1000 100 0.9), and areas of low complexity, were excluded for step 1. Exome sequencing data for step 2 was quality controlled for variants in the autosomes with missingness less than 10%, variants where less than 90% of all genotypes for that variant had a read depth less than 10, deviations from Hardy-Weinberg equilibrium (1x10^-15^), and individuals with more than 10% missingness. Step 2 of Regenie was run over different allele frequencies (singletons, 0.01, 0.001) for 6 overlapping, protein-altering variant, custom masks (LoF only; missense only (flagged by >1 of 5 deleterious software); missense only (all); missense only (flagged by all of 5 deleterious software); protein-altering variants (LoF and missense flagged by >1 of 5 deleterious software); protein-altering variants (all Lof and missense)), where the minimum minor allele count was at least 3. Bonferroni significance for 18,117 included genes was P<2.76x10^-6^.

Cardiomyopathy-associated rare variants were identified as previously published^5,22,23^ for HCM and DCM. Individuals were classified as genotype negative (SARC-NEG) if they had no rare protein-altering genetic variation (minor allele frequency <0.001 in the UKB and the Genome Aggregation Database) in any genes that may cause or mimic HCM or DCM. These genes represented an inclusive list of genes with definitive or strong evidence of an association with cardiomyopathy, moderate evidence, and genes associated with syndromic phenotypes^1,3,10^. This SARC-NEG group was compared with individuals with disease-associated rare variants in genes with strong or definitive evidence for HCM (*MYBPC3, MYH7, MYL2, MYL3, TNNI3, TNNT2, TPM1,* and *ACTC1*) and DCM (*BAG3, DES, DSP, FLNC, LMNA, MYH7, PLN, RBM20, SCN5A, TNNC1, TNNT2,* and *TTN*). Analysis was restricted to robustly disease-associated variant classes for each gene^3,10^ and to variants sufficiently rare to cause penetrant disease (filtering allele frequency <0.00004 for HCM and 0.000084 for DCM^24^). Variants were classified as pathogenic/likely pathogenic (SARC-P/LP) if reported as P/LP for cardiomyopathy in ClinVar and confirmed by manual review.

### Cardiac MRI and ECG analyses

A sub-study recalled participants for imaging, including CMR^25^, and ECG. For CMR, volumetric traits were measured using quality-controlled deep learning algorithms^14^. Deep neural networks were used for short-axis cine segmentation via a fully convolutional network to label pixels containing myocardium. The performance of image annotation using this algorithm is equivalent to a consensus of expert human readers and achieves subpixel accuracy for cardiac segmentation^14^. 5,324 participants with proteomics had imaging data available.

The ECGs were performed according to a defined protocol and analysed using proprietary software (GE CardioSoft, Boston, MA). Data from the first imaging visit (instance 2, n=42,386) was labeled using a previously trained convolutional neural network designed to identify six diagnoses from the ECG^13^: sinus bradycardia, sinus tachycardia, left bundle branch block, right bundle branch block, 1st degree AV block, and atrial fibrillation. Automated diagnoses had F1 scores above 80% and specificity over 99%^13^. The ECGs were preprocessed with a bandpass filter 0.5 to 100hz, a notch filter at 60hz, and re-sampling to 400hz. Zero padding resulted in a signal with 4,096 samples for each lead for a 10s recording, which was used as input to the neural network model. The binary outputs (presence or absence of each diagnosis) were used for subsequent analyses. 4,831 participants with proteomics had ECG data available.

For analyses comparing plasma proteomics sampled at recruitment to records from the subsequent imaging appointment, the analyses were duplicated to include an adjustment for the difference in time.

### Statistical analyses

The analyses were undertaken using R (v4.1.2) and the UKB research analysis platform. The results are expressed as mean and standard deviation (SD). The Student’s t-test was used to assess differences in means for quantitative traits and Fisher’s exact test for counts. Pearson’s correlation coefficient described relationships. Effect sizes are presented as standardised beta coefficients. The proteomic measures were adjusted using multiple linear regression for age at recruitment, age^2^, UKB recruitment centre, genetically determined sex, age x sex interaction, proteomics batch, and genetically determined European ancestry, and the resulting standardized residuals (mean=0,

SD=1) were normalized by an inverse rank normalization. Cardiovascular-associated medication intake was self-reported and curated for participants who reported taking medications of interest (**Table S6**). Phenome-wide association studies (PheWAS) were undertaken using the PheWAS R package with clinical outcomes and coded phenotypes converted to 1,840 categorical PheCodes. P-values were deemed significant with Bonferroni adjustment for the number of PheCodes measured.

Cox proportional hazards regression models were assessed with the full cohort of participants for NT-proBNP, BNP, and ACE2, and created using the first reported UKB data (summarising the first date of a report from all UKB data (Hospital episode statistics, primary care, self-reported, etc.)) for heart failure, cardiomyopathy, atrial fibrillation, hypertension, diabetes, and myocardial infarction, as identified through PheWAS analysis, using the survival and survminer R packages by age to death, diagnosis, or last date of follow up report. Participants diagnosed before recruitment were excluded. Participants who died without a diagnosis were also excluded as a sensitivity analysis **(Figure S15**).

## Results

### Circulating biomarkers of age, sex, alcohol intake, smoking status, and ancestry

46,011 participants had measures of the proteins assessed (participant characteristics: **Table S8**). The correlation between the levels of NT-proBNP and BNP was R=0.67 (**Figure S9**). Levels of ACTA2 had the strongest relationships with the other circulating protein levels (e.g., ACTN4 (R=0.30), NT-proBNP (R=0.30), BNP (R=0.21), BAG3 (R=0.21); **Figure S9**).

A positive relationship was identified with age at recruitment for measures of ACTA2 (R=0.42), NT-proBNP (R=0.34), BNP (R=0.23), and ACE2 (R=0.16; **Figure S1**). ACE2, CDKN1A, and TNNI3, were increased in male compared to female participants (β=0.47, P=1.0x10^-16^; β=0.20, P=6.61x10^-68^; β=0.21, P=3.97x10^-127^; respectively; **Figure S2**).

NOTCH1, NT-proBNP, and BNP were increased in female participants (β=0.06, P=8.61x10^-258^; β=0.42, P=1.62x10^-278^; β=0.25, P=3.50x10^-69^; respectively; **Figure S2**).

A positive relationship was observed between alcohol intake and smoking status (smoker at recruitment compared to never smoked), and measures of ACE2 (R=0.14 and β=0.22, P=1.70x10^-44^; respectively) and NOTCH1 (R=-0.15 and β=-0.16, P=6.11x10^-23^). BAG3, CDKN1A, and TNNI3 also had relationships with smoking status (β=-0.08, P=7.94x10^-8^; β=0.10, P= 2.38x10^-10^; β=-0.05, P=0.0008; respectively).

Proteomic variability was observed with ancestry. Participants of self-reported African or Caribbean ancestry (n=584 and n=434, respectively) had increased average ACE2 and decreased ACTA2, ACTN4, BAG3, NT-proBNP, and BNP, compared to British ancestry which dominates the UKB cohort (n=40,228, 87% British; **Figure 2**). Participants of Chinese ancestry (n=131) had decreased average ACTA2, BAG3, CDKN1A, and NT-proBNP, and participants of Indian ancestry (n=495) had increased average BAG3, CDKN1A, and NOTCH1, compared to British ancestry (**Figure 2**).

**Figure 2.**
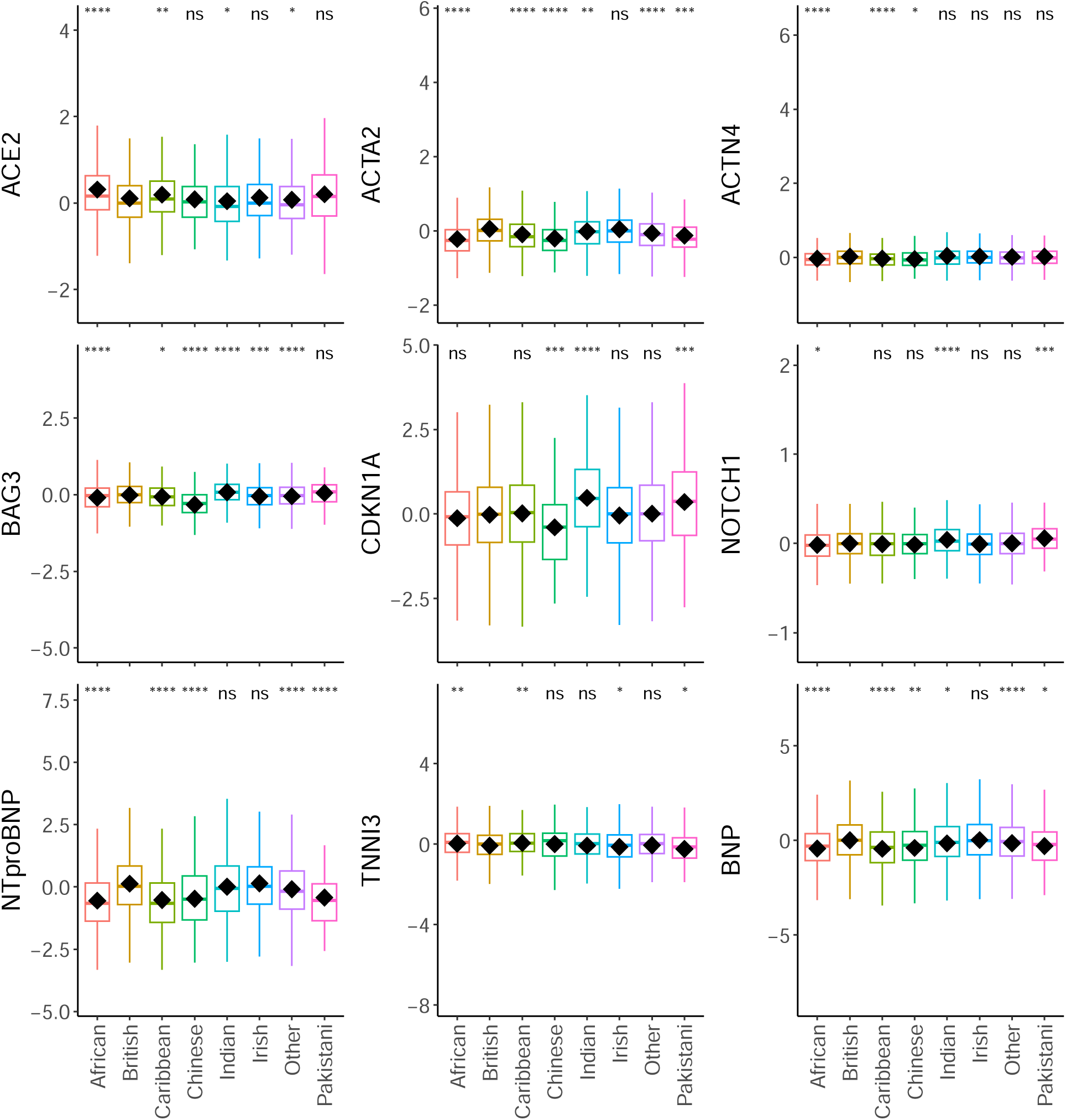
The relationships with proteins and ancestry. The plots depict the significant differences in protein levels across self-reported ancestries. Participants of self-reported African or Caribbean ancestry had increased average ACE2 and decreased ACTA2, ACTN4, BAG3, NT-proBNP, and BNP, compared to British ancestry. Participants with Chinese ancestry had decreased average ACTA2, BAG3, CDKN1A, and NT-proBNP, and participants with Indian ancestry had increased average BAG3, CDKN1A, and NOTCH1, compared to British ancestry. The significance of differences in means as derived by Student’s t-test are denoted as stars compared to British ancestry. The y-axis units are Olink’s arbitrary unit in log_2_ scale. The sample sizes were as follows (African n=584, British n=40228, Caribbean n=434, Chinese n=131, Indian n=495, Irish n=1200, Other n=2736, Pakistani n=143).

### Medication use

Medication use was reported at recruitment when the blood was sampled for proteomics (n=46,011 participants) and 8.4 years later (range 3.8-12.7 years) at the imaging appointment (n=5,324). 39%-75% of the individuals with reported cardiovascular medications at recruitment also reported the medication at the imaging appointment (39% on antiplatelets, 57% on beta blockers, 60% on angiotensin receptor blockers (ARBs), 65% on ACE inhibitors, 69% on calcium channel blockers, and 75% on anticoagulants).

Most of the protein levels (measured from blood samples collected at recruitment) had significant associations with medication use reported at recruitment (**Table S6**). To understand whether the protein levels may herald a disease that will require future treatment, protein measures at recruitment were assessed for an association with medication reported at the imaging visit. Participants with increased ACE2, NT-proBNP, and BNP levels at recruitment positively associated with beta-blocker use at the imaging visit 8 years later. ACE2 was also associated with ACE inhibitor (β=0.27, P=7.90x10^-11^) and ARB use (β=0.22, P=8.44x10^-4^) and participants with increased NT-proBNP or BNP at recruitment were more likely to report anticoagulant use (β=1.02, P=5.44x10^-6^; β=0.91, P=6.07x10^-6^; respectively) at the imaging visit (**Table S6**). The results were similar when the proteins were adjusted for the time between recruitment and the imaging visit (**Table S6**).

To further stratify these associations and understand whether the increase in plasma proteins at recruitment was due to medication use at recruitment or predictive of future medication use, we assessed the change in medication use: whether the protein levels at recruitment were significantly altered with medication i) intake reported only at recruitment, ii) uptake by the imaging visit, or iii) longer-term use and reported during both visits, compared to participants without reported medication (**Figure 3**). ACE2 levels predicted the future uptake of beta-blockers and ACE inhibitors, suggesting that ACE2 levels herald prescription and associated diseases. NT-proBNP and BNP levels predicted long-term use and future uptake of anticoagulants (**Figure 3**). The increase in NT-proBNP and BNP levels observed for beta-blockers may be in part due to the use of the respective medications and/or overt disease at recruitment, as they predicted both current and future use. Adjustment for time since imaging had little effect on the results (**Figure S13**).

**Figure 3.**
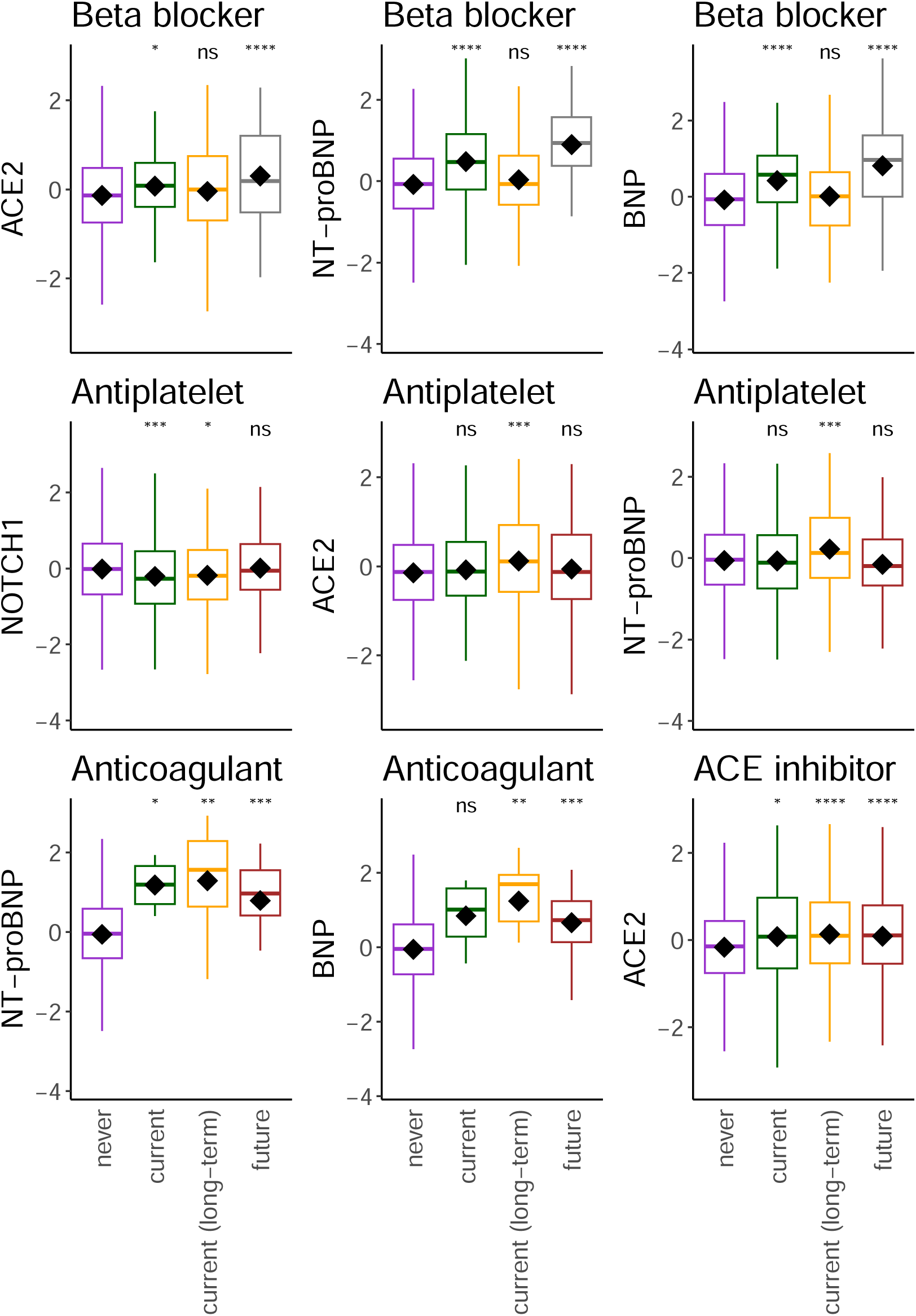
The relationships with proteins and medication intake. The plots depict the proteins measured at recruitment that were significantly increased with medication intake reported only at recruitment (current), or the imaging visit on average 8 years later (future), or reported during both visits (current (long-term)). The significance of differences in means as derived by Student’s t-test are denoted as stars compared to no report of the medication (never). The y-axis units are standardized residuals after adjustment for covariates. The data only includes those with proteomics who attended the imaging visit (n=5,324).

### Association with clinical features and diagnoses

Through phenome-wide association studies (PheWAS), the most significant associations with each protein were with cardiac dysrhythmias (NT-proBNP), atrial fibrillation and flutter (BNP), chronic renal failure (ACTA2, BAG3, NOTCH1, ACTN4), type-2 diabetes (ACE2, CDKN1A), and congestive heart failure (TNNI3; **Figure 4, Table S1, Figures S16-S30**). Renal diseases associated with most of the protein measures analysed (**Figure S25, Table S1**). ACE2, ACTA2, NT-proBNP, and BNP levels associated with a range of respiratory conditions (**Figure S17, Table S1**). ACE2 levels associated with type-2 diabetes, epilepsy, tobacco use disorder, alcohol-related disorders, and liver diseases (**Figures S16-S30, Table S1**).

**Figure 4.**
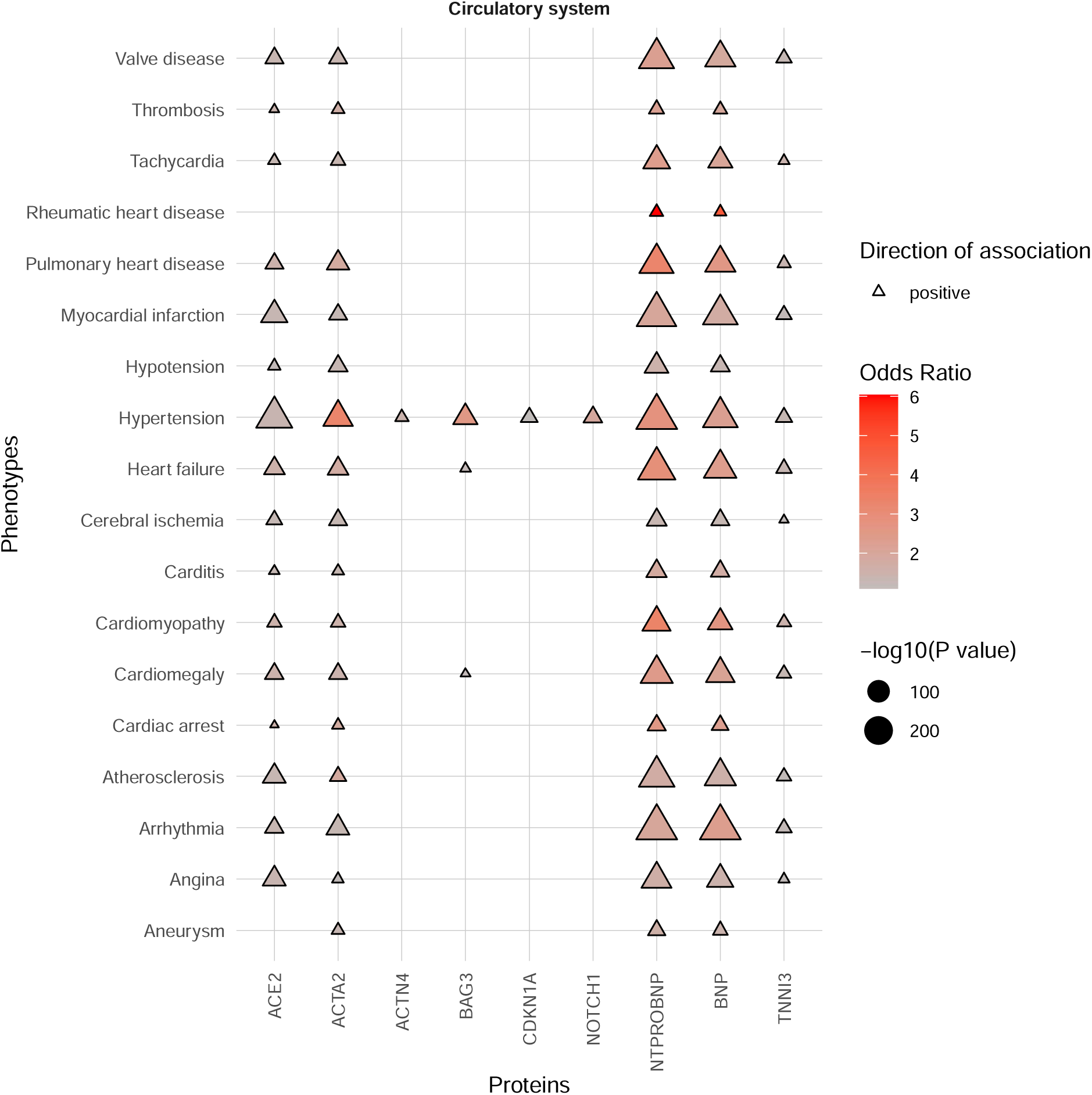
Phenome-wide association study results of the plasma protein levels with selected circulatory disorders. Phenotypes as phecodes are described on the y-axis and the protein traits on the x-axis. Each point denotes a significant PheWAS association with a Bonferroni correction for the number of analyzed phecodes. The shape and colour denote the direction of effect and odds ratio. Only the most significant associations with selected, non-redundant phenotypes of the circulatory disorder category are presented for clarity. See **Table S1** for the full PheWAS results.

A curated analysis was used to assess associations between the nine circulating proteins and specific diagnoses of cardiomyopathies, muscular dystrophy, heart failure, scoliosis, respiratory failure, coronary disease, cardiac arrhythmia (including atrial fibrillation and flutter), stroke, hypertension, valve disease, hypercholesterolemia, and diabetes (**Figure S14, Table S2**). ACE2, ACTA2, TNNI3, NT-proBNP, and BNP levels, had the most associations with curated traits. Hypertension and heart failure correlated with all proteins, except NOTCH1 and CDKN1A, respectively.

### Cardiac ECG-AI diagnoses and MRI parameters

Participants with increased NT-proBNP at recruitment were diagnosed more frequently with sinus bradycardia (β=0.25, P=2.82x10^-7^) and atrial fibrillation (β=0.92, P=7.24x10^-9^) on future ECG-AI analysis. The associations remained significant when individuals with reported beta-blocker use were removed from the analysis (sinus bradycardia, β=0.25, P=9.44x10^-7^; atrial fibrillation, β=0.83, P=0.0001). The association with atrial fibrillation was also observed for BNP (β=0.81, P=2.27x10^-6^). Adjustment for the time between recruitment and imaging had little effect (NT-proBNP and sinus bradycardia, β=0.27, P=2.64x10^-7^; NT-proBNP and atrial fibrillation, β=1.17, P=2.29x10^-10^; BNP and atrial fibrillation, β=0.96, P=1.15x10^-7^).

Analyses of CMR-derived traits identified relationships between NT-proBNP and BNP and increased left atrial volume (R=0.14-0.17) and decreased atrial ejection fraction (R=-0.14-**-**0.19). NT-proBNP also correlated with decreased ventricular wall thickness (R=-0.21) and volumes (R=-0.11-**-**0.18) and increased ventricular ejection fraction (R=0.12). The relationships between NT-proBNP and BNP with atrial volumes (R=0.11-0.21) and with atrial ejection fractions (R=-0.11-**-**0.17) were not affected by adjustment for the time between the recruitment and imaging visit (**Table S7**).

### Carriers of HCM and DCM pathogenic variants

Participants carrying a pathogenic/likely pathogenic variant in a HCM-associated gene, had increased levels of ACE2 and NT-proBNP (but not BNP) at recruitment (β=0.47, P=0.0004; β=0.62, P=0.0007; respectively). Participants carrying a pathogenic/likely pathogenic variant in a DCM-associated gene also had increased NT-proBNP at recruitment (β=0.32, P=0.0002). These associations were likely due to overt cardiomyopathy, as the signals became non-significant with the removal of individuals with diagnosed cardiomyopathy.

### Genetic association studies

SNP-based heritability of the protein levels, the amount of variation in the protein levels estimated to be due to genetic factors, identified ACE2 levels with the highest estimated heritability (34.5%), followed by NT-proBNP (33.5%), BAG3 (25.7%), NOTCH1 (22.3%), BNP (18.9%), ACTA2 (17.0%), and CDKN1A (14.2%). ACTN4 and TNNI3 levels had very low and non-significant heritability estimates; either more influenced by non-genetic factors or measurement error.

To identify genetic modifiers of the circulating protein levels, we undertook a genome-wide association study (GWAS; **Figure 5, Table S3**) and a rare variant association study (RVAS) for each protein measure. The GWAS and/or RVAS of the levels of ACTA2, BAG3, CDKN1A, NOTCH1, NT-proBNP, and BNP, identified the expected gene-protein pair (cis-expression quantitative trait loci were identified at the genetic loci of *ACTA2, BAG3, CDKN1A, NOTCH1,* and *NPPB*).

**Figure 5.**
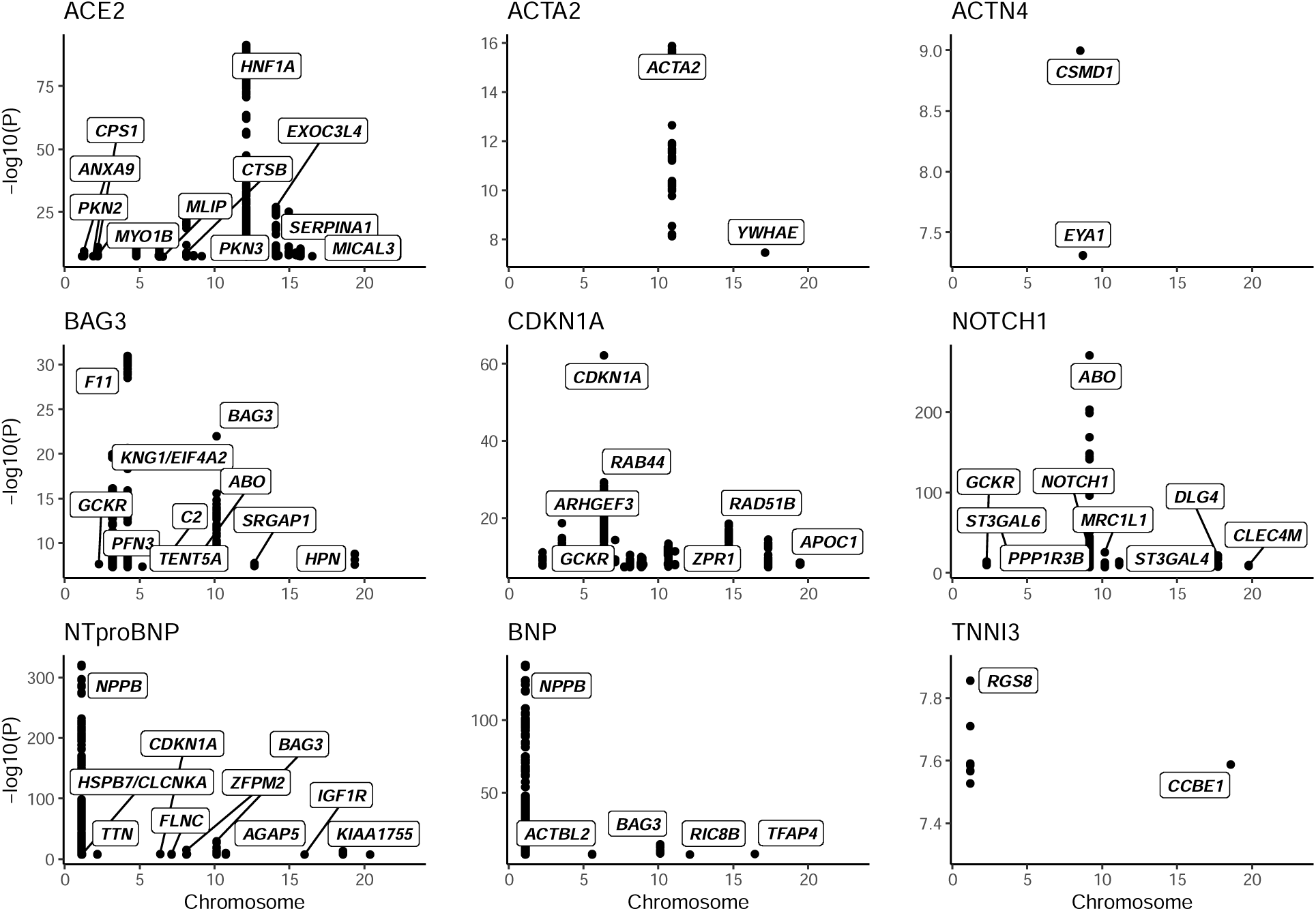
Significant genome-wide association study results. The Manhattan plots present the GWAS significant SNPs for the nine protein levels. The prioritized gene is noted for the significant loci identified. A subset of gene labels for ACE2, CDKN1A, and NT-proBNP, have been selected to allow for presentation. The y-axis is cut at a minimum of 5e-08 (7.3). Please see **Table S3** for the full GWAS results.

Recurrent modifiers included *GCKR* for BAG3, CDKN1A, and NOTCH1 levels, *APOE* for CDKN1A and NOTCH1 levels, and *SERPINA1* for ACE2 and NOTCH1 levels (**Table S3**). Glucokinase regulator (*GCKR*) is a regulatory protein that inactivates glucokinase in liver and pancreatic islet cells and has been previously associated with hyperlipidemia and diabetes. Apolipoprotein E (*APOE*) is an apolipoprotein involved in lipoprotein metabolism. Serpin family A member 1 (*SERPINA1*) is a serine protease inhibitor associated with alpha 1-antitrypsin deficiency. The gene is linked to chronic obstructive pulmonary disease, emphysema, and chronic liver disease.

Rare variant association studies (RVAS; **Table S4**) found that ACTA2 levels associated with variants in *LMOD1*. Leiomodin 1 (*LMOD1*) has been implicated in smooth muscle dysfunction and thoracic aortic aneurysm and dissection previously and is predicted to interact with ACTA2^26^.

### ACE2 and its potential role in hypertension and diabetes

Increasing ACE2 levels at recruitment was correlated with systolic blood pressure (R=0.21; **Figure S14**)) and was predictive of an incident hypertension diagnosis (**Figure 7, Figure S15**). Two-sample Mendelian randomisation with a genetic instrument for systolic blood pressure^19^ showed evidence for decreased systolic blood pressure to increase circulating ACE2 (**Figure 6, Figure S3-S4, Table S5**). This suggests a protective role for increased ACE2 levels in blood pressure control.

**Figure 6.**
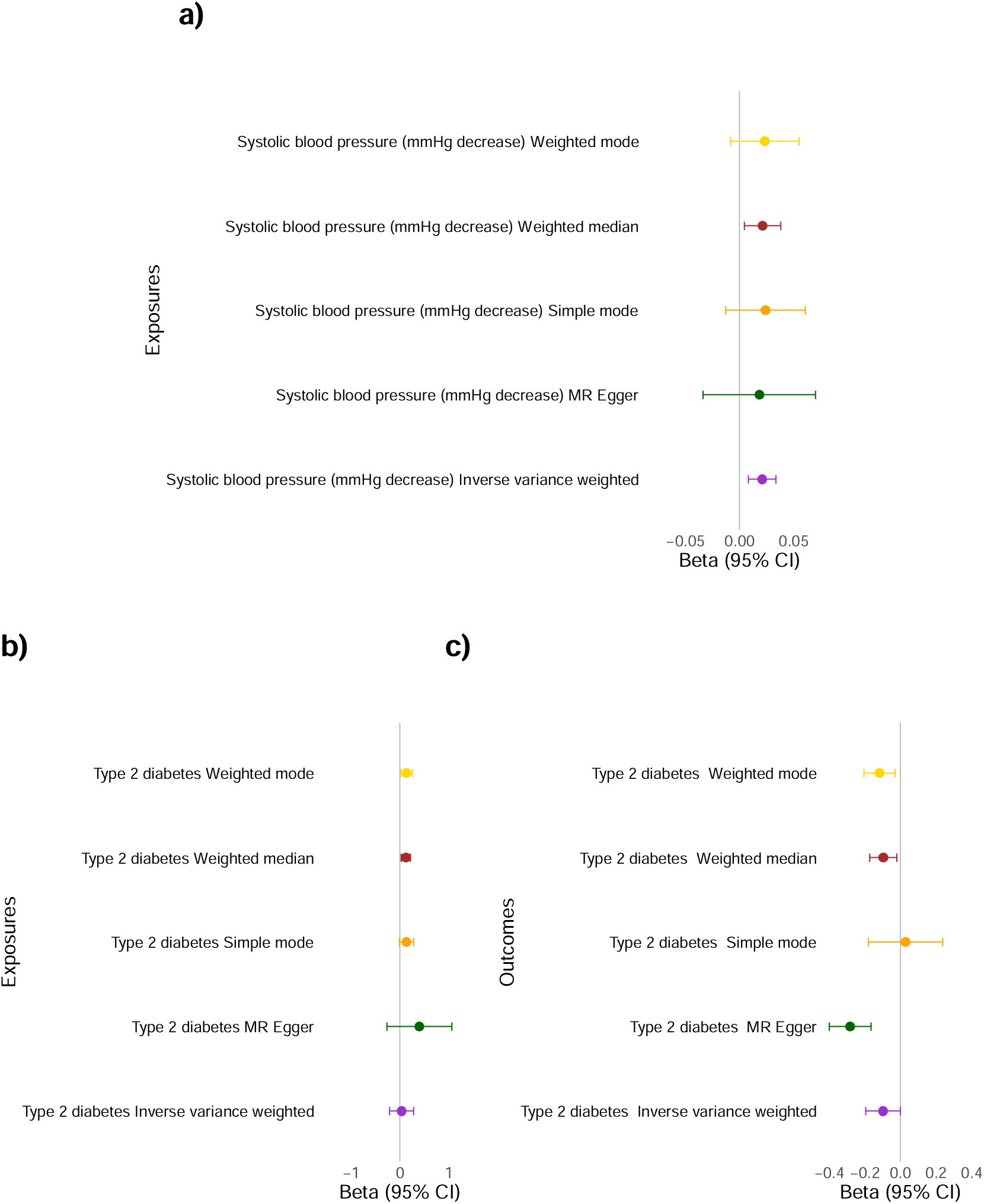
Evidence of a causal relationship between systolic blood pressure, type-**2 diabetes, and ACE2.** Increased circulating ACE2 can decrease blood pressure through the creation of vasodilators and is causally associated with type-2 diabetes. A genetic predisposition for decreased systolic blood pressure is associated with increased ACE2. **a)** Mendelian randomization genetic determination model of systolic blood pressure (mmHg decrease) genetic instruments as exposures for ACE2 outcome. Two-sample Mendelian randomization was undertaken with ACE2 (using the GWAS results) and decreased systolic blood pressure (from GWAS summary statistics of published data). **b)** Mendelian randomization genetic determination model of type-2 diabetes genetic instruments as exposures for ACE2 outcome. Two-sample Mendelian randomization was undertaken with ACE2 (using the GWAS results) and type-2 diabetes (from GWAS summary statistics of published data). **c)** Mendelian randomization genetic determination model of ACE2 genetic instrument as an exposure for type-2 diabetes. See **Table S5** and **Figures S3-S4** for further details.

**Figure 7.**
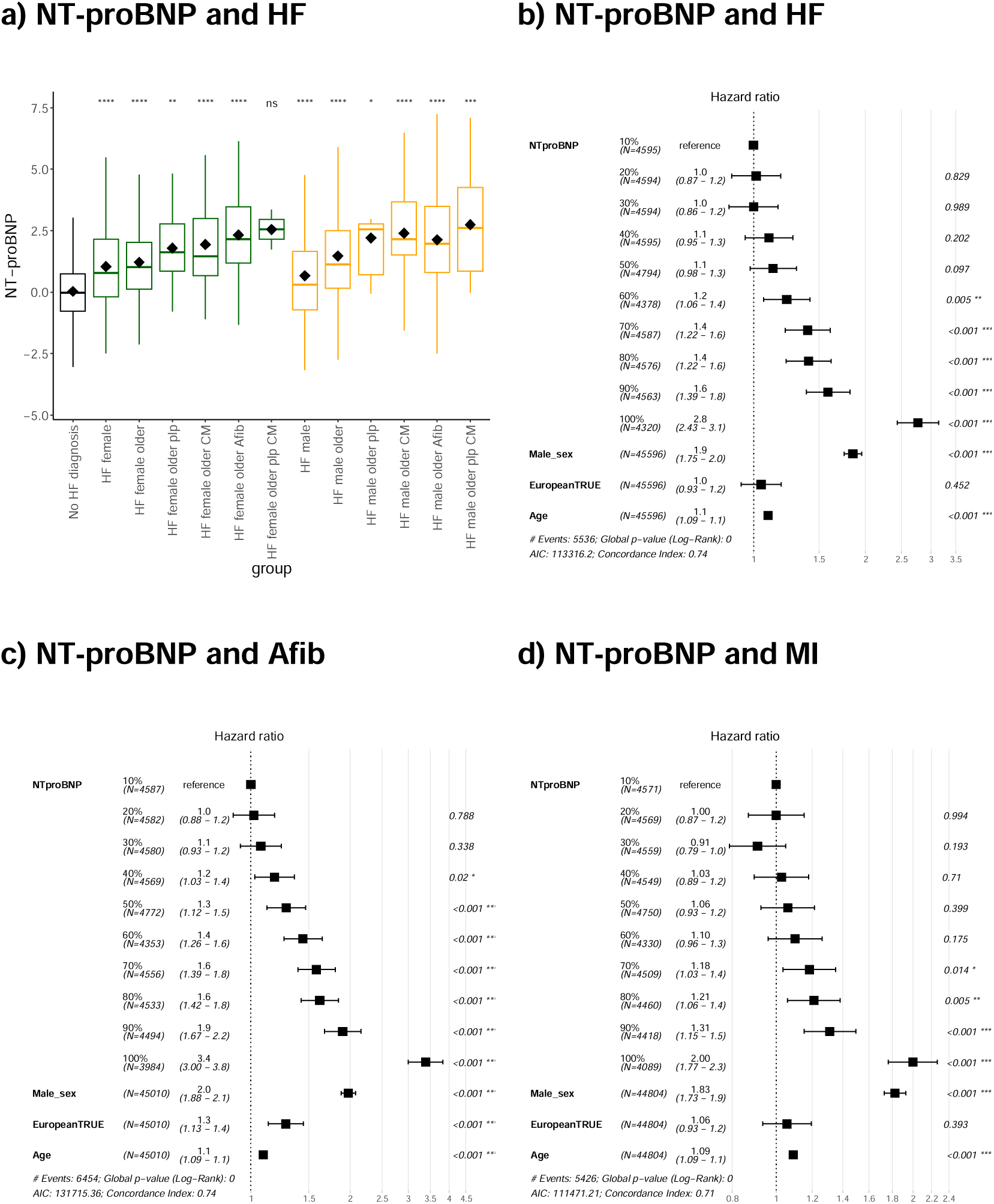
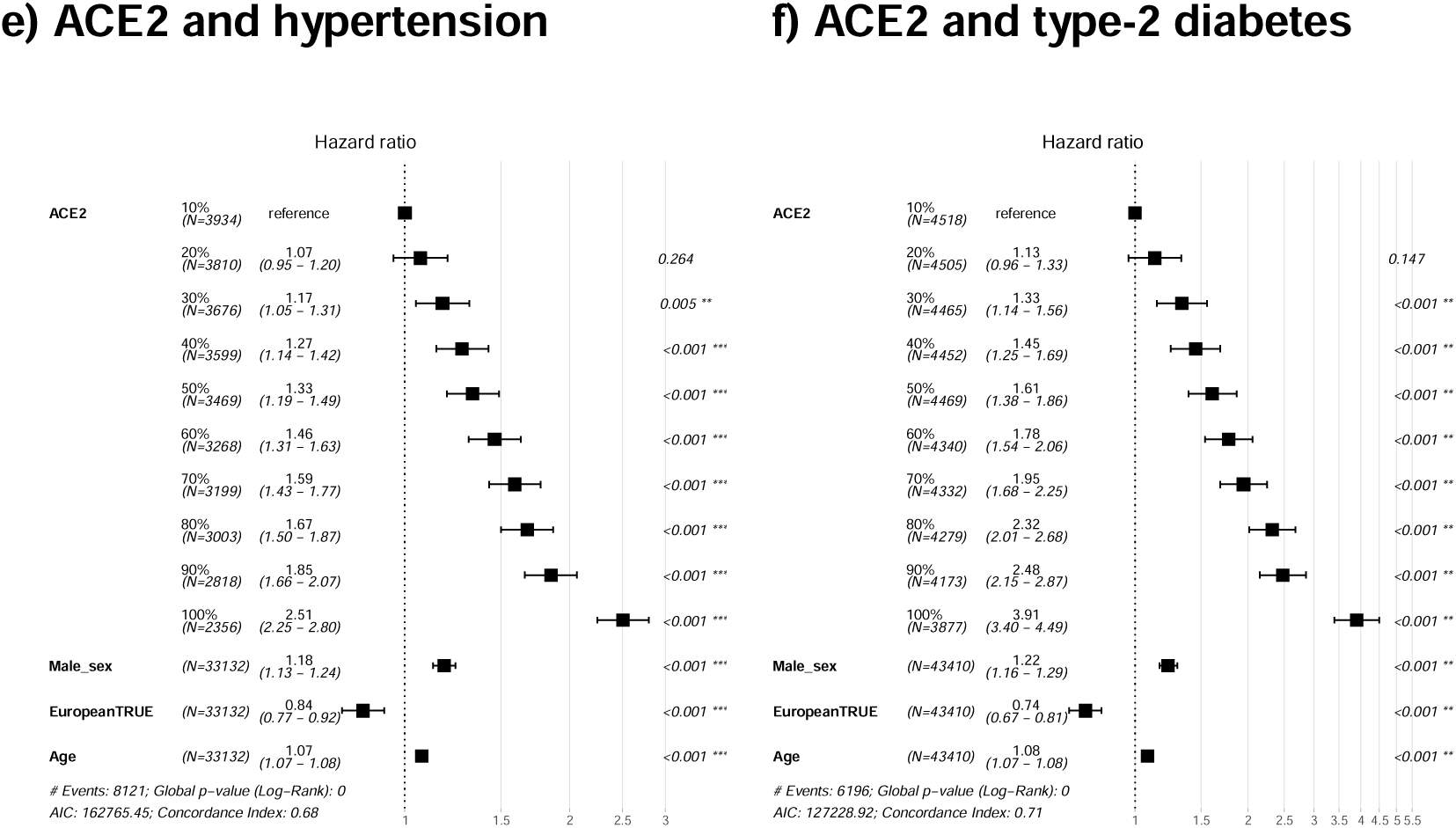
Increasing levels of NT-proBNP are observed in participants diagnosed with incident heart failure, atrial fibrillation, and myocardial infarction. Increasing levels of ACE2 are observed in participants diagnosed with incident hypertension and type-2 diabetes. **a)** The figure shows the sequential increase in mean NT-proBNP with overt diseases and other modifiers influencing the protein’s levels. This includes a heart failure (HF) diagnosis alongside sex, age at recruitment, a diagnosis of cardiomyopathy or atrial fibrillation, and carriers of pathogenic cardiomyopathy- associated variants. The Student’s t-test compared “No HF diagnosis” as the reference group. The groups contained the following sample sizes, respectively: (No HF diagnosis) 44050, (female groups:) 103, 286, 12, 33, 234, 2, (male groups:) 202, 496, 7, 42, 470, 14. NT-proBNP units are Olink’s arbitrary unit in log_2_ scale. HF, heart failure; plp, P/LP variant carrier; Afib, atrial fibrillation; CM, cardiomyopathy diagnosis. The forest plots (**b)-f)**) of Cox proportional hazards regression models were created assessing death or diagnosis from recruitment with those diagnosed before recruitment excluded. Sex (increasing risk is male), European ancestry (increasing risk is European), and age at recruitment (incremental risk per year lived), were added to this multivariable analysis for comparison. Forest plots are presented for deciles of NT- proBNP levels with incident **b)** heart failure (HF), **c)** atrial fibrillation (Afib), **d)** myocardial infarction (MI), from recruitment, and ACE2 levels by decile with incident **e)** hypertension and **f)** type-2 diabetes, from recruitment.

We showed that ACE2 was significantly increased on average in participants of African or Caribbean ancestry and some guidelines suggest ACE inhibitors (targeting the ACE1 protein) are less effective in “Black” individuals (see the **Discussion**). A high level of ACE2 at recruitment in participants of African or Caribbean ancestry was not significantly predictive of a hypertension diagnosis, however, a limitation of this experiment is the small sample size (n=682).

ACE2 levels were associated with the loci of diabetes-associated genes *HNF1A* and *HNF4A* at GWAS and RVAS (**Table S3-S4**). Increasing ACE2 levels at recruitment were predictive of an incident type-2 diabetes diagnosis (**Figure 7, Figure S15**). Two-sample Mendelian randomisation with the results of a GWAS of type-2 diabetes^27^, showed evidence for an inverse relationship: ACE2 genetic instrument decreased the risk of type-2 diabetes while type-2 diabetes increases ACE2 levels (**Figure 6, Figure S7**). The causal protective relationship of ACE2 as an exposure for type-2 diabetes can be exemplified through a missense variant (rs1169288) in the gene hepatocyte nuclear factor-1 alpha (*HNF1A*). The variant is associated with an increased risk of type-2 diabetes (GWAS β=0.04) and decreased levels of circulating ACE2 (GWAS β=-0.16).

### NT-proBNP and BNP and their role in heart failure

The GWAS of NT-proBNP identified cardiomyopathy- or heart failure-associated genetic loci (e.g., *HSPB7/CLCNKA*, *CDKN1A*, *TTN, FLNC,* and *BAG3*). BNP also associated with variants in the loci of *BAG3* at GWAS. Through RVAS, NT-proBNP and BNP associated with variants in *NPPB* and the region surrounding the natriuretic peptide genomic locus (*NPPA/NPPB*). This suggests that the natriuretic peptide locus is the major effect locus of NT-proBNP and BNP levels as the loci of the suggested furin and corin convertase enzymes involved in their biosynthesis were not identified.

NT-proBNP and BNP levels were increased in individuals with overt cardiomyopathy (**Figure S14**) and were predictive of incident diagnoses (heart failure, cardiomyopathy, and atrial fibrillation (**Figure 7, Figure S8, Figure S12, Figure S15**)). To understand whether these increased levels are disease-causing or protective in cardiomyopathies, we undertook two-sample Mendelian randomisation with the results of GWAS of HCM^28^ and DCM^5^. This analysis suggested that left ventricular hypertrophy increases NT-proBNP and BNP circulating levels (**Figures S5-S6, Table S5**). An example of this effect is the lead SNP (downstream variant rs198379) of the *NPPB* locus associated with increased NT-proBNP and BNP circulating levels, here and previously^29,30^. The variant increases *NPPB* atrial appendage expression on GTEx^9^ and increases the risk of heart failure and HCM through case-control GWAS^20,31^ (**Figure S5**).

The opposite was observed for the DCM Mendelian randomisation; common genetic variants associated with DCM risk^5^ predisposed to decreased NT-proBNP levels (**Figure S5, Table S5**). Using the same *NPPB* SNP as described as an example, the variant increased levels of NT-proBNP and BNP, and decreased the risk of DCM. Opposing genetic relationships between HCM and DCM common variants have been described elsewhere^6,28^ and indicate that genetic loci underlying the variability of left ventricular function in the general population may be differentially involved in susceptibility to HCM and DCM.

Atrial fibrillation^32^ increased NT-proBNP circulating levels observationally and through Mendelian randomisation (**Figure S10, Table S5**). However, the observed association between NT-proBNP and myocardial infarction^33^ at PheWAS (**Figure 4**) and with incident myocardial infarction risk (**Figure 7, Figure S15**) did not have evidence of causality (**Table S5, Figure S11**).

NT-proBNP had a positive, relationship with a heart failure diagnosis alongside older age, a diagnosis of cardiomyopathy or atrial fibrillation, and carriers of pathogenic cardiomyopathy-associated variants (**Figure 7, Figure S12, Figure S15**). We showed that NT-proBNP and BNP were significantly decreased on average in participants of African or Caribbean ancestry. Increasing NT-proBNP levels at recruitment in an analysis of participants of African or Caribbean ancestry only remained significantly predictive of heart failure (BNP was not significant).

## Discussion

We identified relationships between circulating proteins with cardiovascular disease and those that mediate causal relationships between cardiovascular risk factors and disease development. The results presented identified age, ancestry, and sex-specific protein biomarkers of cardiovascular disease risk. Genetic studies identified variants in *GCKR*, *APOE*, and *SERPINA1*, as modifiers for more than one cardiovascular-associated protein.

We show that variants in *BAG3* influence plasma NT-proBNP and BNP levels. We have previously described a particular common missense variant within BAG3 (C151R; rs2234962) that demonstrates BAG3’s potential cardioprotective function in GWAS of DCM, HF, and ejection fraction, alongside risk for HCM^6^. The variant is associated with proteins maintaining myofibrillar integrity and causes improved response to proteotoxic stress. BAG3’s role in the protection or risk for opposing cardiomyopathies and carcinomas holds promise for devising therapeutic interventions, diagnostics, and tailored treatments.

Six of the nine proteins assessed have been previously identified (Table S19 at reference^34^) as the strongest predictors in large-scale proteomic risk scores of atrial fibrillation (NT-proBNP, BNP), cardiomyopathy (NT-proBNP, TNNI3, BNP), heart failure (NT-proBNP), hypertension (NT-proBNP, ACE2, ACTA2), nonrheumatic mitral valve disorders (NT-proBNP, BNP), pulmonary hypertension (NT-proBNP, ACE2, BNP), stable angina (NT-proBNP), hyperplasia of prostate (NT-proBNP, ACTA2, BNP), kidney disease (NT-proBNP, BNP), infections (NT-proBNP, ACTN4, BNP), and pleural effusion (NT-proBNP).

### ACE2 and COVID-19, diabetes, and hypertension

The observation of the upregulation of ACE2 with smoking^35,36^ and alcohol^37–39^ has been identified in extensive studies of ACE2 in COVID-19 (where the COVID-19 virus was found to engage ACE2 for cellular entry). There is minimal information on the impact of this on cardiovascular disease risk. Through two-sample Mendelian randomisation, we showed that genetic instruments for decreased systolic blood pressure inversely correlated with circulating ACE2 levels. The relationship of ACE2 circulating levels with cardiovascular risk mediators and genetic factors here suggests that it may be an important target for therapeutic intervention. The increased ACE2 levels observed here with ACE inhibitor treatment (targeting the ACE1 protein) suggests that teasing apart the role of the increase in vasodilator ACE2 effects alongside the decrease in vasoconstriction ACE1 effects would aid our understanding of the success of ACE inhibitors, whether their impact is modified by ACE2, and whether the upregulation or stimulation of ACE2-induced vasodilation would be beneficial in clinical practice.

However, ACE inhibitors are thought to have less blood pressure-lowering effects and increased risk of angioedema in “Black” individuals with hypertension and international guidelines preferentially recommend diuretics and calcium channel blockers over ACE inhibitor treatment in these individuals. Concerns have been expressed with the generalisability of the guidelines and the lack of mechanistic understanding^40^. Further exploration is required into the increased average circulating ACE2 protein levels observed here in individuals of self-reported African or Caribbean ancestry.

Angiotensin receptor blockers prevent the action of angiotensin II for high blood pressure regulation, preventing heart failure, and preventing kidney failure in people with diabetes. *ACE2* increased expression in the endocrine pancreas in diabetes is hypothesized to act in a compensatory manner^41^ and here we provide evidence of genetic associations that strengthen a potential role for *ACE2* expression and ACE2 protection against diabetes. Conversely, as a biomarker, ACE2 may have potential as a measure of increased cardiovascular disease risk.

### NT-proBNP: a predictive biomarker of hypertrophic cardiomyopathy

NT-proBNP is a prohormone with an N-terminal that is cleaved to release brain or b-type natriuretic peptide 32 (BNP). BNP is released by the heart upon myocardial wall stretch; it reduces fluid and sodium retention and causes mild vasodilation, regulating blood pressure. It has been implicated in hypertrophy, fibrosis, angiogenesis, and cardiomyocyte proliferation and viability. BNP and NT-proBNP are circulating biomarkers of heart failure and hypertrophy due to the reactivation of *NPPA* and *NPPB*^42^. Here, NT-proBNP circulating levels are associated with variants in the loci of these atrial-expressed genes.

We identified an opposing relationship between HCM and DCM and the BNPs; while the levels of BNPs are increased with the progression of both cardiomyopathies, causality via Mendelian randomisation suggested that BNPs are part of HCM pathology and progression while the observed increase in DCM is an adaptive response to contractile dysfunction and cardiomyocyte stretch. The roles of BNPs in natriuresis and promoting hypertrophy may protect against DCM-associated systolic dysfunction. In models, endogenous natriuretic peptides have been shown to protect the heart in a mouse model of DCM and sudden death, *NPPB* knockout rats at 3 months showed hypertrophy without alteration to the ventricles or function, which transitioned at 6 months into DCM, and diabetic cardiomyopathy mice models treated with exogenous BNP prevented the development of DCM, while knockdown of endogenous BNP accelerated DCM^43–45^.

ARBs have been trialled as a treatment for early-stage HCM^46^ based on data that suggest they abrogated the development of hypertrophy and fibrosis. Initial results of the VANISH trial were encouraging. Other studies are investigating the combination of sacubitril-valsartan^47^ (e.g., NCT04164732). Sacubitril is a neprilysin inhibitor that inhibits the degradation of BNPs. Our results question whether this combination may have negative consequences in patients with hypertrophy, given the possible effects of increased BNP in this context. Our results are in keeping with existing data that support a positive effect in patients with heart failure with reduced ejection fraction^48^, including those with dilated cardiomyopathy. Further study is required to address this.

We show better predictive capacity for measures of circulating NT-proBNP than BNP with cardiovascular disease. While the measures are highly correlated (R=0.67), NT-proBNP is measured at a higher concentration, has a higher prognostic value^49^, has been shown to have sustained elevation for 12 weeks^50^, the predictive capacity of the ratio of NT-proBNP:BNP has been explored previously^51^, we show that NT-proBNP was more predictive of heart failure, other cardiovascular diseases, and risk factors, and is more influenced by genetic factors (heritability) than BNP, suggesting important differences in measurable levels. NT-proBNP is inactive and has a longer half-life than BNP which likely explains the improved prediction capacity and increased heritability of plasma measures of NT-proBNP identified here.

The relationships identified with sex, ancestry, and specific cardiovascular diseases, may have implications for NT-proBNP’s predictive capacity in clinical settings. It could be suggested that the association of NT-proBNP levels with female sex may be due to lower BMI, but adjustment for BMI did not alter this finding. NT-proBNP >125 pg/mL is common in females without classical cardiovascular risk factors as well as older people^52^. The alteration in average NT-proBNP levels with ancestry is thought to convey an altered risk for hypertension^53,54^, but as average levels are lower in individuals of Chinese and African ancestry, NT-proBNP may not have similar predictive capacity across ancestries. Analyses of proteomics in more diverse ancestries would aid this assessment.

Altered NT-proBNP has been previously noted in patients with atrial fibrillation^55^, which may influence heart failure or cardiac event risk prediction capabilities in atrial fibrillation patients. Enlargement of the left atrium has been shown to increase NT-proBNP and BNP in individuals diagnosed with atrial fibrillation^56^. Enlarged atria are associated with both existing and incident atrial fibrillation and atrial fibrillation and heart failure have shared pathogenesis. The association of NT-proBNP (and BNP) with future uptake of anticoagulant medication is likely due to a diagnosis of atrial fibrillation or flutter.

The increase of NT-proBNP circulating levels in participants with a pathogenic/likely pathogenic cardiomyopathy-associated variant may be indicative of individuals at particular risk of cardiomyopathy, incident heart failure, or atrial fibrillation. The association of NT-proBNP circulating levels with sinus bradycardia has been identified previously and thought to be through increased stroke volume and wall tension^57^, and we show evidence here that it is unlikely to be due to beta-blockers prescribed for hypertension, as previously suggested^58^.

### Limitations

There are several limitations to this study. The UKB has biases (survivorship, dominated by European ancestry, etc.). Assessing single time point data limits assessments of causality and directionality. While informative, assessments of medication use require longitudinal follow-up, and the medication associations described may be due to the diagnoses they were prescribed for. The proteomics was measured in samples collected at baseline recruitment while the imaging appointment was on average 8 years later and sensitivity analyses adjusting for this have been untaken. Participants with both proteomics and imaging is currently limited.

### Conclusions

We describe the relationships between nine plasma proteins with roles in genetic or structural cardiovascular disease or treatment pathways, that may mediate relationships between cardiovascular risk factors and disease development. We discuss the potential for additional avenues of therapeutic intervention with studies of ACE2 in hypertension and diabetes, and BAG3 in cardiomyopathies, and the need to understand the relationships with NT-proBNP for diagnostic purposes in stratified groups of patients. This study provides an improved understanding of the circulating pathways depicting cardiovascular disease dynamics and the influencing modifiers and risk factors.

### Sources of funding

K.A.M. is supported by the British Heart Foundation [BHF; FS/IPBSRF/22/27059, RE/18/4/34215]. L.C. is supported by the BHF [RE/18/4/34215]. A.S. is supported by the BHF [FS/CRTF/21/24183] and a National Institute for Health Research (NIHR) Clinical Lectureship. F.S.N. is supported by the BHF [RG/F/22/110078, RE/18/4/34215, RE/24/130023]. B.H. is funded by the BHF [FS/ICRF/21/26019] and Rosetrees Trust. J.S.W. is supported by the Sir Jules Thorn Charitable Trust [21JTA], the Medical Research Council (UK) [MC_UP_1605/13], and the BHF [RG/19/6/34387, RE/18/4/34215]. D.P.O is supported by the Medical Research Council [MC_UP_1605/13], and the BHF [RG/19/6/34387, RE/18/4/34215, CH/P/23/80008]. All authors were supported by the National Institute for Health Research (NIHR) Imperial College Biomedical Research Centre. The views expressed in this work are those of the authors and not necessarily those of the funders. For open access, the authors have applied a CC BY public copyright license to any Author Accepted Manuscript version arising from this submission. The graphical abstract was created with Biorender.com.

### Disclosures

K.A.M. has consulted for Checkpoint Capital LP. D.P.O’R has consulted for Bayer AG and Bristol Myers-Squibb. J.S.W. has consulted for MyoKardia, Inc., Pfizer, Foresite Labs, Health Lumen, and Tenaya Therapeutics, and has received research support from Bristol Myers-Squibb. None of these activities are directly related to the work presented here. All other authors have nothing to disclose.

### Data availability

All data included in this study is available through the UK Biobank (https://biobank.ndph.ox.ac.uk/showcase/) and GWAS results will be made available through the GWAS catalog (https://www.ebi.ac.uk/gwas/).

### CrediT statement

Conceptualization and guarantor: K.A.M.; Formal analysis: K.A.M.; Resources: L.C., A.S., F.S.N., J.S.W., D.P.O.; Writing – original draft: K.A.M.; Writing – review & editing: (all authors).

## Supplementary information

### Supplementary figures

**Figure S1.**
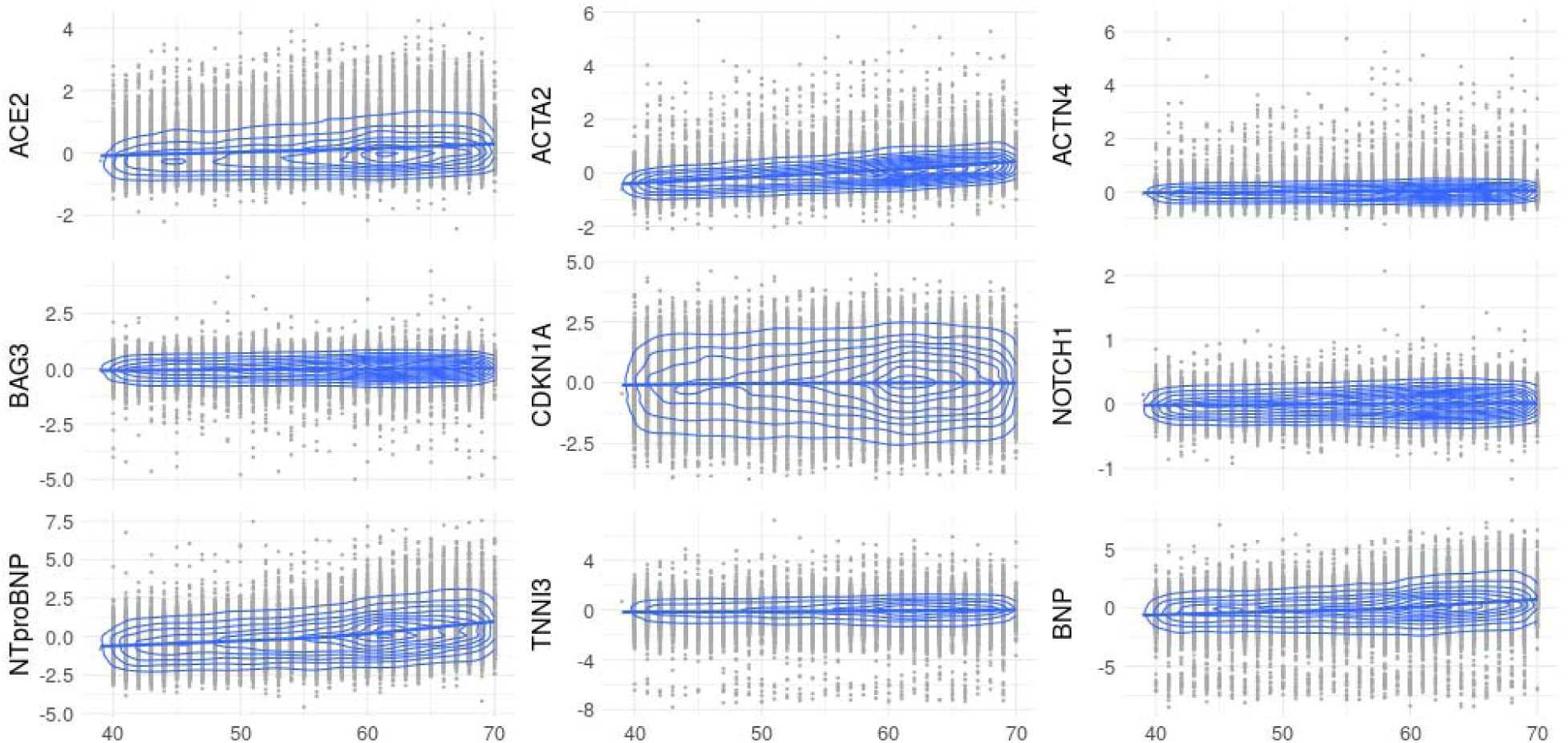
The relationships with age. The plots depict the relationships between age at recruitment (x-axis, years) and the nine plasma protein levels. Pearson’s correlation coefficient between the protein levels and age were as follows; R=0.16 ACE2, R=0.42 ACTA2, R=0.34 NT-proBNP, R=0.06 ACTN2, R=0.06 BAG3, R=0.02 CDKN1A, R=0.06 NOTCH1, R=0.06 TNNI3, and R=0.23 BNP.

**Figure S2.**
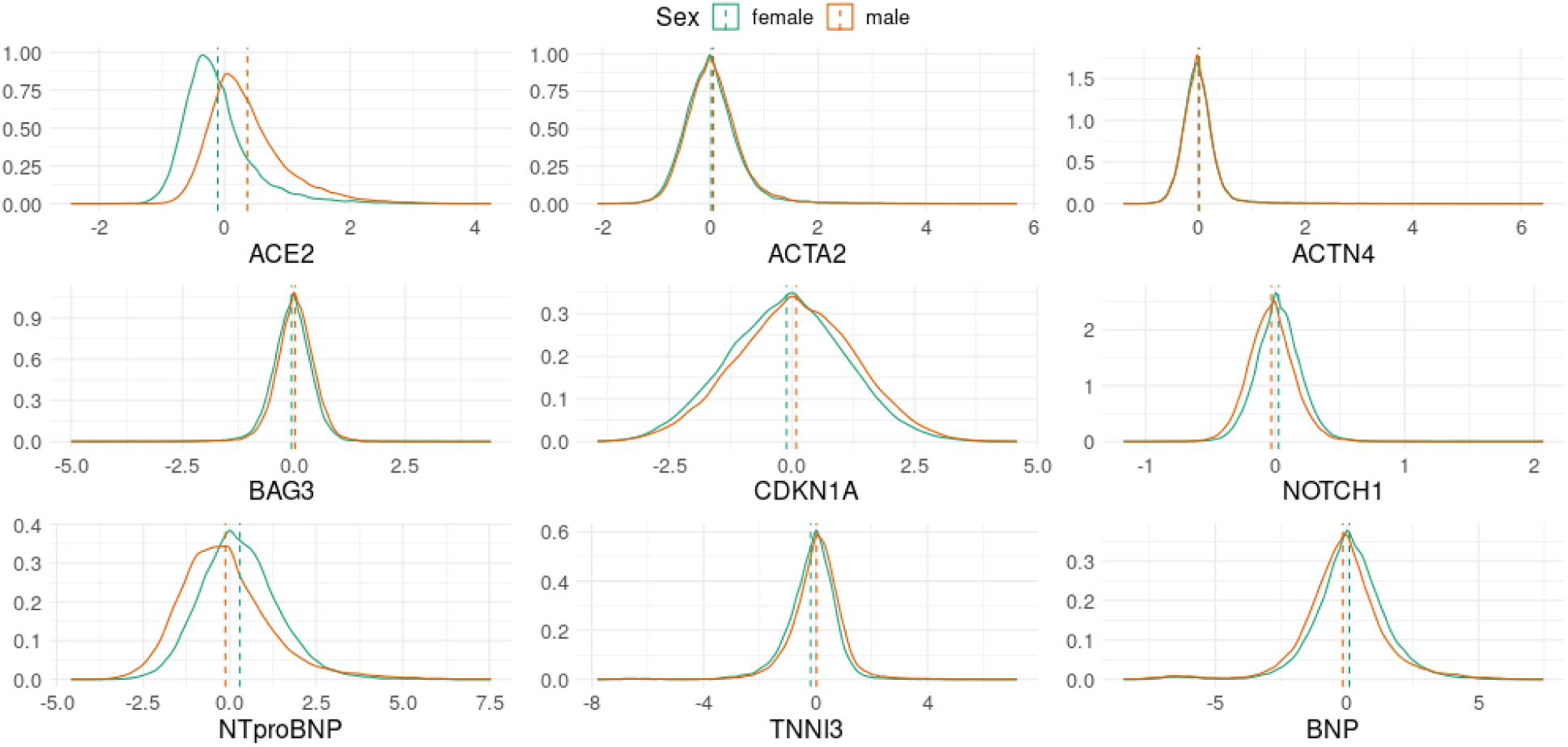
The relationships with sex. The distributions depict the relationships between sex and the nine plasma protein levels. ACE2 (P=1.0x10^-16^, β=0.47), ACTA2 (P=1.08x10^-13^, β=0.04), BAG3 (P=4.97x10^-85^, β=0.08), CDKN1A (P=6.61x10^-68^, β=0.20), and TNNI3 (P=3.97x10^-1^^27^, β=0.21) were significantly increased with male sex. NOTCH1 (P=8.61x10^-2^^58^, β=0.06), NT-proBNP (P=1.62x10^-278^, β=0.42), and BNP (P=3.50x10^-69^, β=0.25), were significantly increased with female sex.

**Figure S3.**
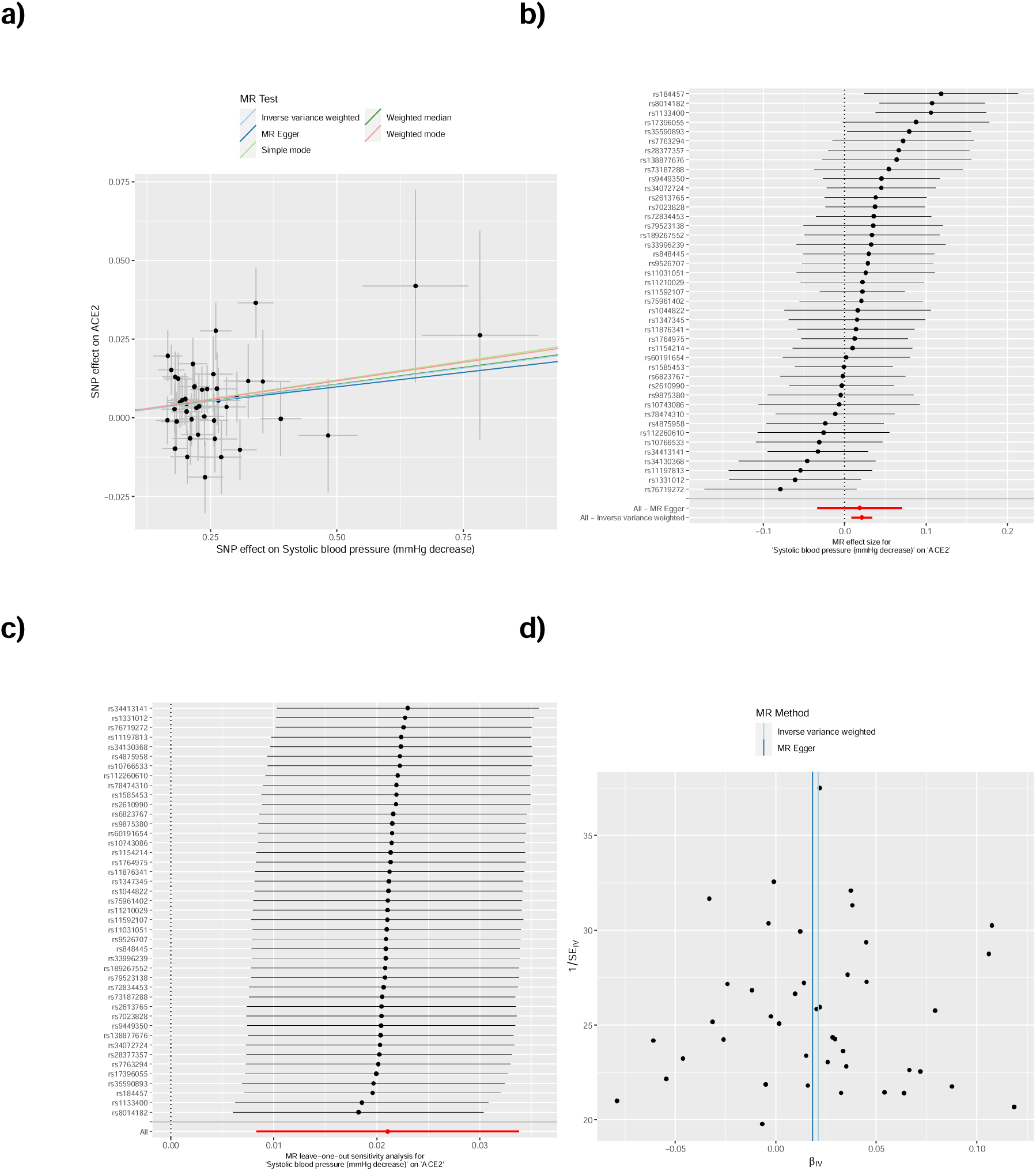
Mendelian randomization analysis of decreased systolic blood pressure as an exposure for ACE2 levels. The plots show summary information on the analyses, performed as per the TwoSampleMR R package. **a)** Mendelian randomization scatter plot for decreased systolic blood pressure as an exposure for ACE2. **b)** Mendelian randomization single SNP funnel plot for decreased systolic blood pressure as an exposure for ACE2. **c)** Mendelian randomization single SNP forest plot for decreased systolic blood pressure as an exposure for ACE2. **d)** Mendelian randomization leave one out plot for decreased systolic blood pressure as an exposure for ACE2.

**Figure S4.**
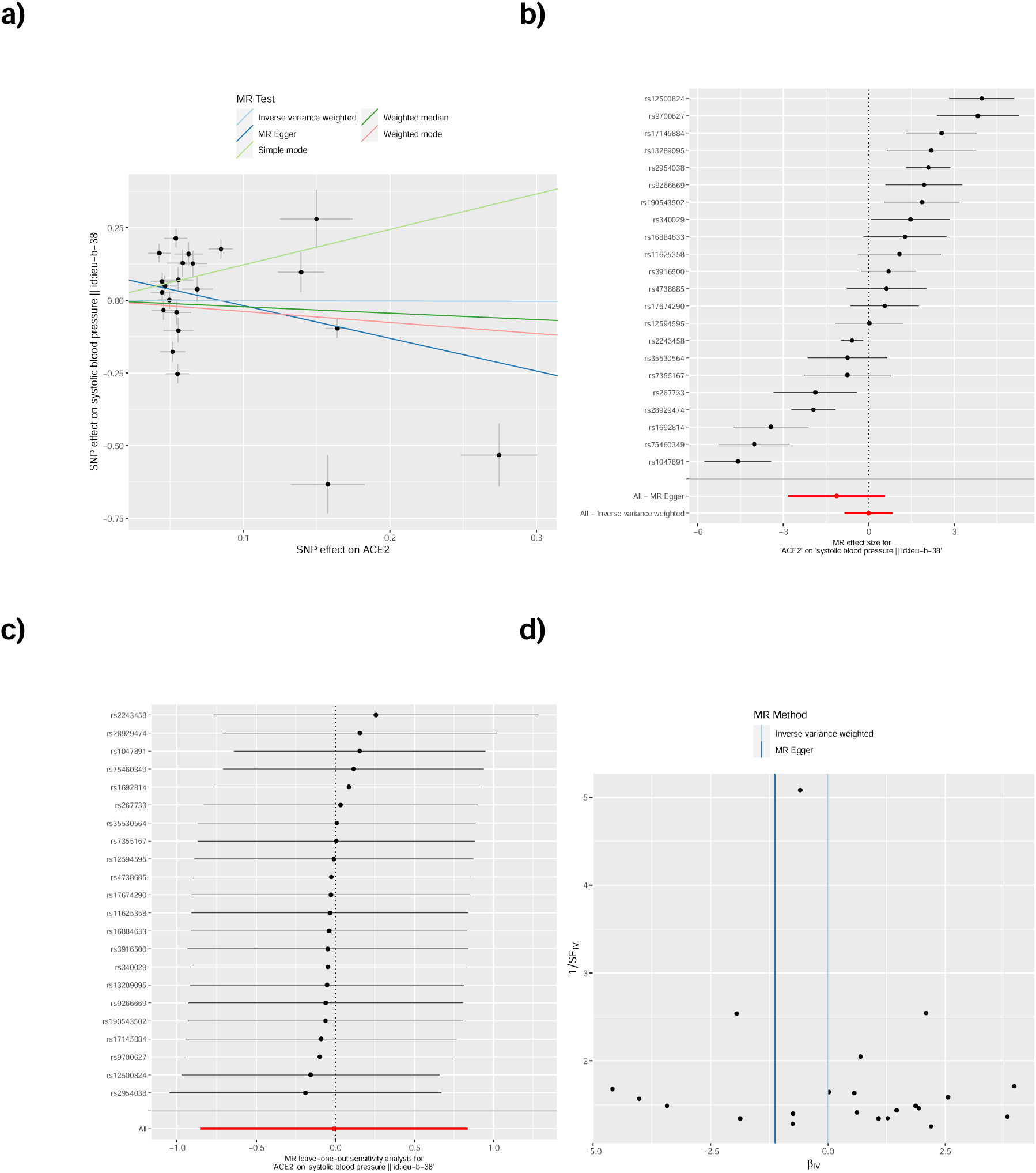
Mendelian randomization analysis of ACE2 levels as an exposure for systolic blood pressure. The plots show summary information on the analyses, performed as per the TwoSampleMR R package. **a)** Mendelian randomization scatter plot for systolic blood pressure as an outcome of ACE2 exposure. **b)** Mendelian randomization single SNP funnel plot for systolic blood pressure as an outcome of ACE2 exposure. **c)** Mendelian randomization single SNP forest plot for systolic blood pressure as an outcome of ACE2 exposure. **d)** Mendelian randomization leave one out plot for systolic blood pressure as an outcome of ACE2 exposure.

**Figure S5.**
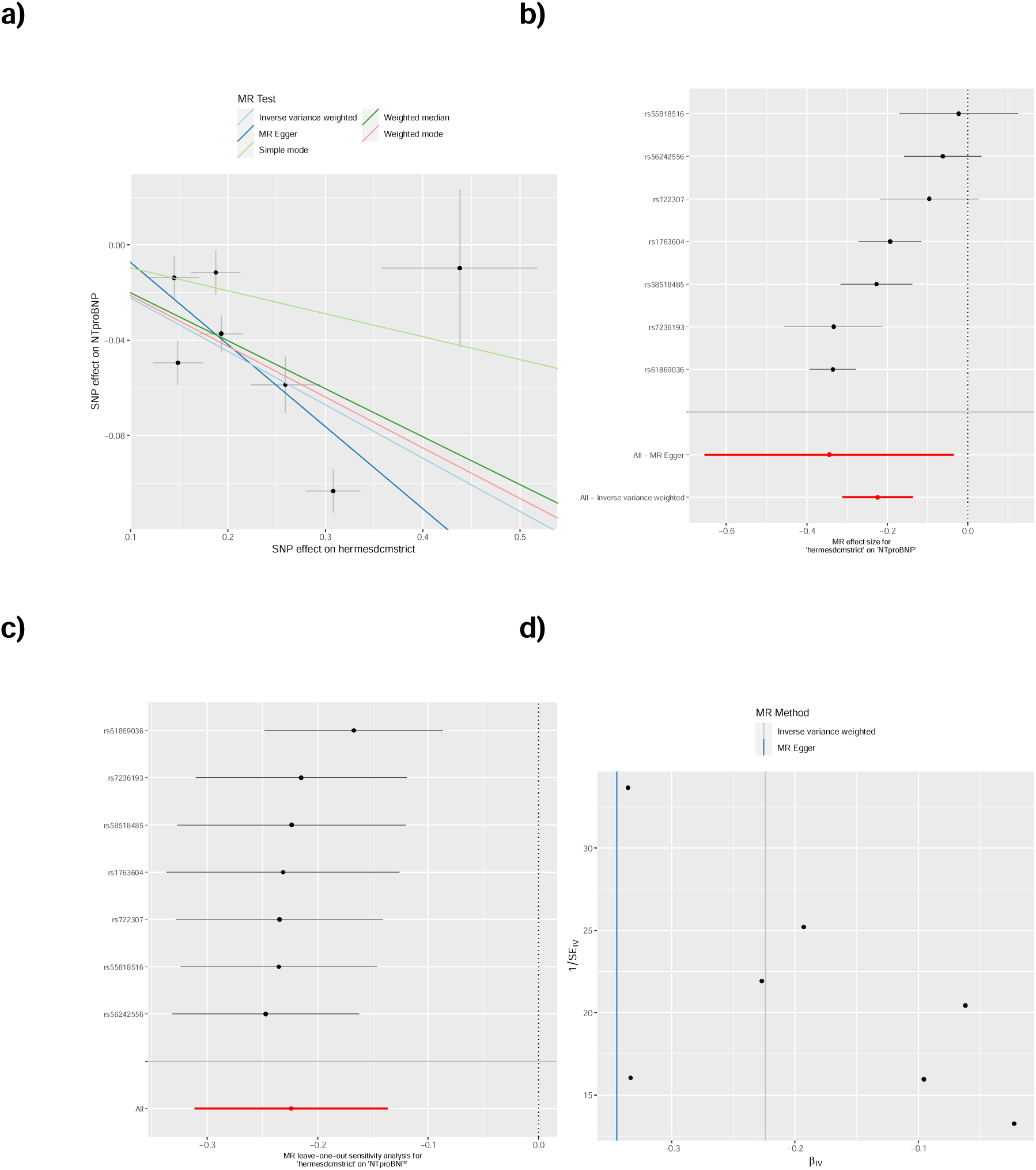
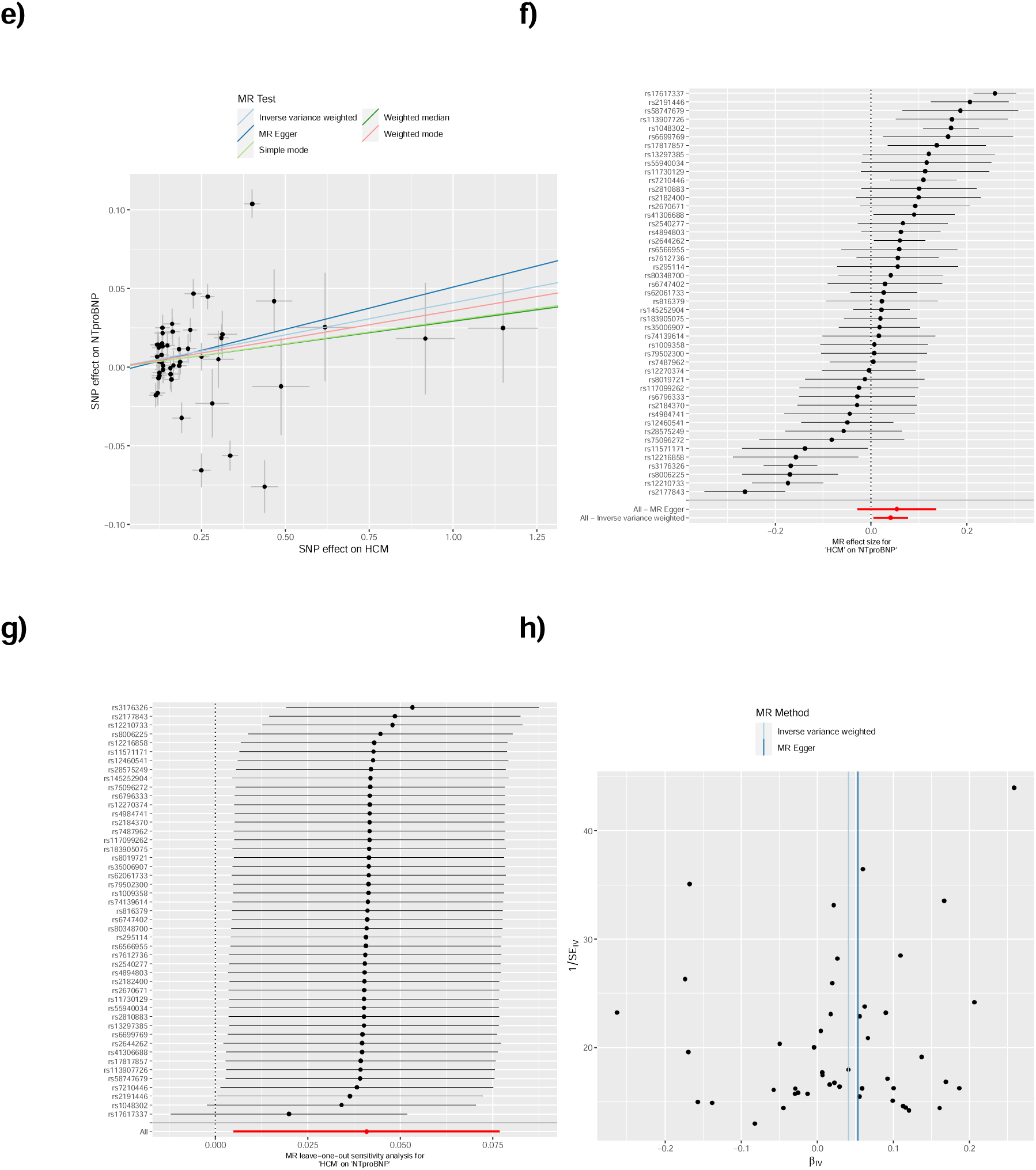
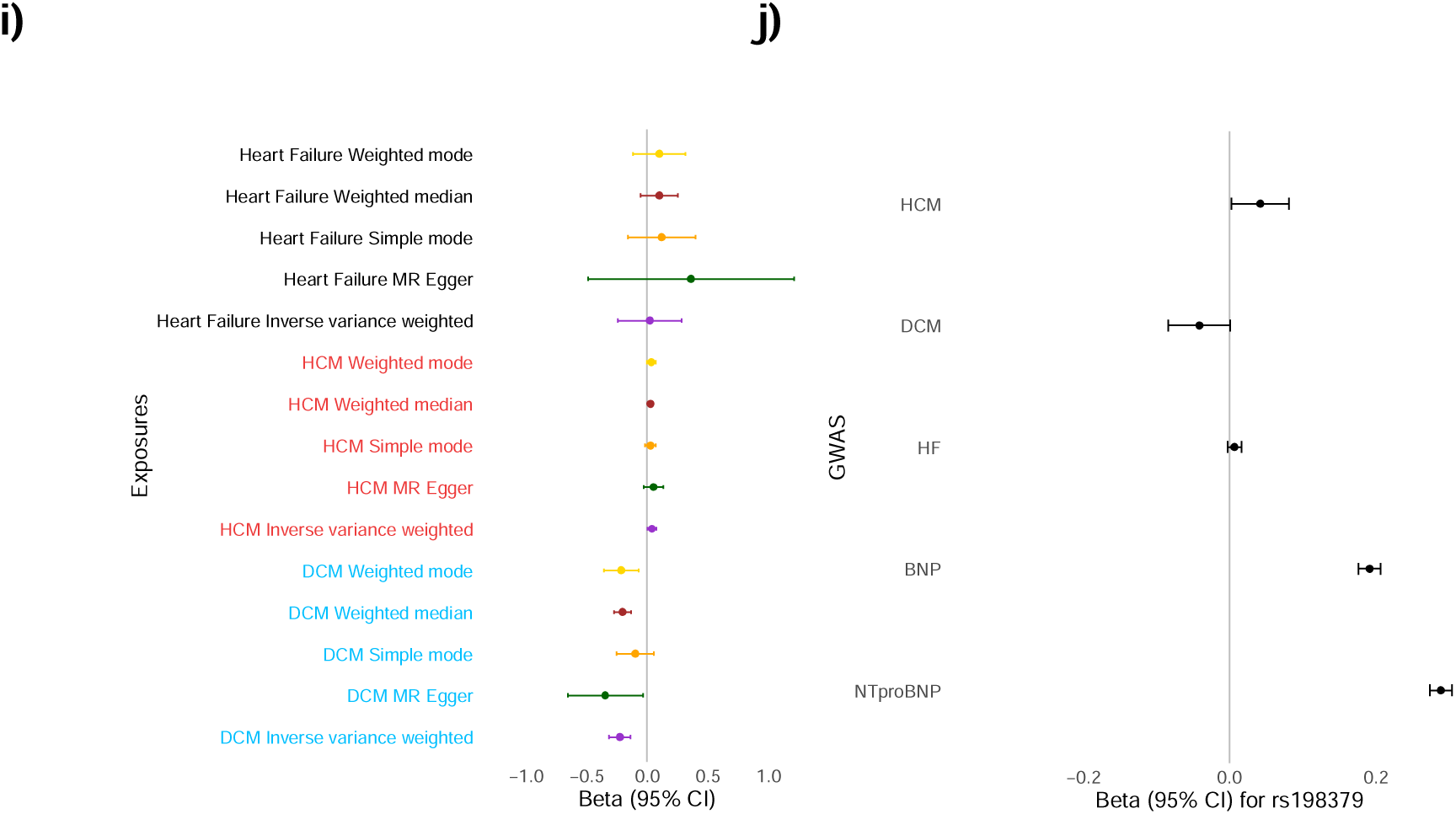
Mendelian randomization analysis of cardiomyopathies as an exposure for NT-proBNP levels. Mendelian randomisation for cardiomyopathy exposures: **a)-d)** DCM and **e)-h)** HCM, for NT-proBNP outcome. Summary plots are presented in **i)-j)**. Left ventricular hypertrophy increases NT-proBNP and BNP circulating levels, with the effect mainly through variants in *BAG3* and *CLCNKA.* The plots show summary information on the analyses, performed as per the TwoSampleMR R package. **a)** Mendelian randomization scatter plot for dilated cardiomyopathy (DCM) as an exposure for NT-proBNP. **b)** Mendelian randomization single SNP funnel plot for DCM as an exposure for NT-proBNP. **c)** Mendelian randomization single SNP forest plot for DCM an exposure for NT-proBNP. **d)** Mendelian randomization leave one out plot for DCM as an exposure for NT-proBNP. **e)** Mendelian randomization scatter plot for hypertrophic cardiomyopathy (HCM) as an exposure for NT-proBNP. **f)** Mendelian randomization single SNP funnel plot for HCM as an exposure for NT-proBNP. **g)** Mendelian randomization single SNP forest plot for HCM as an exposure for NT-proBNP. **h)** Mendelian randomization leave one out plot for HCM as an exposure for NT-proBNP. **i)** Mendelian randomization genetic determination model of CM and HF genetic instruments as exposures for NT-proBNP outcome. **j)** The effect size of the lead *NPPB* SNP (downstream variant rs198379) identified here to associate at GWAS with increased NT-proBNP and BNP, and from GWAS summary statistics of published case-control studies of cardiomyopathies and heart failure. The eQTL variant increases *NPPB* expression in the atria (GTEx). The results suggest that the variant increases NT-proBNP and BNP production, the risk of HCM and heart failure, and decreases the risk of DCM. See **Table S5** for further details.

**Figure S6.**
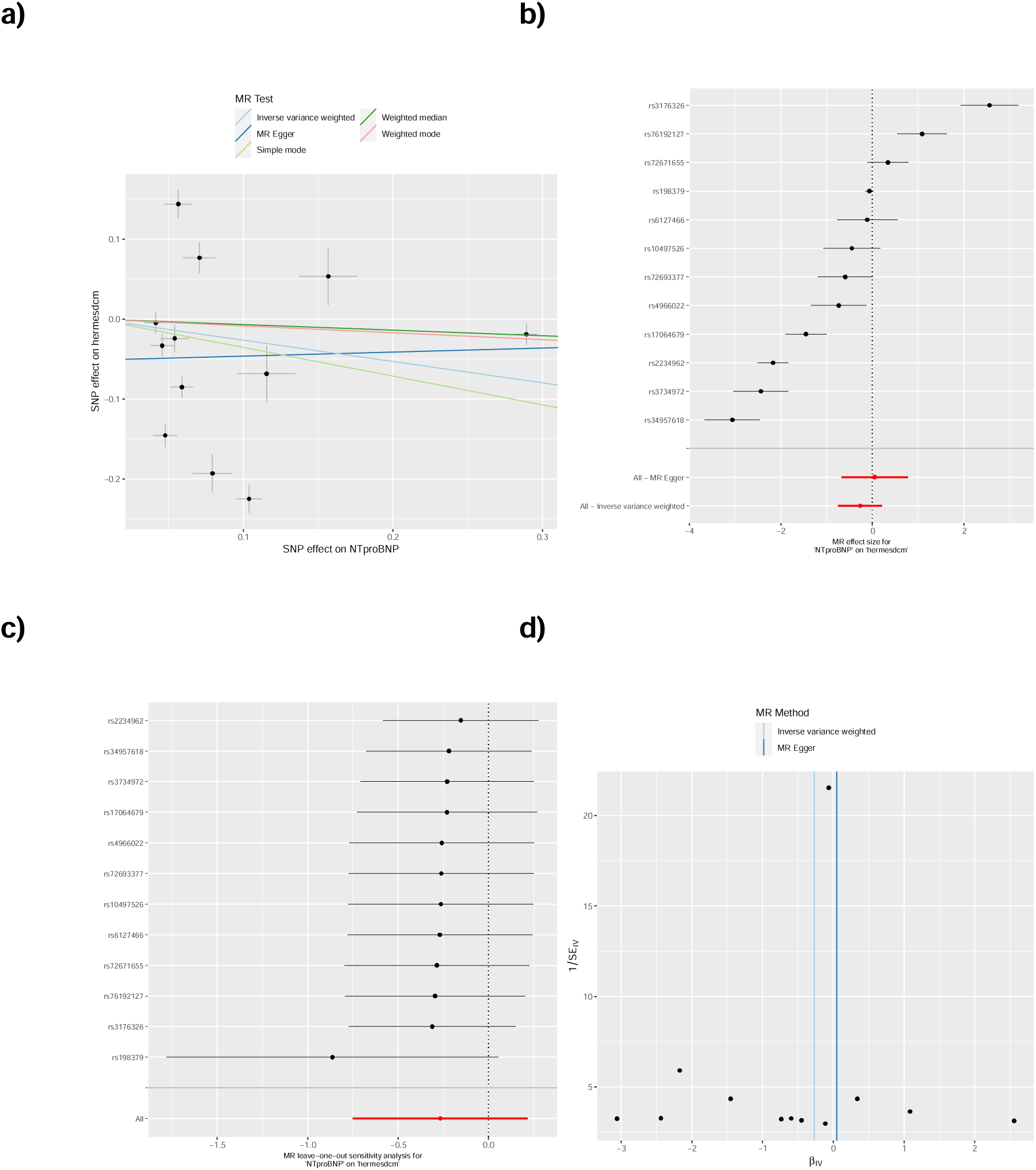
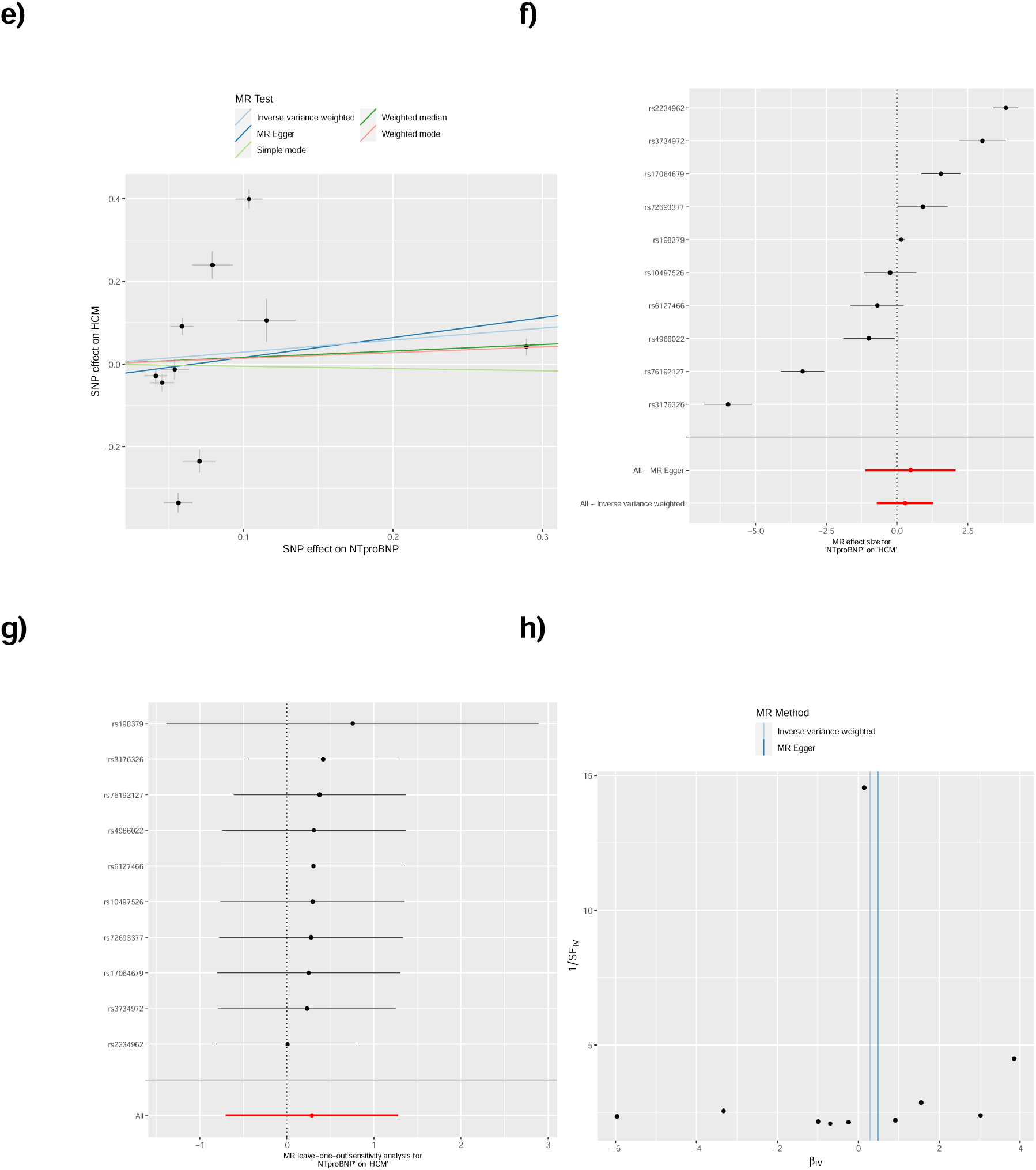
Mendelian randomization analysis of NT-proBNP as an exposure for cardiomyopathies. Mendelian randomisation for cardiomyopathy outcomes: **a)-d)** DCM and **e)-h)** HCM, for NT-proBNP exposure levels. The plots show summary information on the analyses, performed as per the TwoSampleMR R package. **a)** Mendelian randomization scatter plot for NT-proBNP as an exposure for dilated cardiomyopathy (DCM). **b)** Mendelian randomization single SNP funnel plot for NT-proBNP as an exposure for DCM. **c)** Mendelian randomization single SNP forest plot for NT-proBNP as an exposure for DCM. **d)** Mendelian randomization leave one out plot for NT-proBNP as an exposure for DCM. **e)** Mendelian randomization scatter plot for NT-proBNP as an exposure for hypertrophic cardiomyopathy (HCM). **f)** Mendelian randomization single SNP funnel plot for NT-proBNP as an exposure for HCM. **g)** Mendelian randomization single SNP forest plot for NT-proBNP as an exposure for HCM. **h)** Mendelian randomization leave one out plot for NT-proBNP as an exposure for HCM.

**Figure S7.**
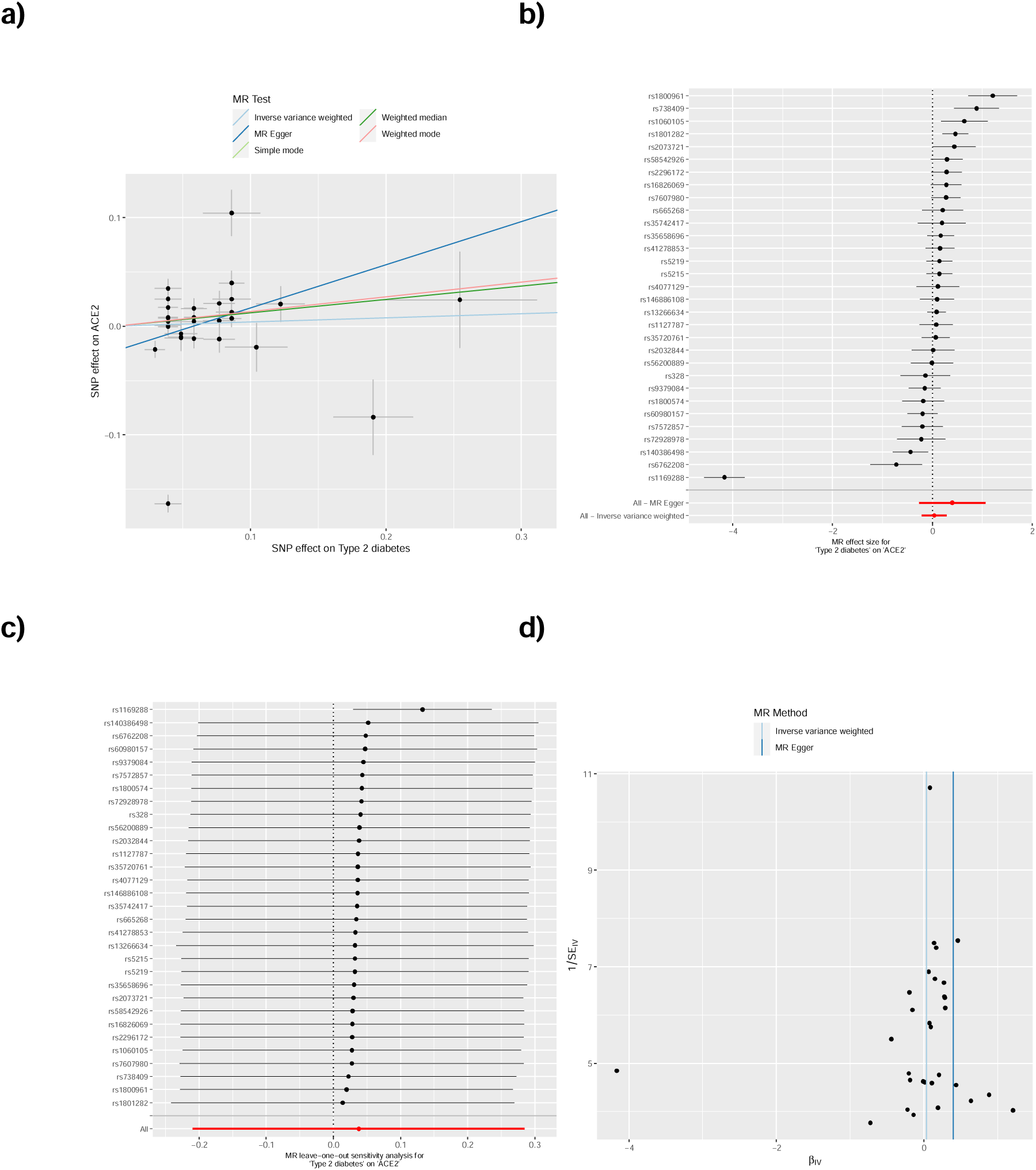
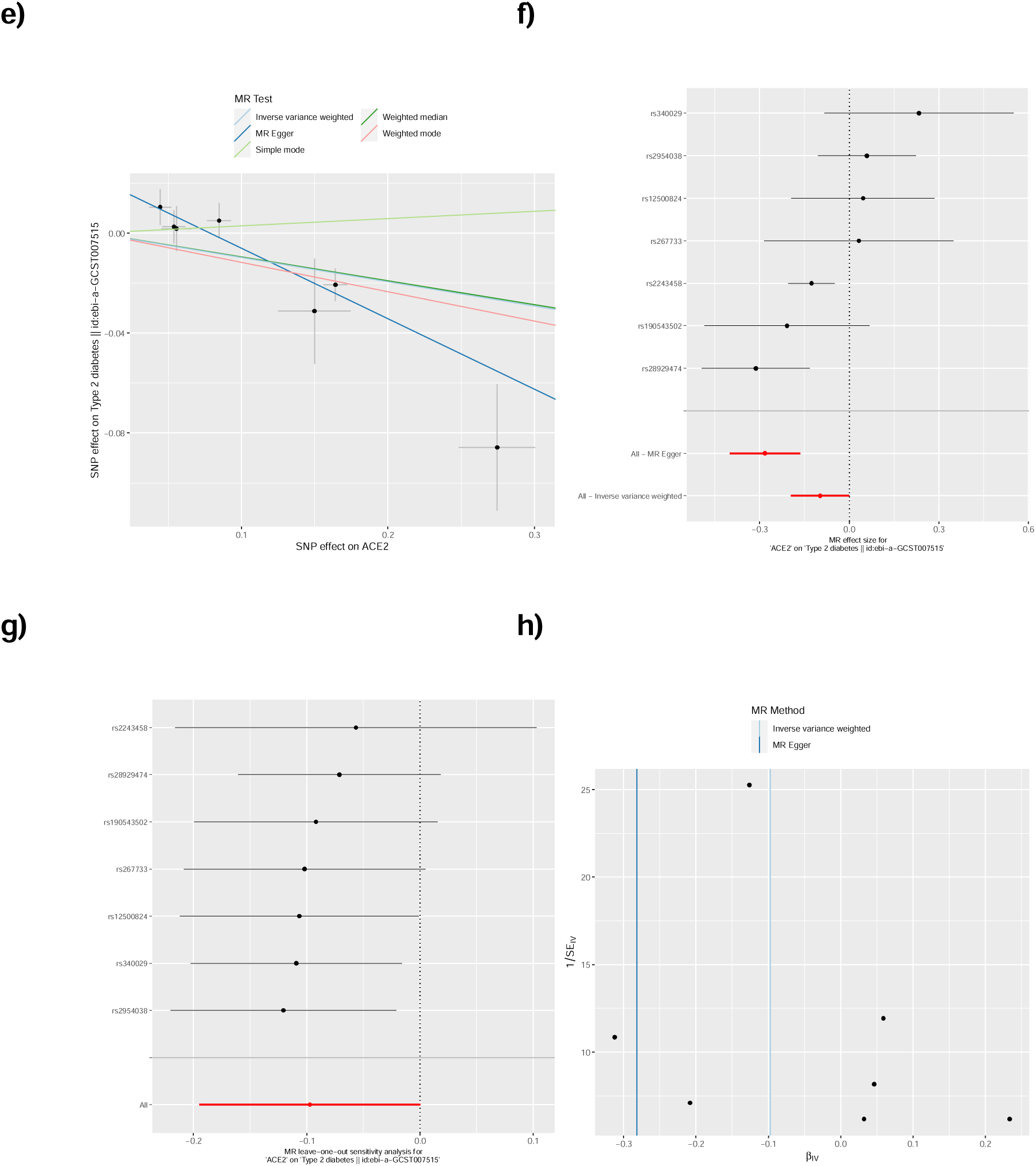
Mendelian randomization analysis of type-2 diabetes and ACE2 levels. Mendelian randomisation for type-2 diabetes as an exposure **a)-d)** and outcome **e)-h)** for ACE2 levels. The plots show summary information on the analyses, performed as per the TwoSampleMR R package. **a)** Mendelian randomization scatter plot for type-2 diabetes (T2D) as an exposure for ACE2. **b)** Mendelian randomization single SNP funnel plot for T2D as an exposure for ACE2. **c)** Mendelian randomization single SNP forest plot for T2D as an exposure for ACE2. **d)** Mendelian randomization leave one out plot for T2D as an exposure for ACE2. **e)** Mendelian randomization scatter plot for ACE2 as an exposure for T2D. **f)** Mendelian randomization single SNP funnel plot for ACE2as an exposure for T2D. **g)** Mendelian randomization single SNP forest plot for ACE2as an exposure for T2D. **h)** Mendelian randomization leave one out plot for ACE2 as an exposure for T2D.

**Figure S8.**
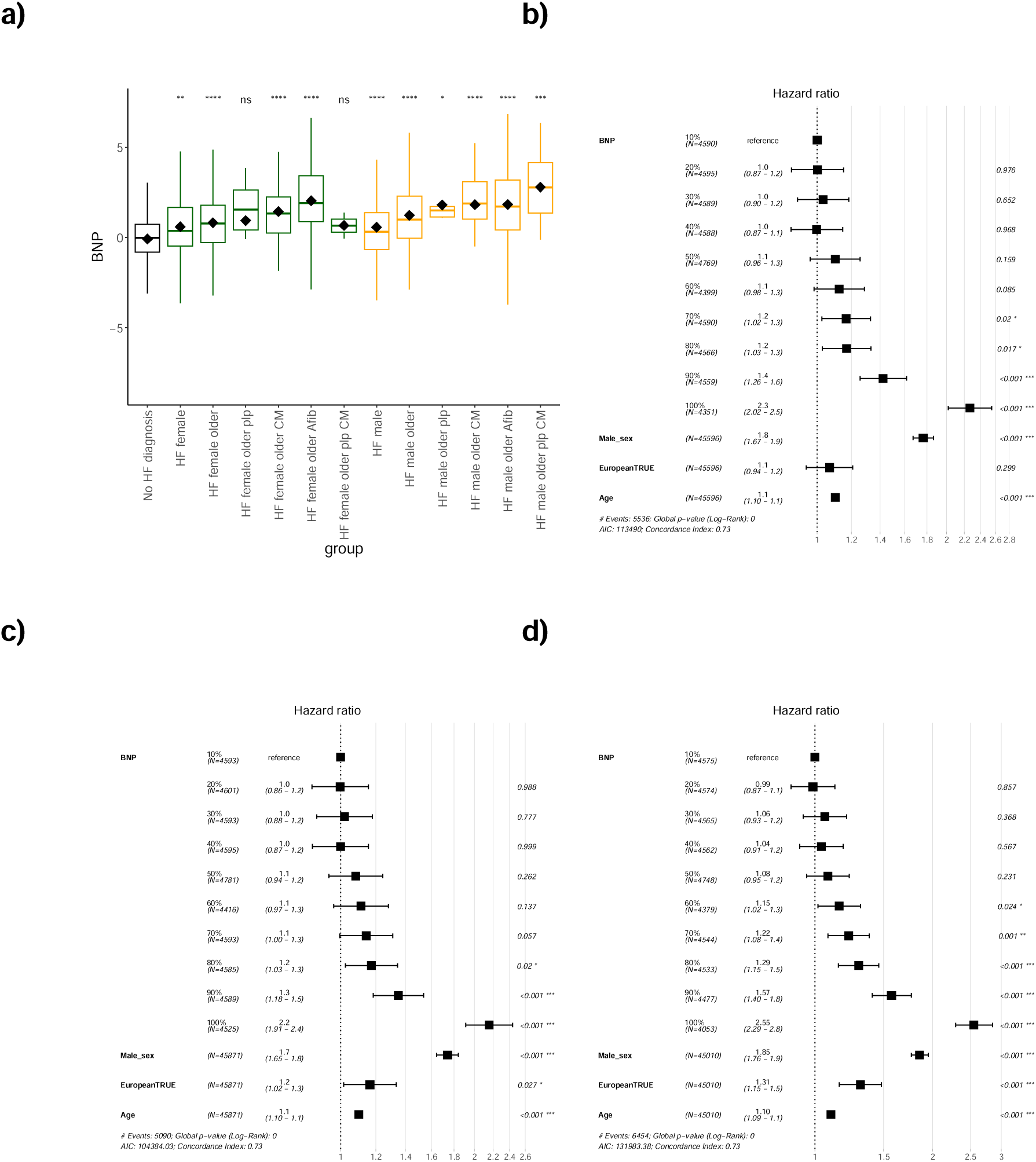
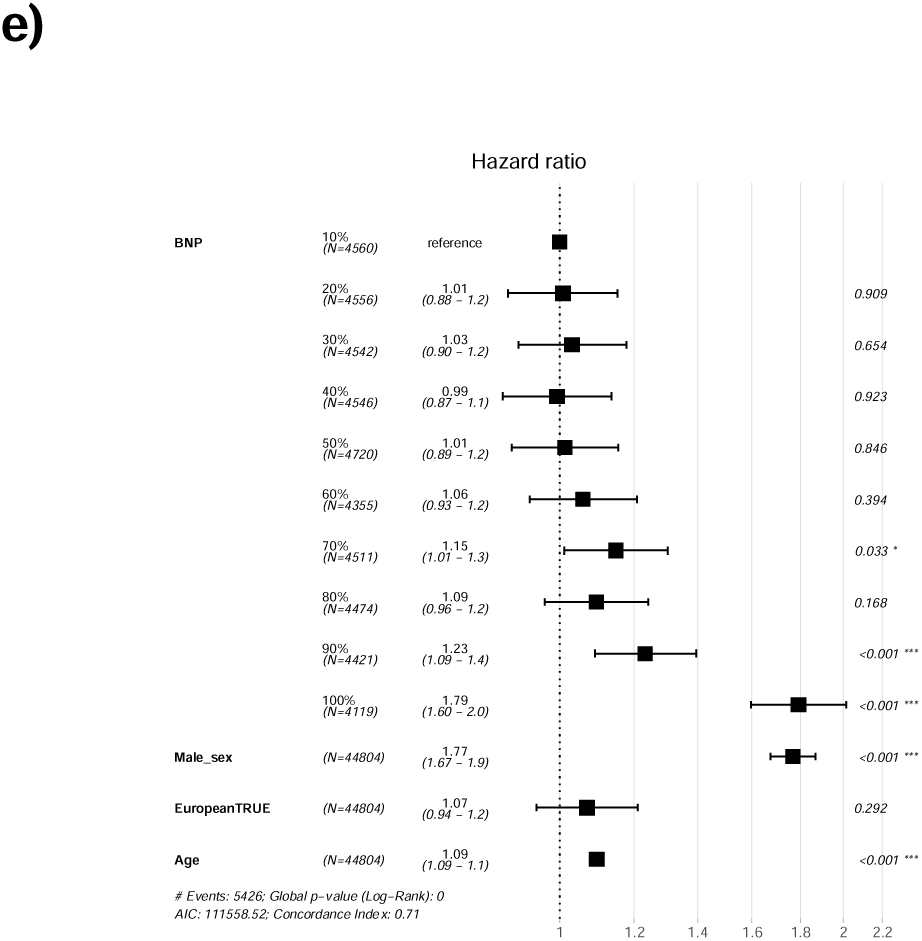
Increasing levels of BNP are observed in participants diagnosed with incident heart failure, cardiomyopathies, atrial fibrillation, and myocardial infarction. **a)** The figure shows the sequential increase in mean BNP with a heart failure diagnosis alongside sex, age, a diagnosis of cardiomyopathy or atrial fibrillation, and carriers of pathogenic cardiomyopathy-associated variants. The Student’s t-test significance was using “No HF diagnosis” as the reference group. The groups contained the following sample sizes, respectively: (No HF diagnosis) 44050, (female groups:) 103, 286, 12, 33, 234, 2, (male groups:) 202, 496, 7, 42, 470, 14. NT-proBNP units are Olink’s arbitrary unit in log_2_ scale. HF, heart failure; plp, P/LP variant carrier; Afib, atrial fibrillation; CM, cardiomyopathy diagnosis. **b, c, d)** Forest plots of Cox proportional hazards regression models for deciles of BNP levels with incident b) heart failure, c) cardiomyopathy, d) atrial fibrillation, and e) myocardial infarction, since recruitment. The forest plots were created assessing death or diagnosis from recruitment with those diagnosed before recruitment excluded. Sex (increasing risk is male), European ancestry (increasing risk is European), and age at recruitment, were added to this multivariable analysis.

**Figure S9.**
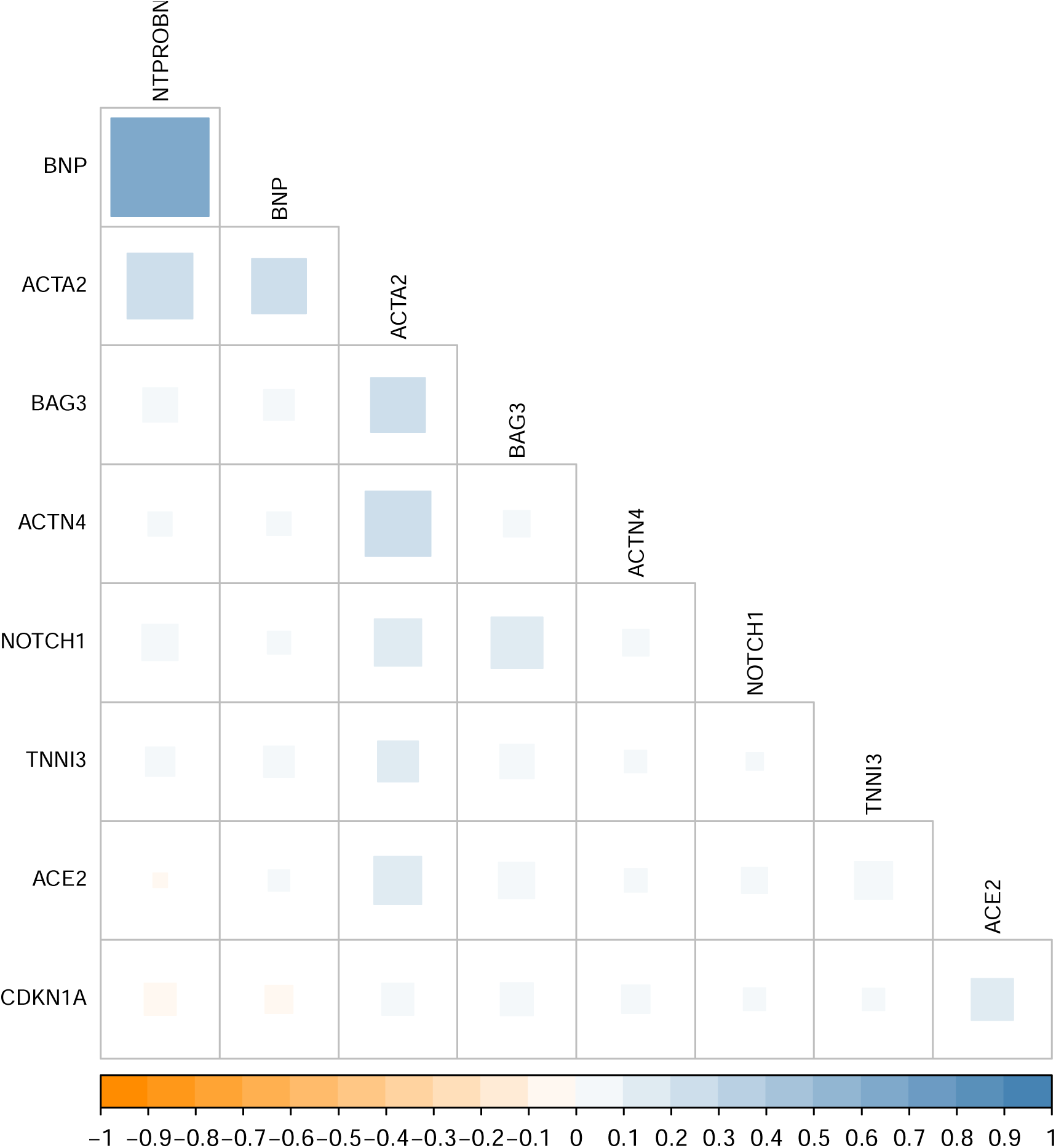
Correlation between nine circulating protein levels. The plot depicts the Pearson’s correlation coefficient (R) between the raw levels of the circulating proteins analysed. ACTA2 had the most relationships. BNP and NT-proBNP correlated with the strongest relationship to R=0.67.

**Figure S10.**
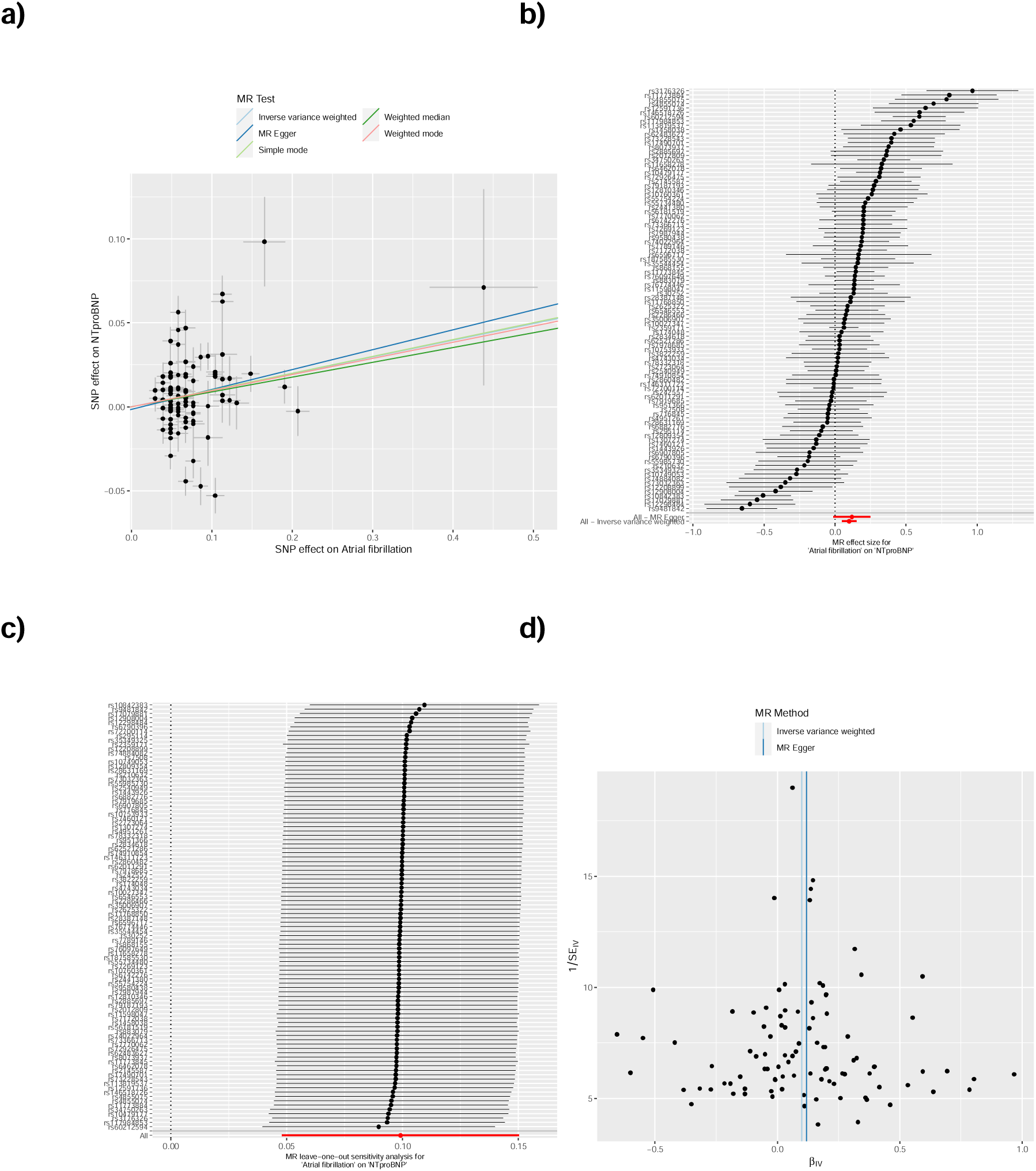
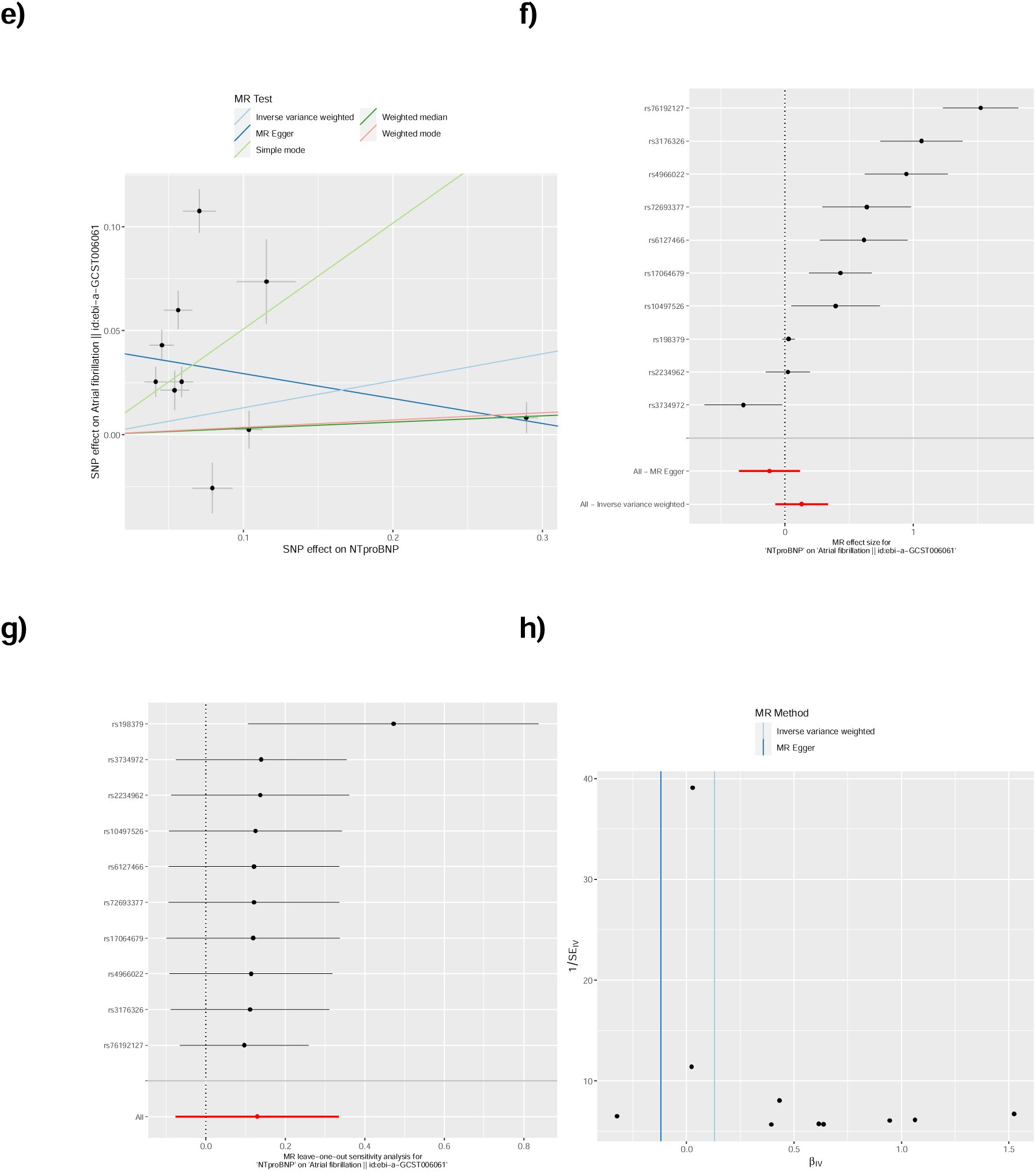
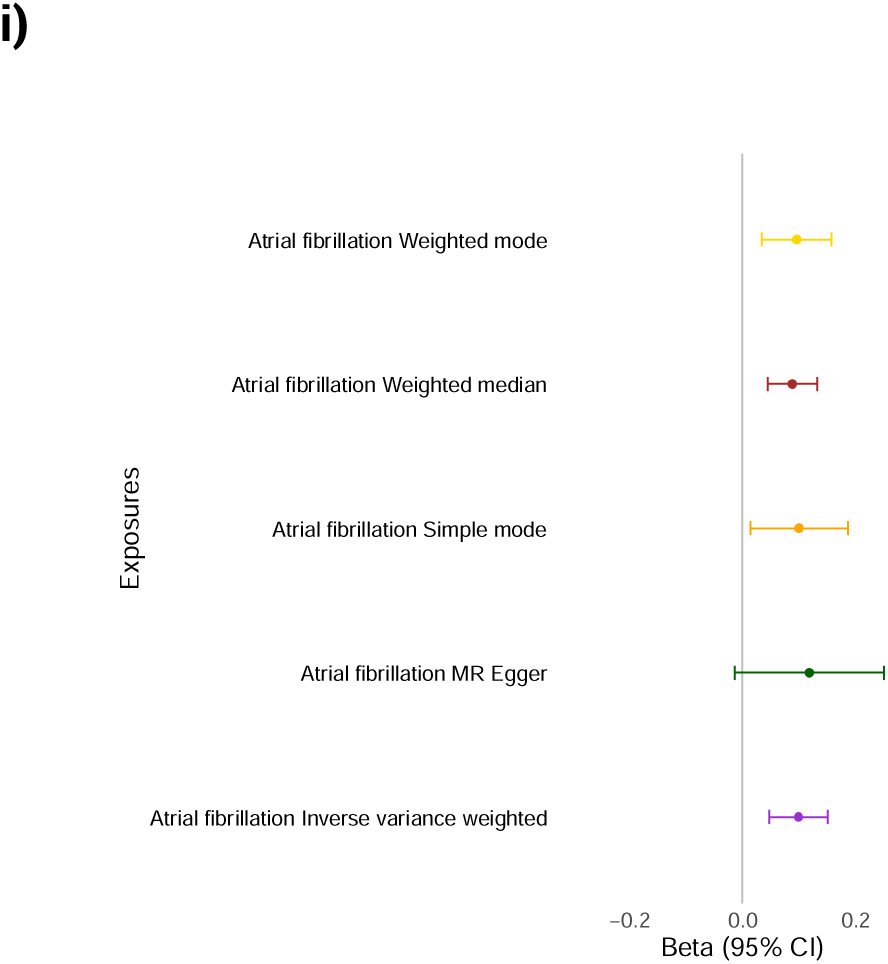
Mendelian randomization analysis of atrial fibrillation and NT-proBNP levels. Mendelian randomisation for atrial fibrillation (afib) as an exposure **a)-d)** and outcome **e)-h)** for NT-proBNP levels. The plots show summary information on the analyses, performed as per the TwoSampleMR R package. **a)** Mendelian randomization scatter plot for afib as an exposure for NT-proBNP. **b)** Mendelian randomization single SNP funnel plot for afib as an exposure for NT-proBNP. **c)** Mendelian randomization single SNP forest plot for afib as an exposure for NT-proBNP. **d)** Mendelian randomization leave one out plot for afib as an exposure for NT-proBNP. **e)** Mendelian randomization scatter plot for NT-proBNP as an exposure for afib. **f)** Mendelian randomization single SNP funnel plot for NT-proBNP as an exposure for afib. **g)** Mendelian randomization single SNP forest plot for NT-proBNP as an exposure for afib. **g)** Mendelian randomization leave one out plot for NT-proBNP as an exposure for afib. **h)** Summary plot of Mendelian randomization genetic determination model of atrial fibrillation genetic instruments as exposures for NT-proBNP outcome.

**Figure S11.**
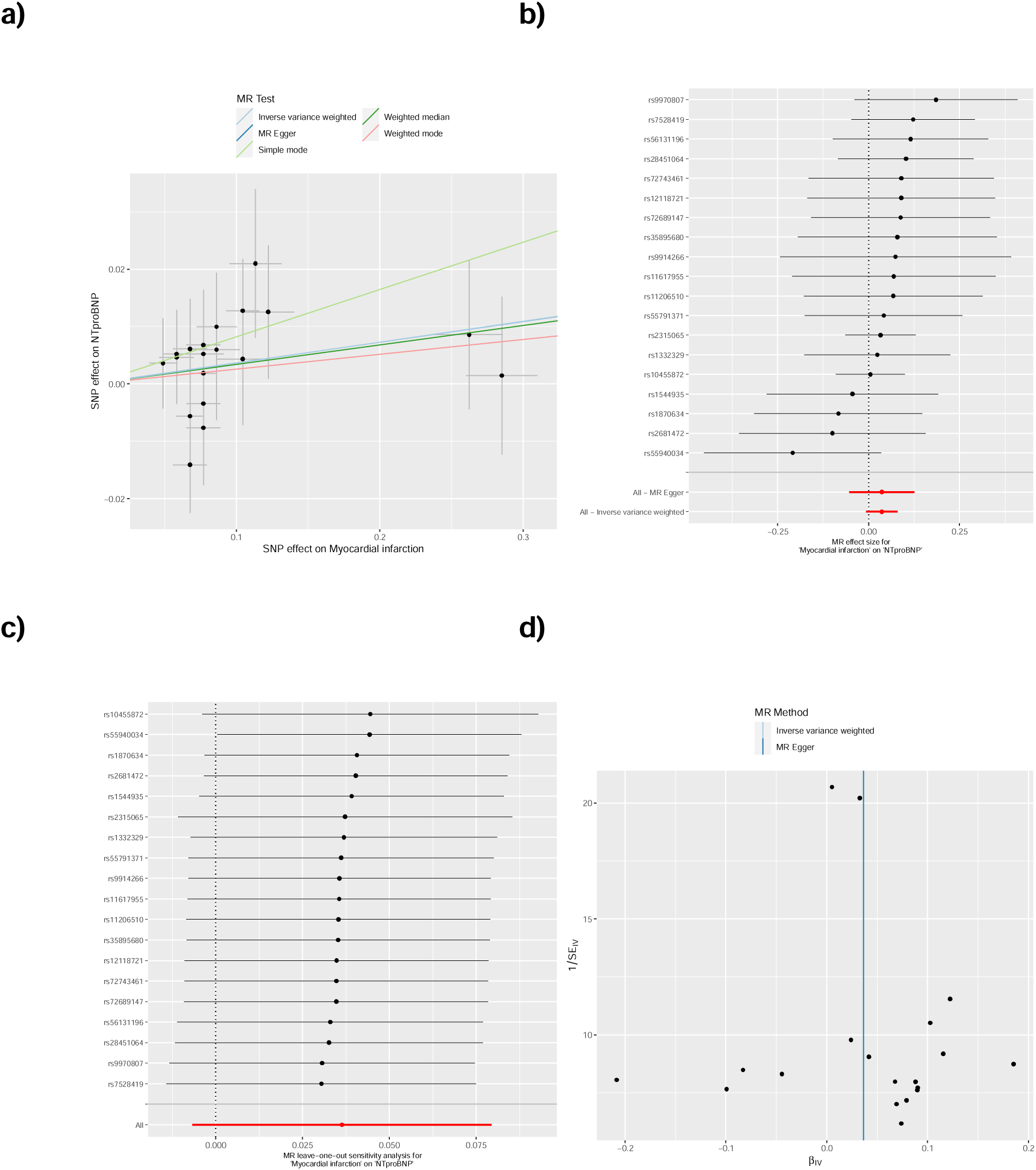
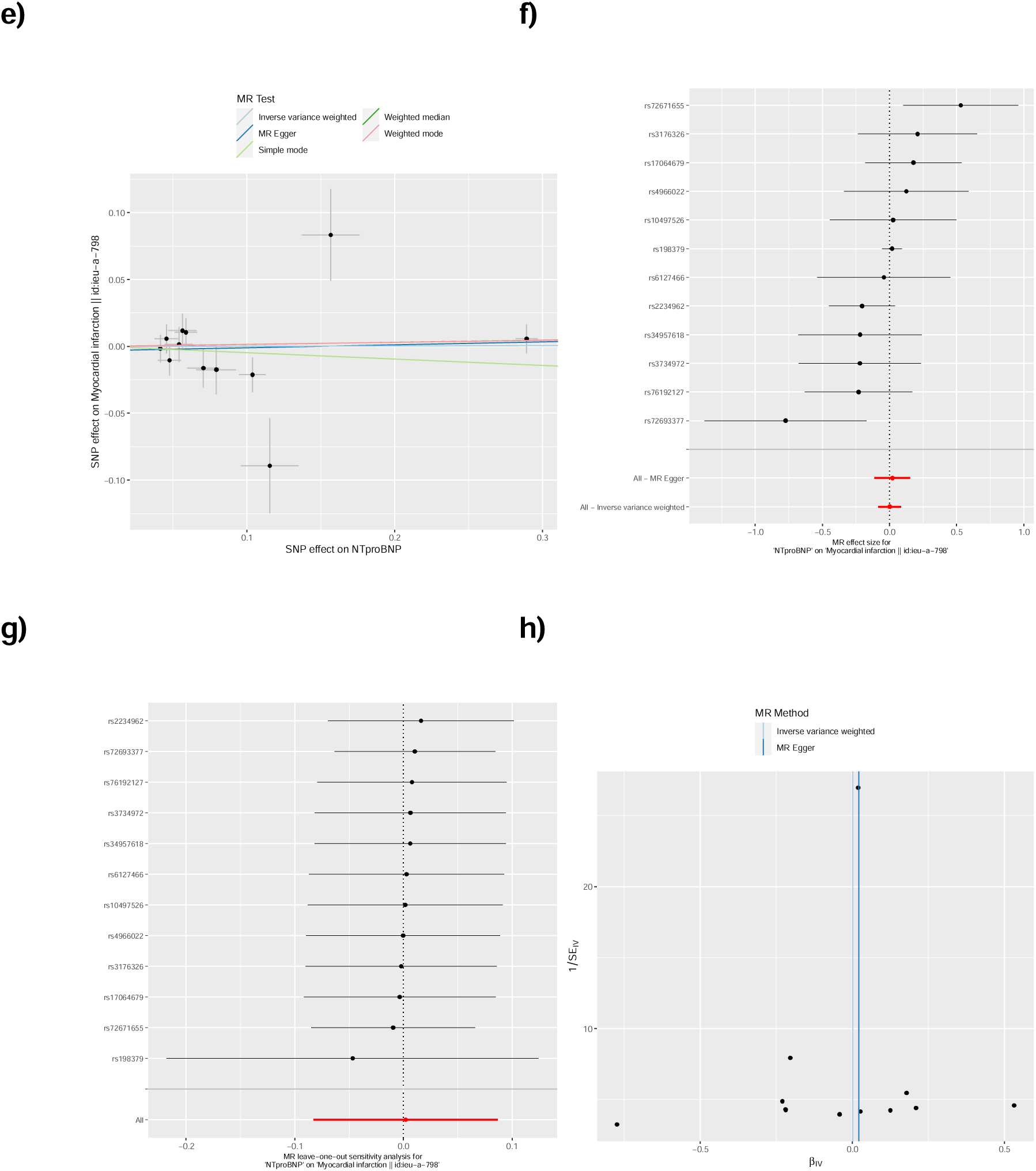
Mendelian randomization analysis of myocardial infarction and NT-proBNP levels. Mendelian randomisation for myocardial infarction (MI) as an exposure **a)-d)** and outcome **e)-h)** for NT-proBNP levels. The plots show summary information on the analyses, performed as per the TwoSampleMR R package. **a)** Mendelian randomization scatter plot for MI as an exposure for NT-proBNP. **b)** Mendelian randomization single SNP funnel plot for MI as an exposure for NT-proBNP. **c)** Mendelian randomization single SNP forest plot for MI as an exposure for NT-proBNP. **c)** Mendelian randomization leave one out plot for MI as an exposure for NT-proBNP. **e)** Mendelian randomization scatter plot for NT-proBNP as an exposure for MI. **f)** Mendelian randomization single SNP funnel plot for NT-proBNP as an exposure for MI. **g)** Mendelian randomization single SNP forest plot for NT-proBNP as an exposure for MI. **h)** Mendelian randomization leave one out plot for NT-proBNP as an exposure for MI.

**Figure S12.**
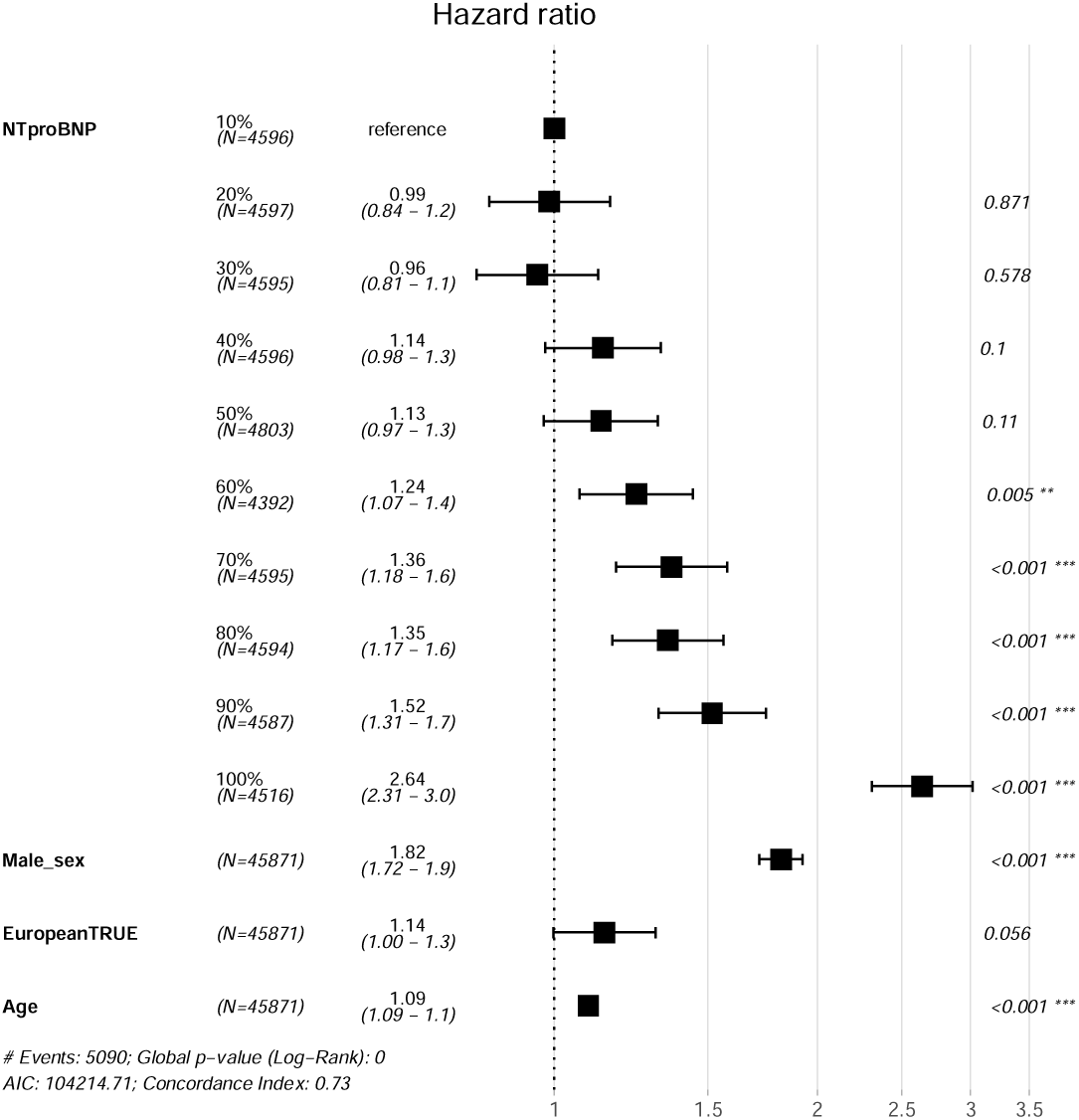
NT-proBNP increases with incident cardiomyopathy. Forest plot of cardiomyopathies by NT-proBNP deciles. The forest plots of Cox proportional hazards regression models were created assessing death or diagnosis from recruitment with those diagnosed before recruitment excluded. Sex (increasing risk is male), European ancestry (increasing risk is European), and age at recruitment, were added to this multivariable analysis.

**Figure S13.**
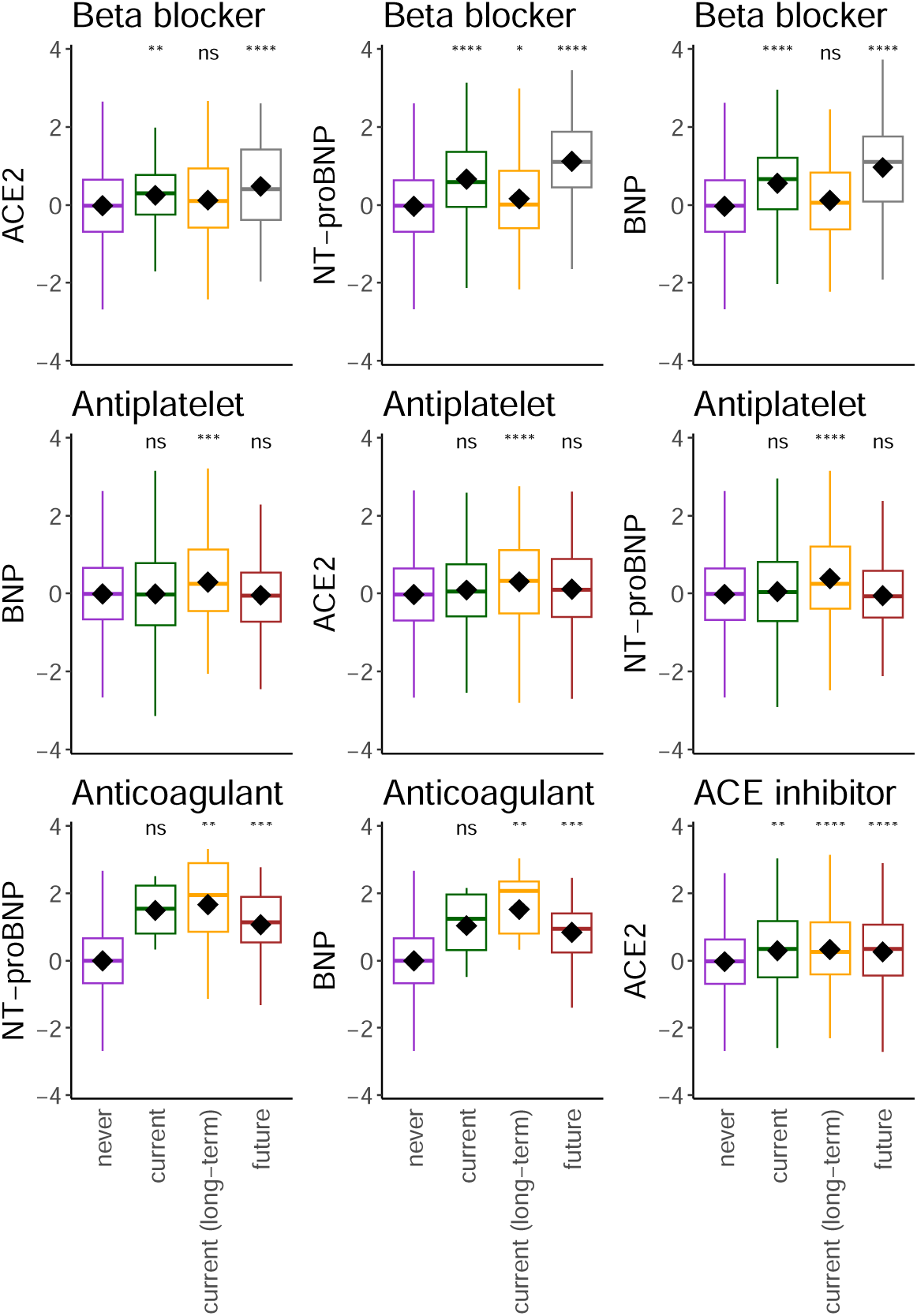
Relationships with medication intake adjusted for time between recruitment and imaging visit. The plots depict the proteins measured at recruitment and adjusted for days to the imaging visit that were significantly increased with medication intake reported only at recruitment (current), or the imaging visit on average 8 years later (future), or reported during both visits (current (long-term)). The significance of differences in means as derived by Student’s t-test are denoted as stars compared to individuals with no report of the medication (never). The data only includes those with proteomics who attended the imaging visit (n=5,324).

**Figure S14.**
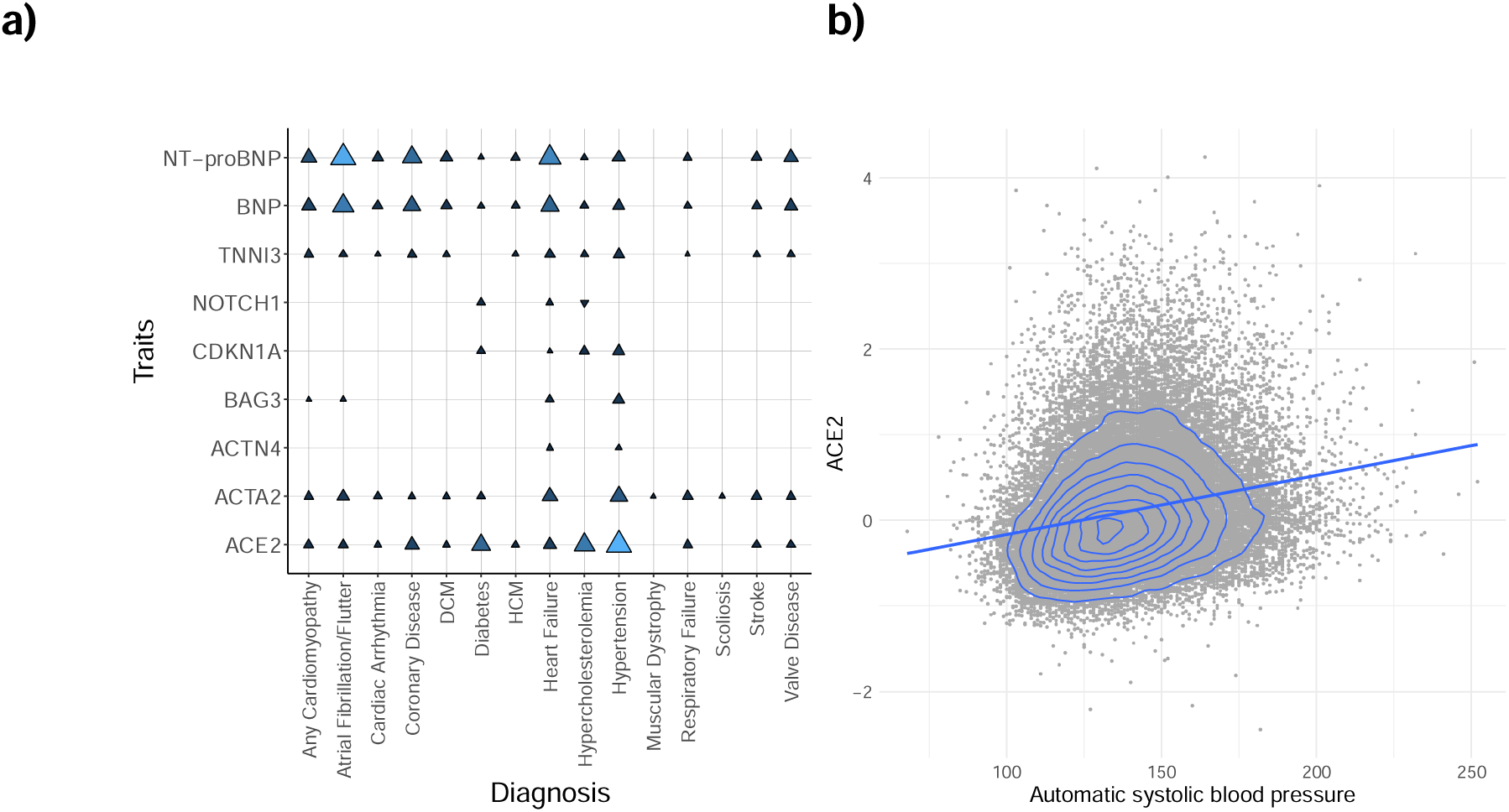
**Curated cardiovascular disease association study and ACE2’s relationship with systolic blood pressure**. **a)** ICD codes as diagnoses are described on the x-axis and the protein traits are on the y-axis. Each point denotes a significant association. The shape and colour denote the direction of effect and odds ratio. The size represents the significance (where larger is more significant). See **Table S2** for the full results. DCM, dilated cardiomyopathy; HCM, hypertrophic cardiomyopathy. **b)** The linear relationship between ACE2 levels and an automatic measure of systolic blood pressure measured at recruitment (R=0.21).

**Figure S15.**
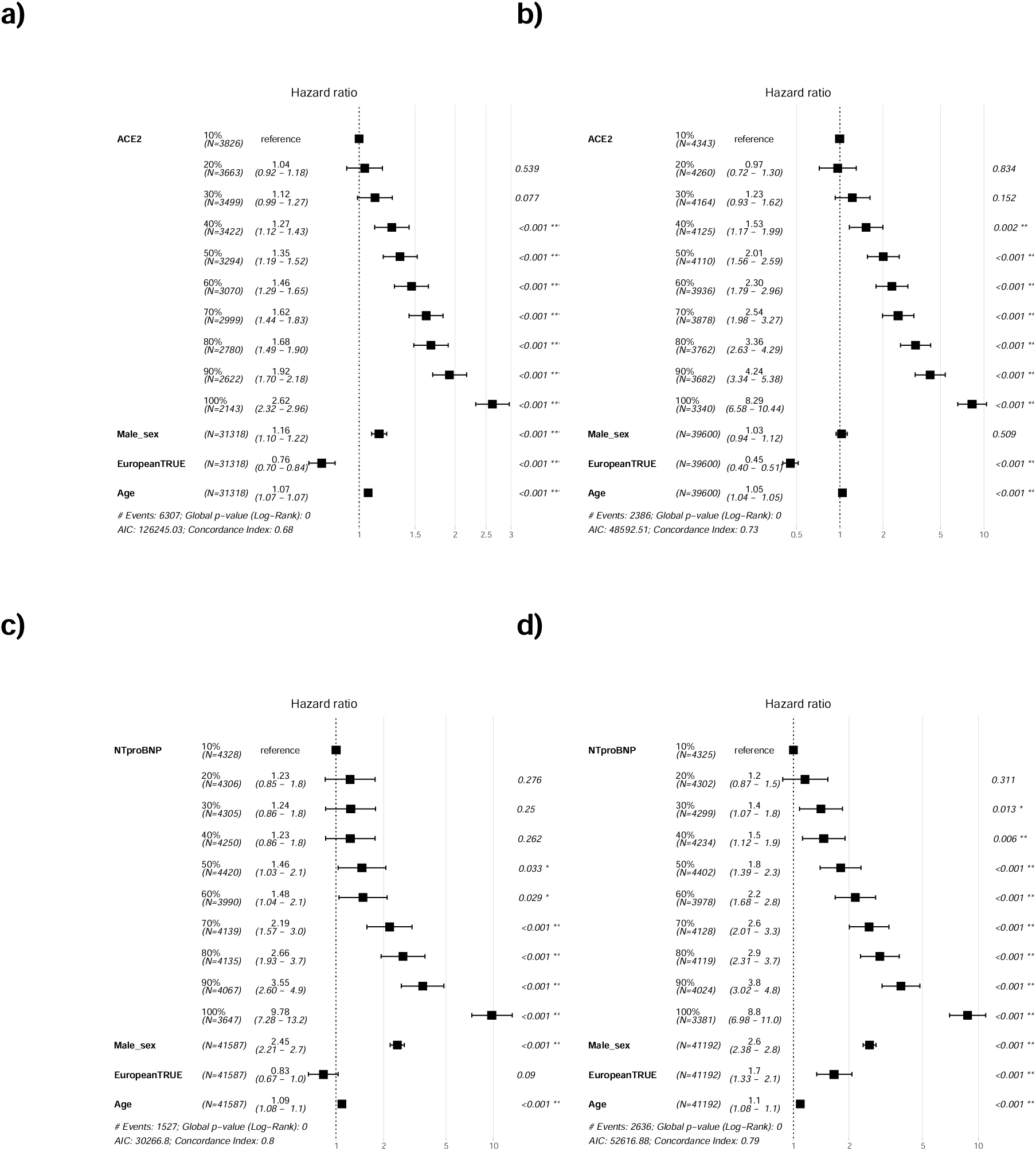
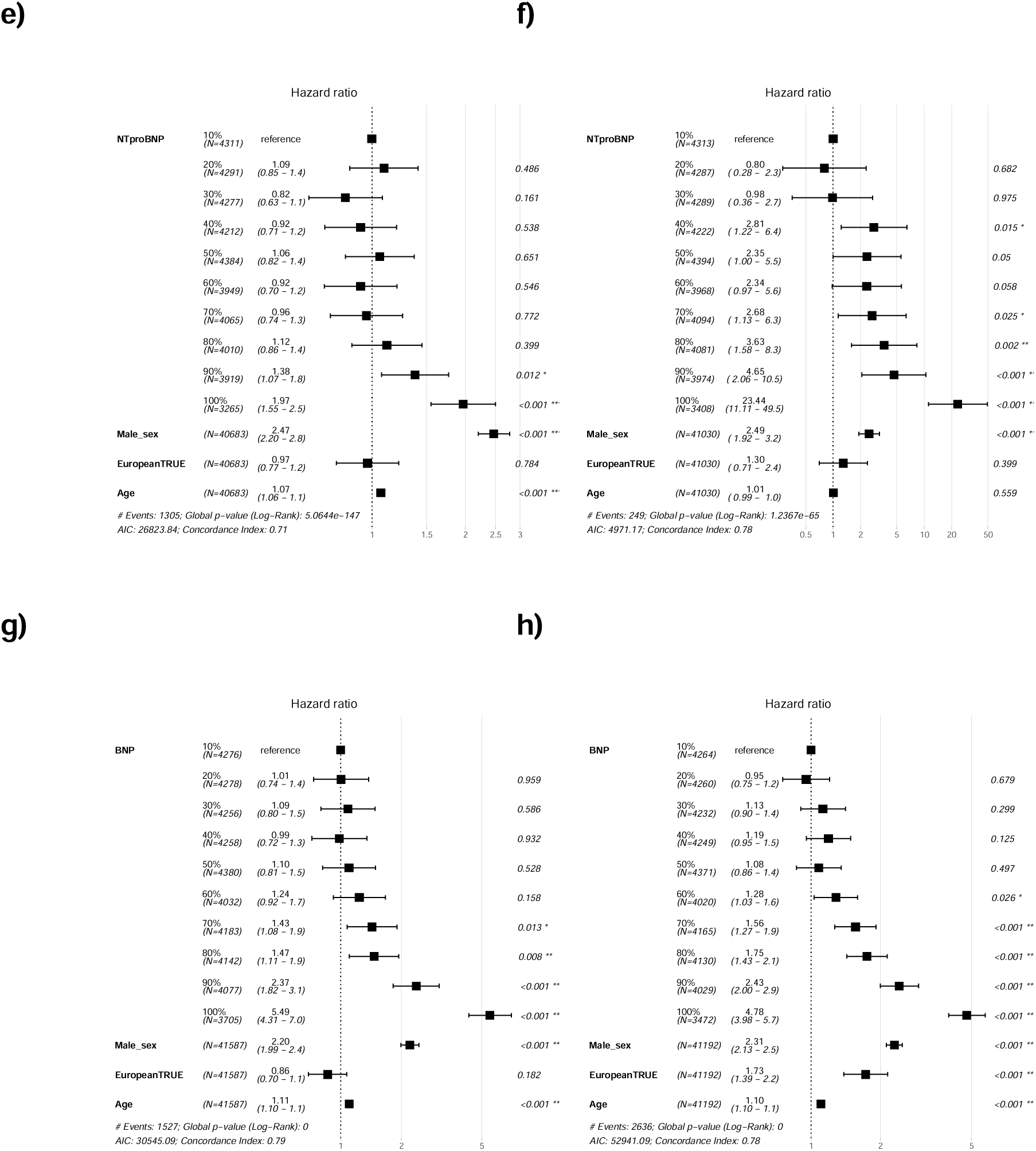
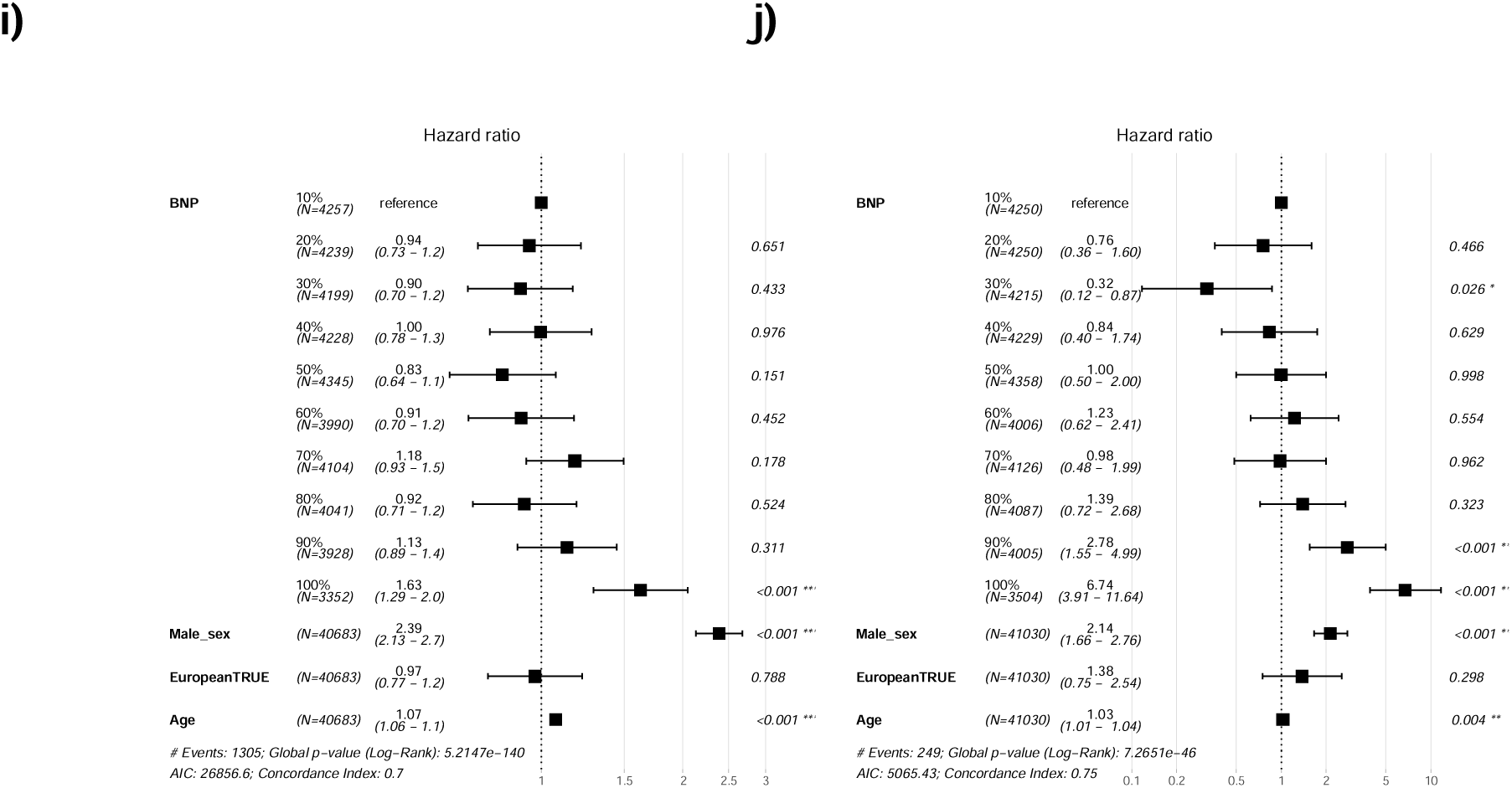
Sensitivity analysis of removing all-cause death from Cox proportional hazards regression models. Deciles of ACE2 levels with incident **a)** hypertension and **a)** diabetes, since recruitment. Deciles of NT-proBNP levels with incident **c)** heart failure, **d)** atrial fibrillation, **e)** myocardial infarction, and **f)** cardiomyopathy, since recruitment. Deciles of BNP levels with incident **g)** heart failure, **h)** atrial fibrillation, **i)** myocardial infarction, and **j)** cardiomyopathy, since recruitment. The forest plots were created assessing diagnosis from recruitment with those diagnosed before recruitment or died without a diagnosis excluded. Sex (increasing risk is male), European ancestry (increasing risk is European), and age at recruitment, were added to this multivariable analysis for comparison.

**Figure S16.**
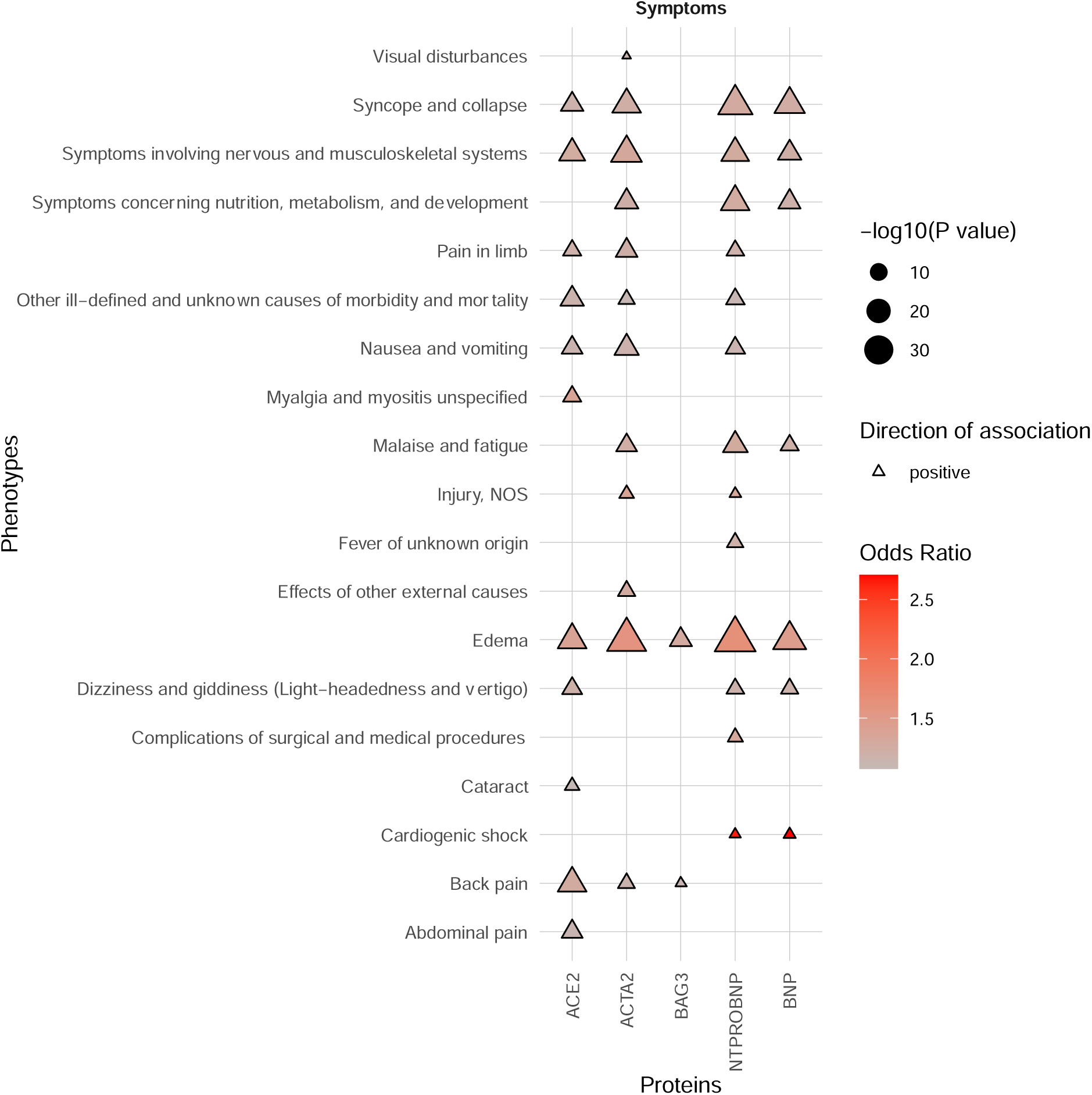
Phenome-wide association study results of the plasma protein levels with the symptoms category of phenotypes. Phenotypes as phecodes are described on the y-axis and the protein traits on the x-axis. Each point denotes a significant PheWAS association with a Bonferroni correction for the number of analyzed phecodes. The shape and colour denote the direction of effect and odds ratio. See **Table S1** for the full PheWAS results.

**Figure S17.**
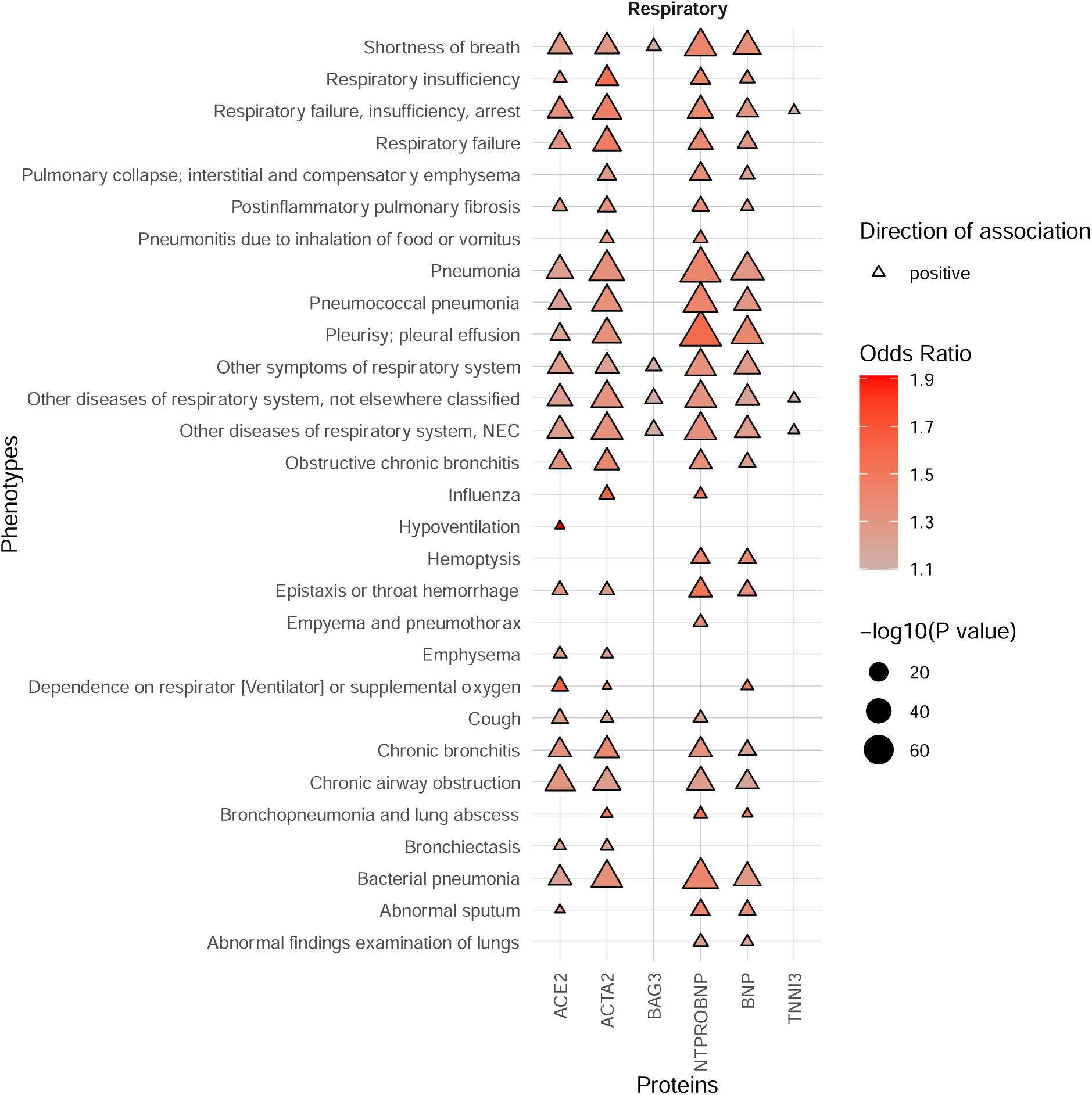
Phenome-wide association study results of the plasma protein levels with the respiratory category of phenotypes. Phenotypes as phecodes are described on the y-axis and the protein traits on the x-axis. Each point denotes a significant PheWAS association with a Bonferroni correction for the number of analyzed phecodes. The shape and colour denote the direction of effect and odds ratio. See **Table S1** for the full PheWAS results.

**Figure S18.**
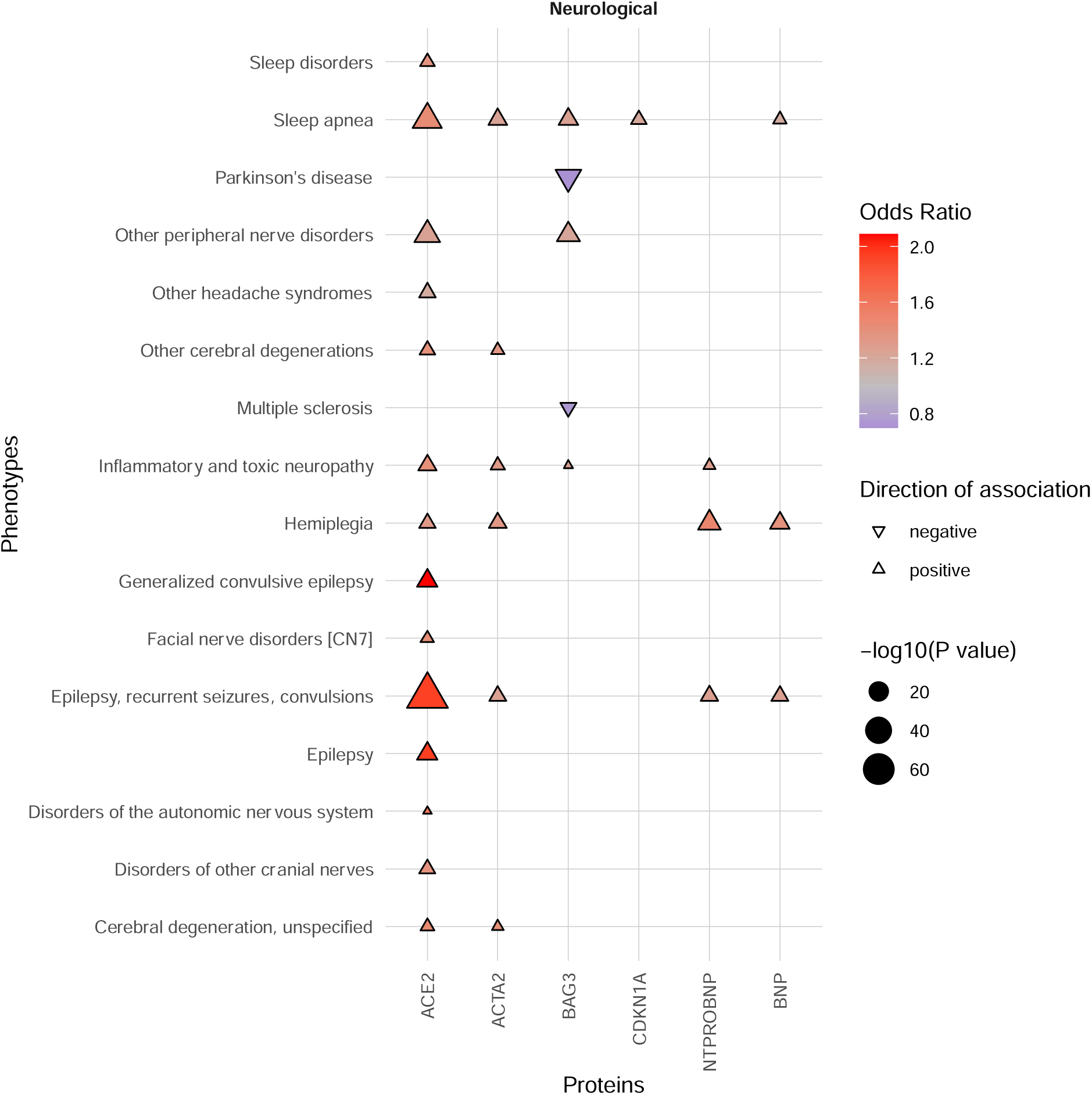
Phenome-wide association study results of the plasma protein levels with the neurological category of phenotypes. Phenotypes as phecodes are described on the y-axis and the protein traits on the x-axis. Each point denotes a significant PheWAS association with a Bonferroni correction for the number of analyzed phecodes. The shape and colour denote the direction of effect and odds ratio. See **Table S1** for the full PheWAS results.

**Figure S19.**
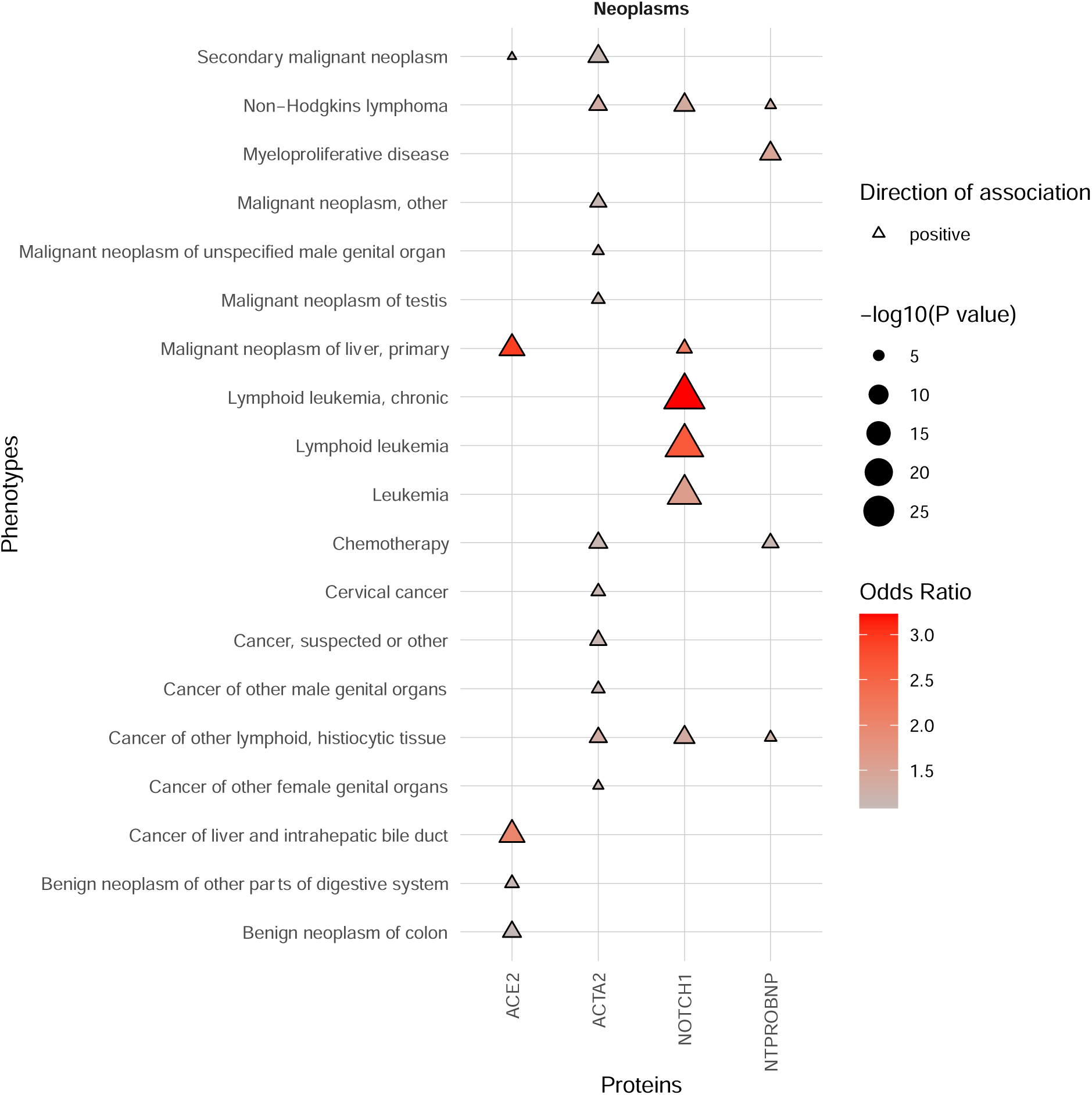
Phenome-wide association study results of the plasma protein levels with the neoplasms category of phenotypes. Phenotypes as phecodes are described on the y-axis and the protein traits on the x-axis. Each point denotes a significant PheWAS association with a Bonferroni correction for the number of analyzed phecodes. The shape and colour denote the direction of effect and odds ratio. See **Table S1** for the full PheWAS results.

**Figure S20.**
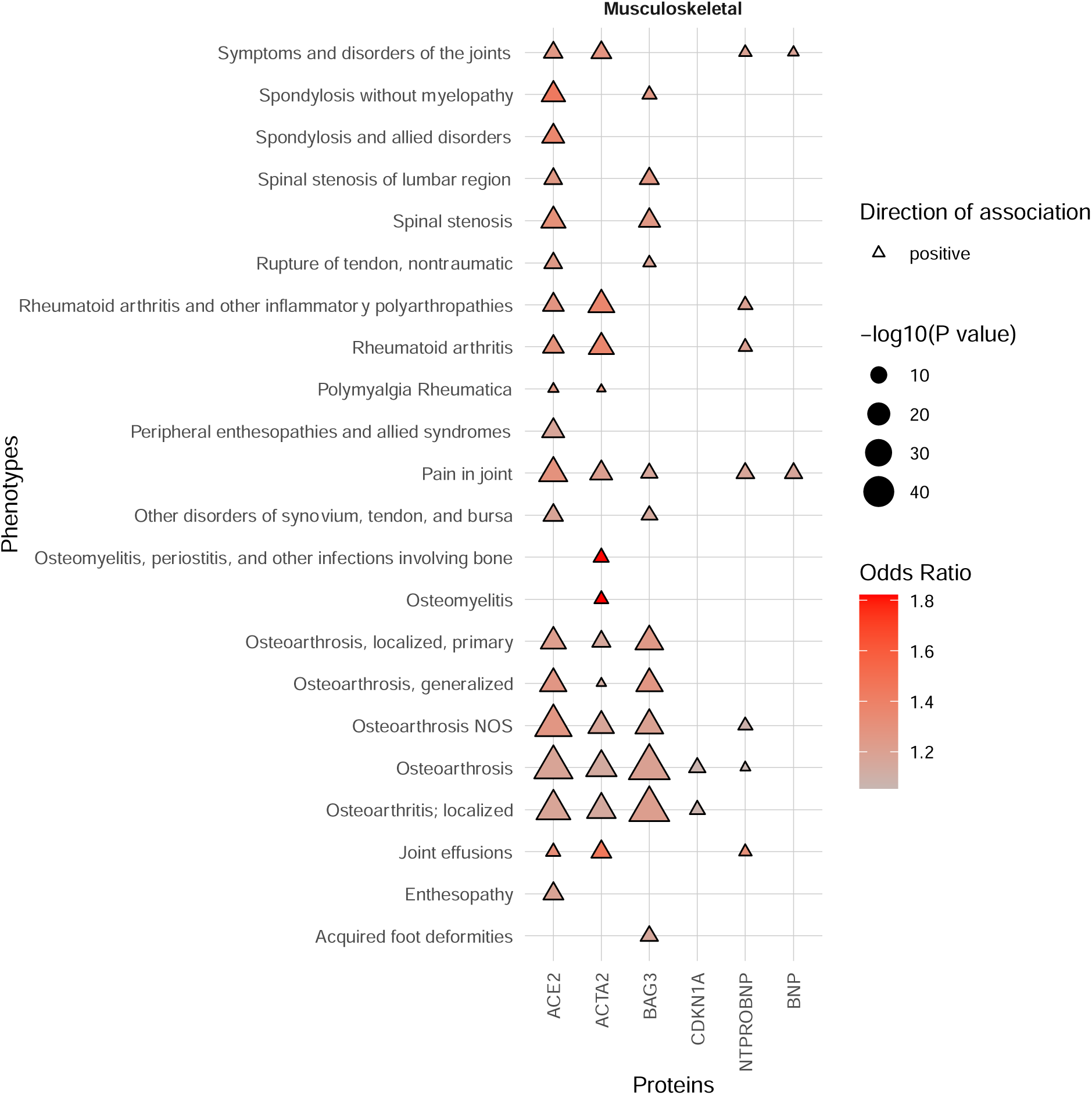
Phenome-wide association study results of the plasma protein levels with the musculoskeletal category of phenotypes. Phenotypes as phecodes are described on the y-axis and the protein traits on the x-axis. Each point denotes a significant PheWAS association with a Bonferroni correction for the number of analyzed phecodes. The shape and colour denote the direction of effect and odds ratio. See **Table S1** for the full PheWAS results.

**Figure S21.**
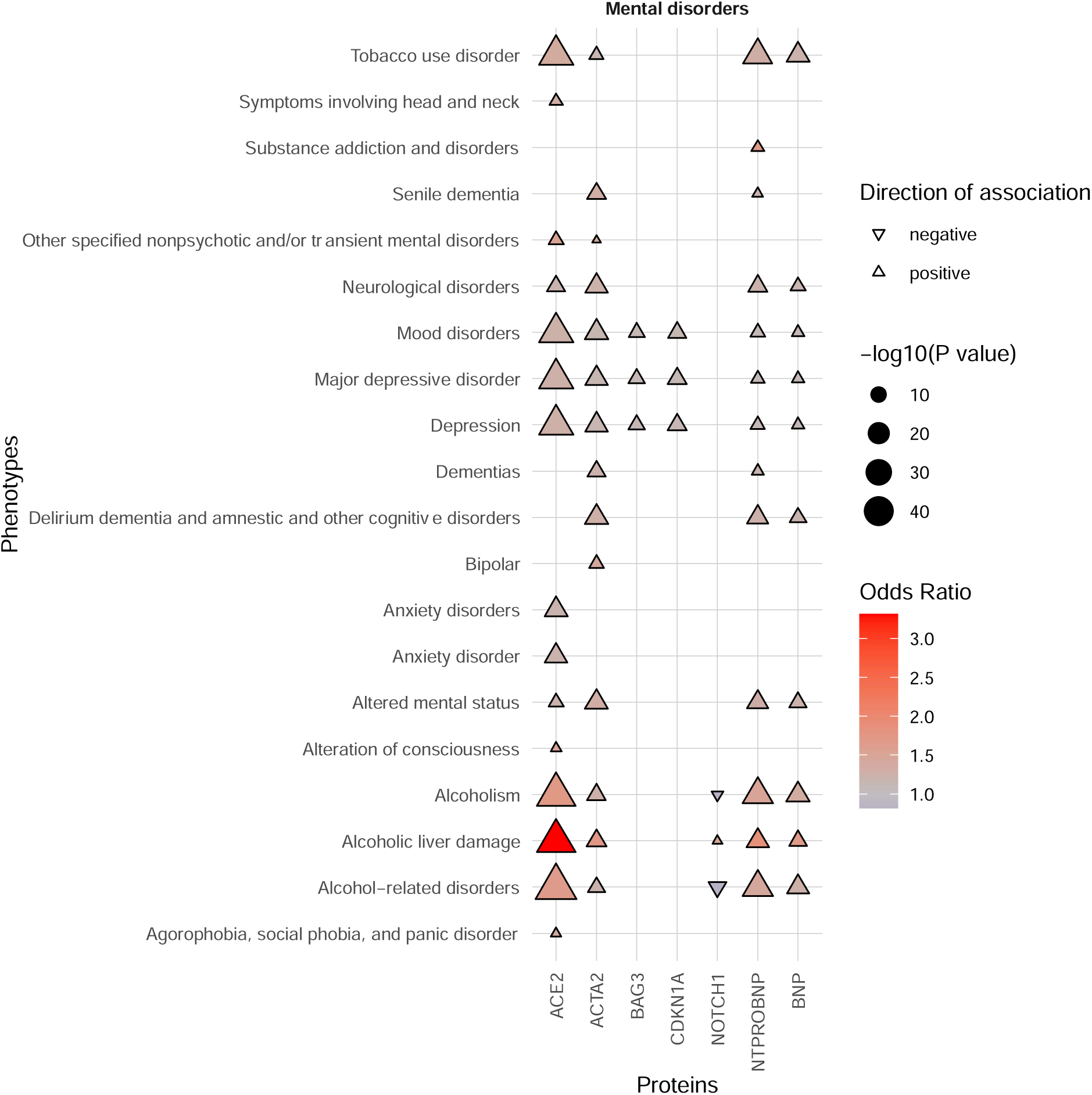
Phenome-wide association study results of the plasma protein levels with the mental disorders category of phenotypes. Phenotypes as phecodes are described on the y-axis and the protein traits on the x-axis. Each point denotes a significant PheWAS association with a Bonferroni correction for the number of analyzed phecodes. The shape and colour denote the direction of effect and odds ratio. See **Table S1** for the full PheWAS results.

**Figure S22.**
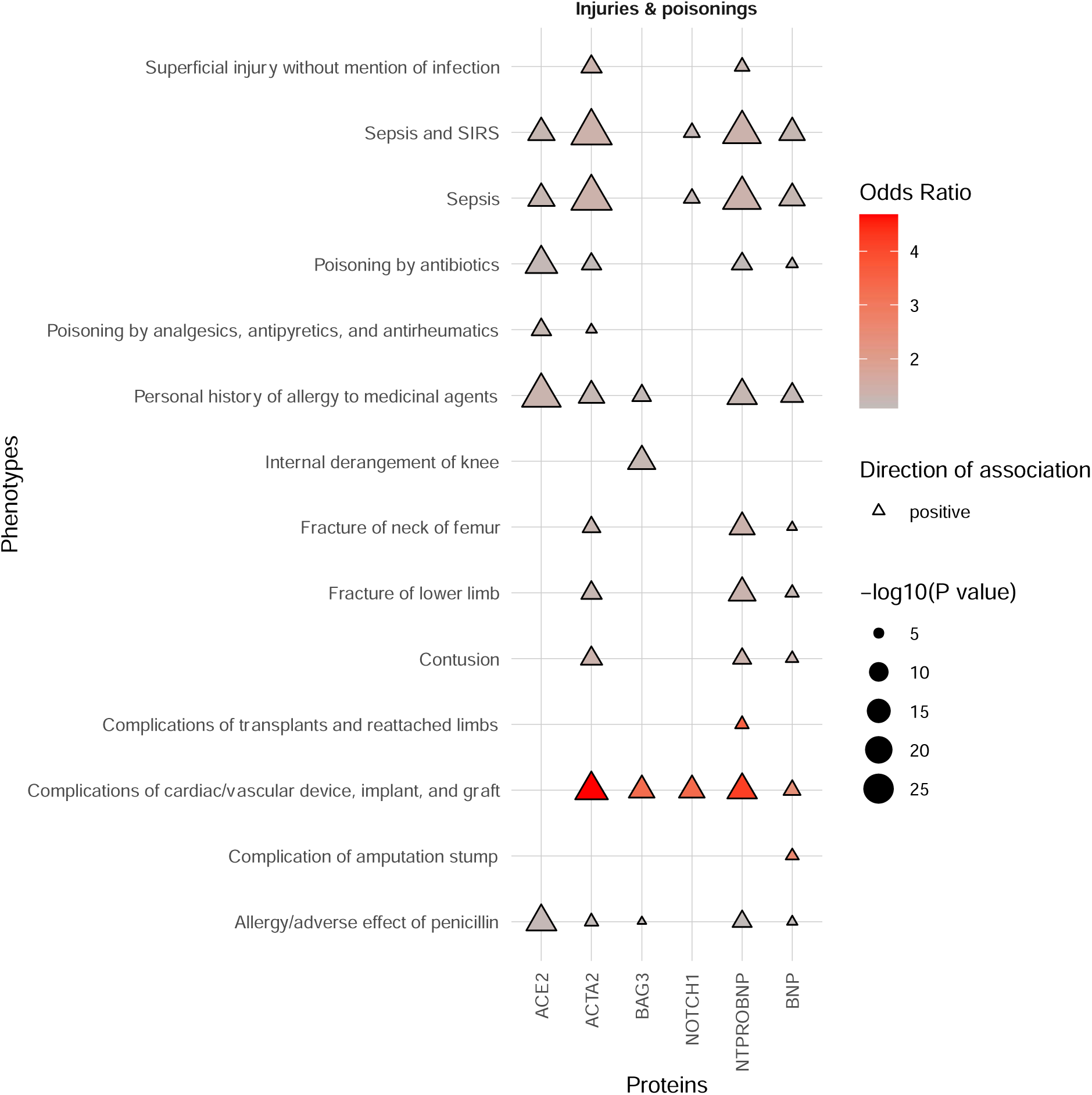
Phenome-wide association study results of the plasma protein levels with the injuries and poisonings category of phenotypes. Phenotypes as phecodes are described on the y-axis and the protein traits on the x-axis. Each point denotes a significant PheWAS association with a Bonferroni correction for the number of analyzed phecodes. The shape and colour denote the direction of effect and odds ratio. See **Table S1** for the full PheWAS results.

**Figure S23.**
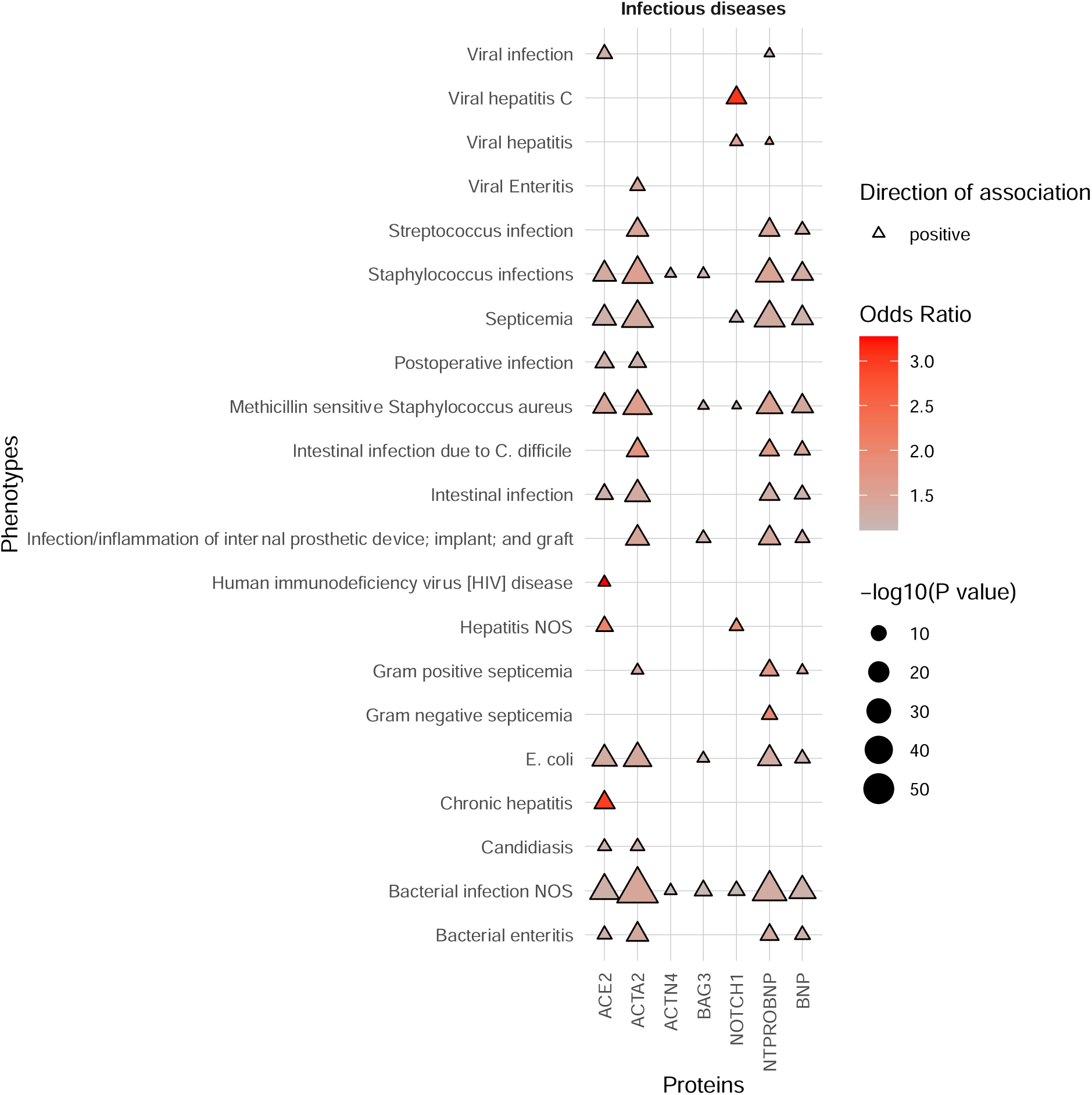
Phenome-wide association study results of the plasma protein levels with the infectious diseases category of phenotypes. Phenotypes as phecodes are described on the y-axis and the protein traits on the x-axis. Each point denotes a significant PheWAS association with a Bonferroni correction for the number of analyzed phecodes. The shape and colour denote the direction of effect and odds ratio. See **Table S1** for the full PheWAS results.

**Figure S24.**
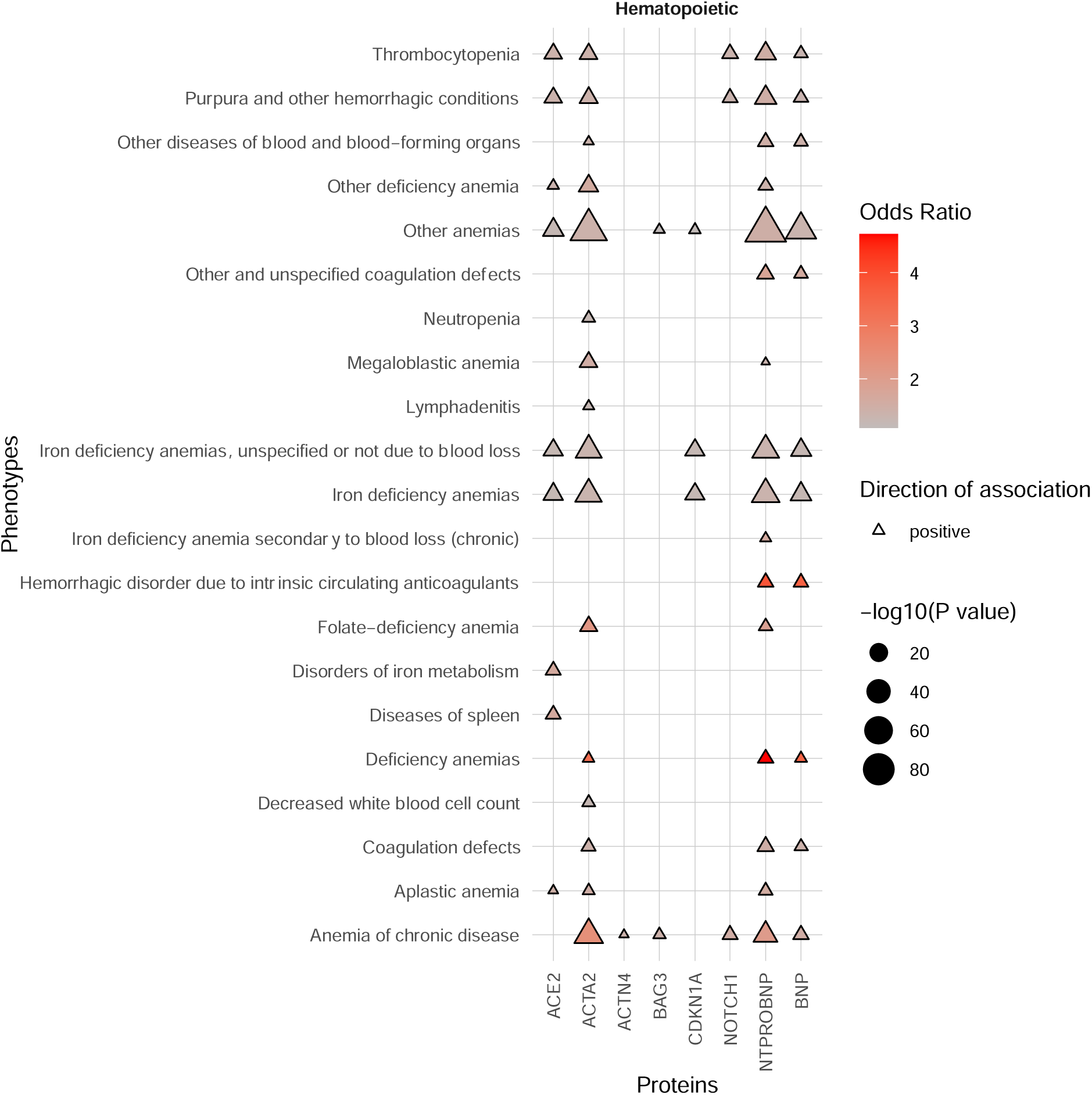
Phenome-wide association study results of the plasma protein levels with the hematopoietic category of phenotypes. Phenotypes as phecodes are described on the y-axis and the protein traits on the x-axis. Each point denotes a significant PheWAS association with a Bonferroni correction for the number of analyzed phecodes. The shape and colour denote the direction of effect and odds ratio. See **Table S1** for the full PheWAS results.

**Figure S25.**
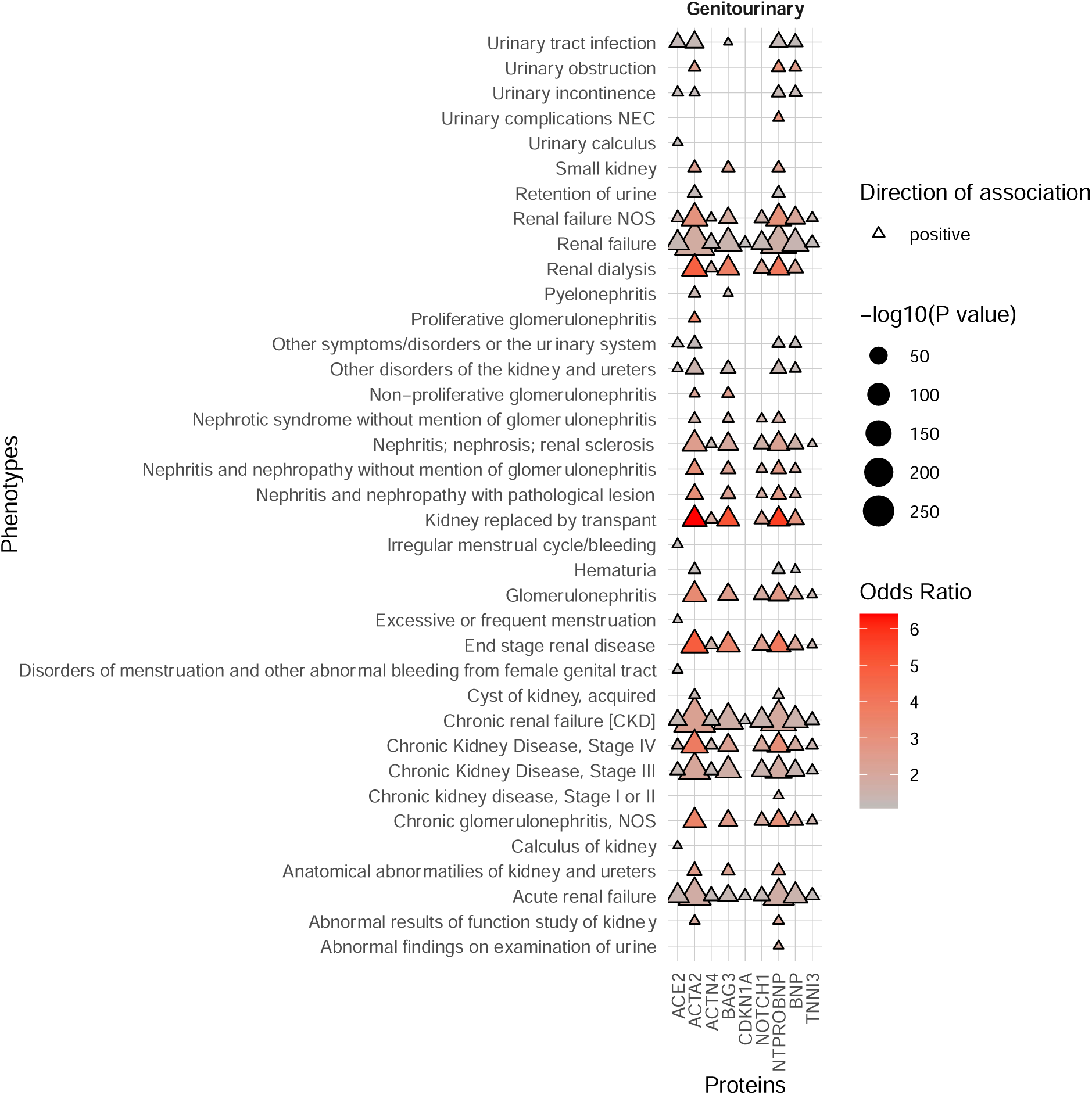
Phenome-wide association study results of the plasma protein levels with the genitourinary category of phenotypes. Phenotypes as phecodes are described on the y-axis and the protein traits on the x-axis. Each point denotes a significant PheWAS association with a Bonferroni correction for the number of analyzed phecodes. The shape and colour denote the direction of effect and odds ratio. See **Table S1** for the full PheWAS results.

**Figure S26.**
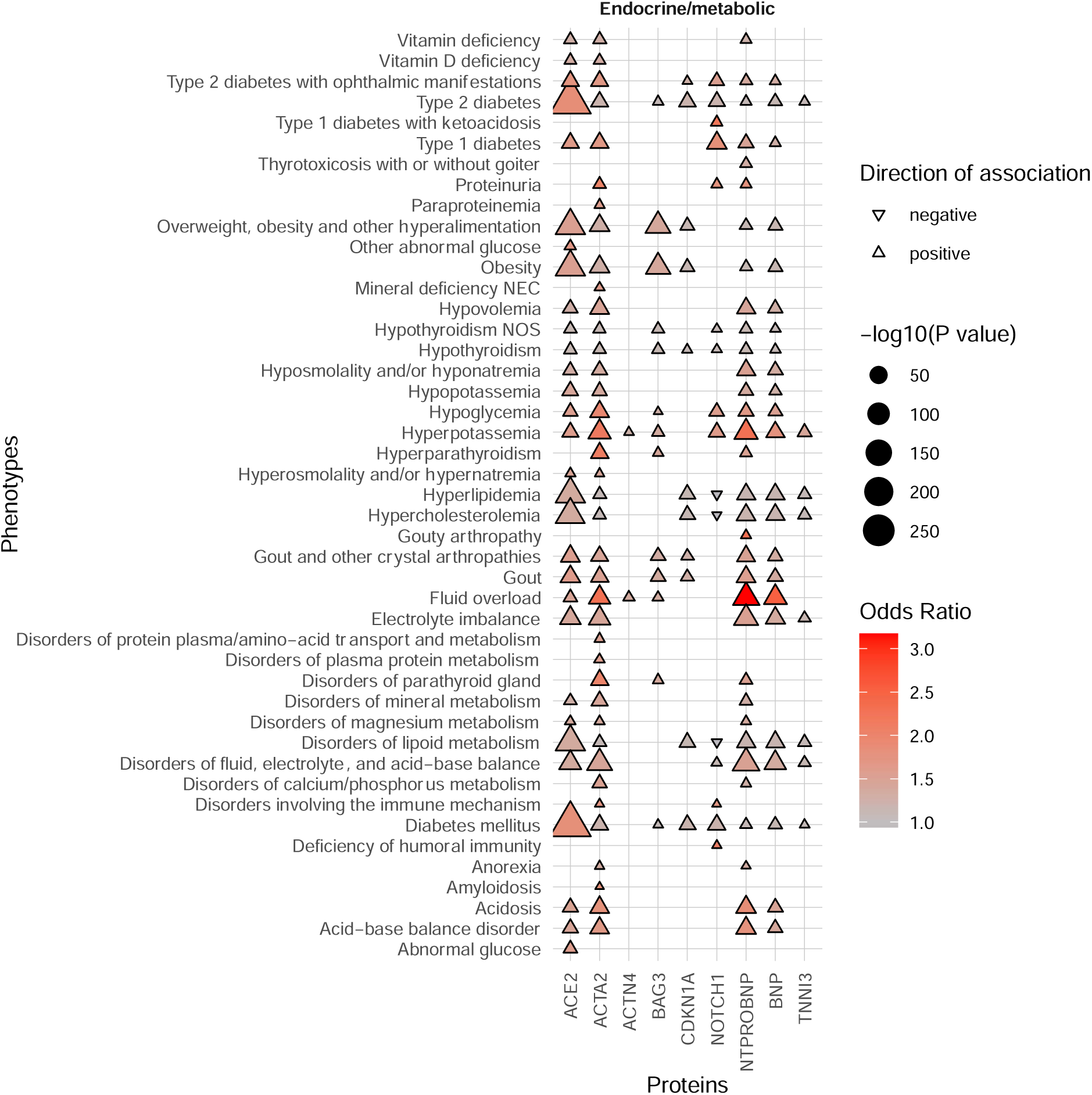
Phenome-wide association study results of the plasma protein levels with the endocrine and metabolic category of phenotypes. Phenotypes as phecodes are described on the y-axis and the protein traits on the x-axis. Each point denotes a significant PheWAS association with a Bonferroni correction for the number of analyzed phecodes. The shape and colour denote the direction of effect and odds ratio. See **Table S1** for the full PheWAS results.

**Figure S27.**
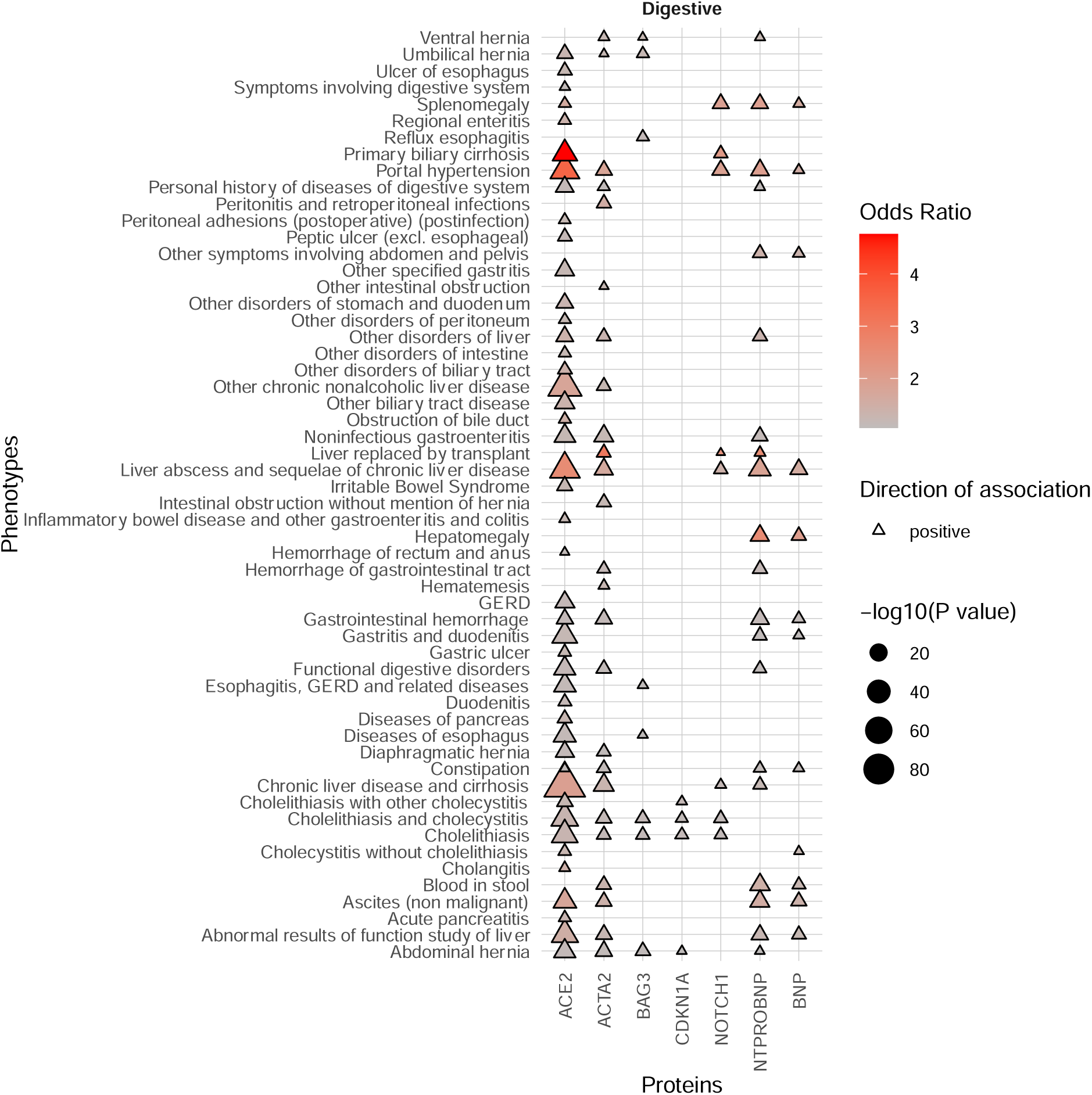
Phenome-wide association study results of the plasma protein levels with the digestive category of phenotypes. Phenotypes as phecodes are described on the y-axis and the protein traits on the x-axis. Each point denotes a significant PheWAS association with a Bonferroni correction for the number of analyzed phecodes. The shape and colour denote the direction of effect and odds ratio. See **Table S1** for the full PheWAS results.

**Figure S28.**
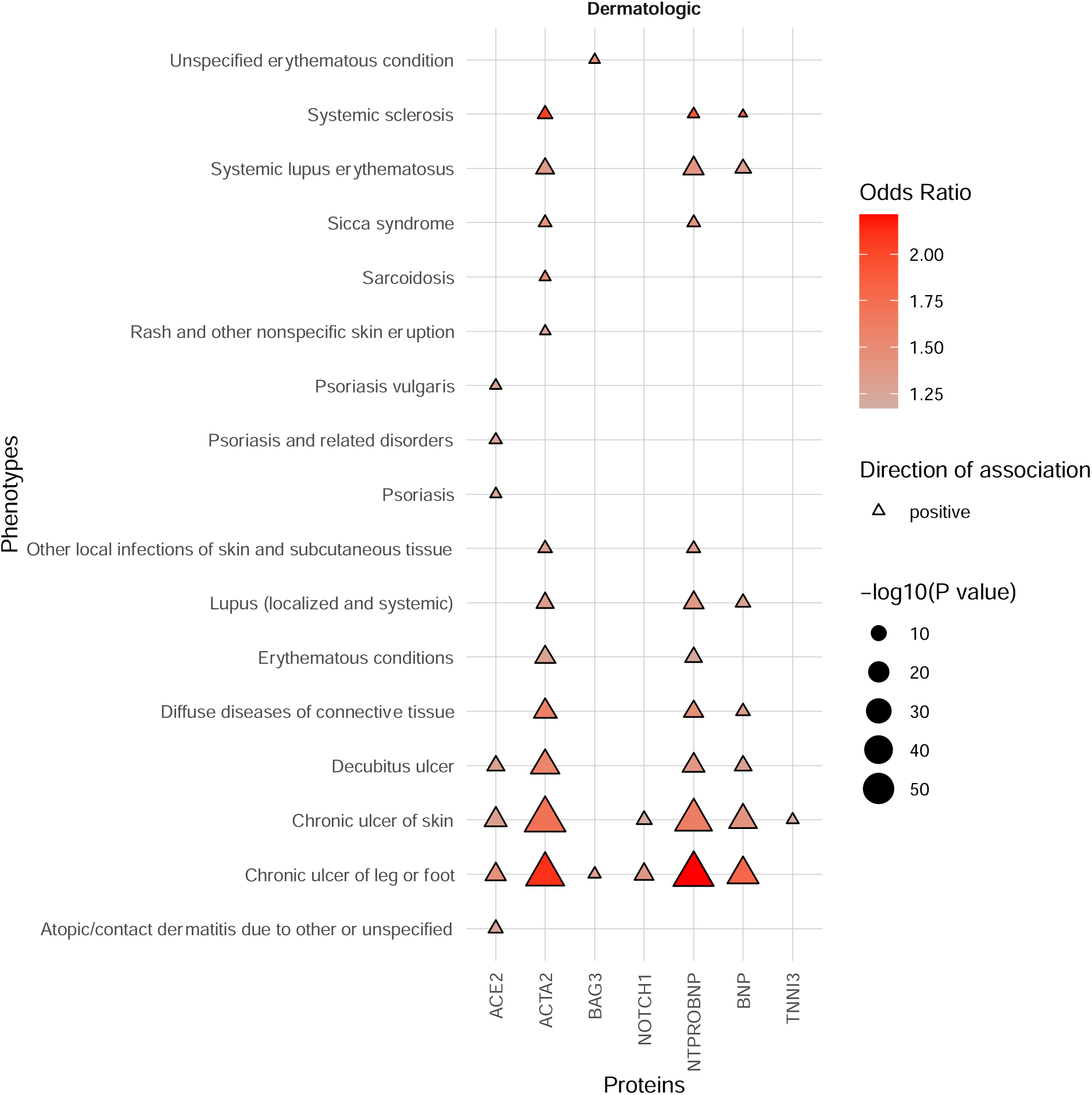
Phenome-wide association study results of the plasma protein levels with the dermatologic category of phenotypes. Phenotypes as phecodes are described on the y-axis and the protein traits on the x-axis. Each point denotes a significant PheWAS association with a Bonferroni correction for the number of analyzed phecodes. The shape and colour denote the direction of effect and odds ratio. See **Table S1** for the full PheWAS results.

**Figure S29.**
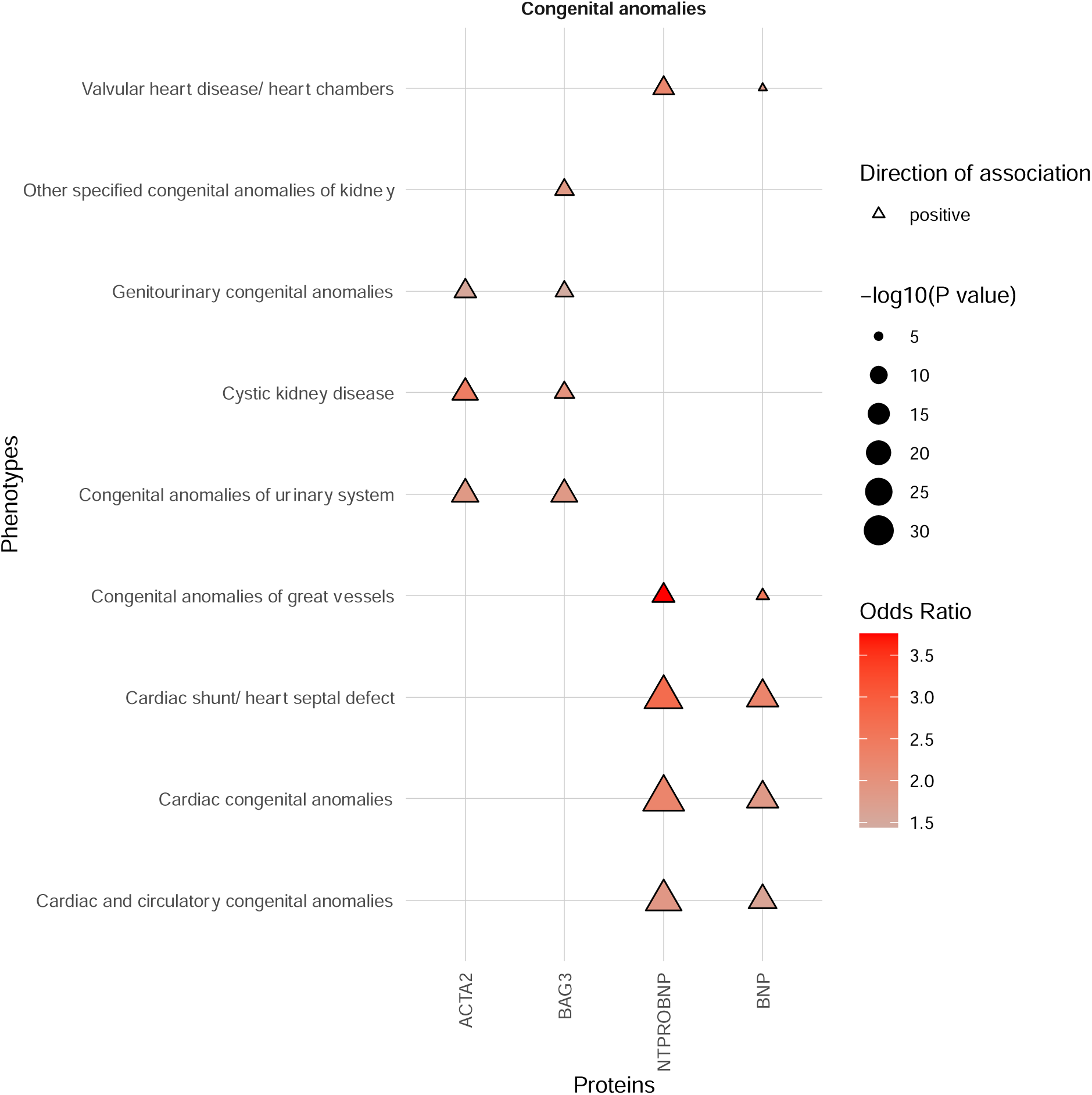
Phenome-wide association study results of the plasma protein levels with the congenital anomalies category of phenotypes. Phenotypes as phecodes are described on the y-axis and the protein traits on the x-axis. Each point denotes a significant PheWAS association with a Bonferroni correction for the number of analyzed phecodes. The shape and colour denote the direction of effect and odds ratio. See **Table S1** for the full PheWAS results.

**Figure S30.**
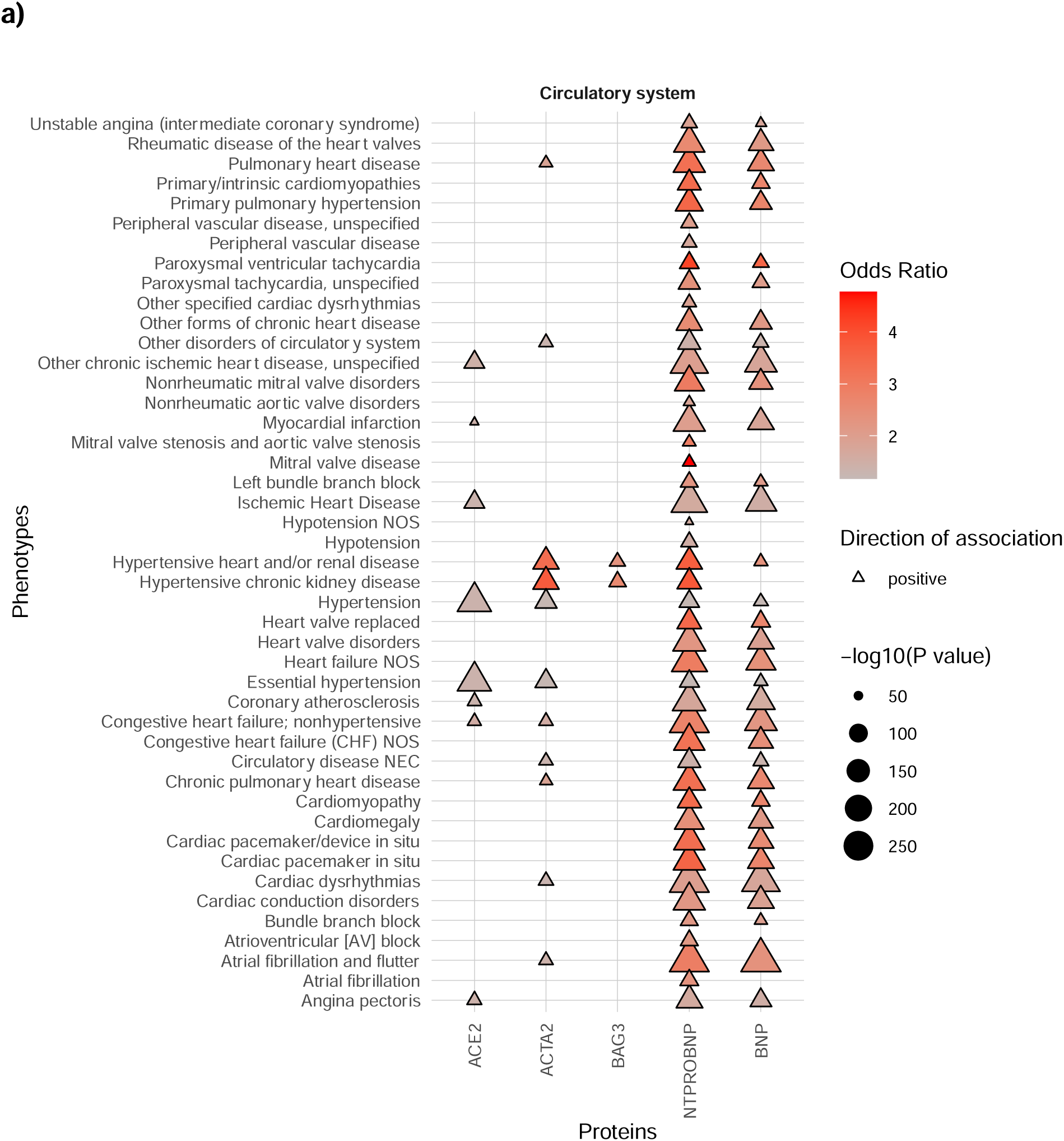
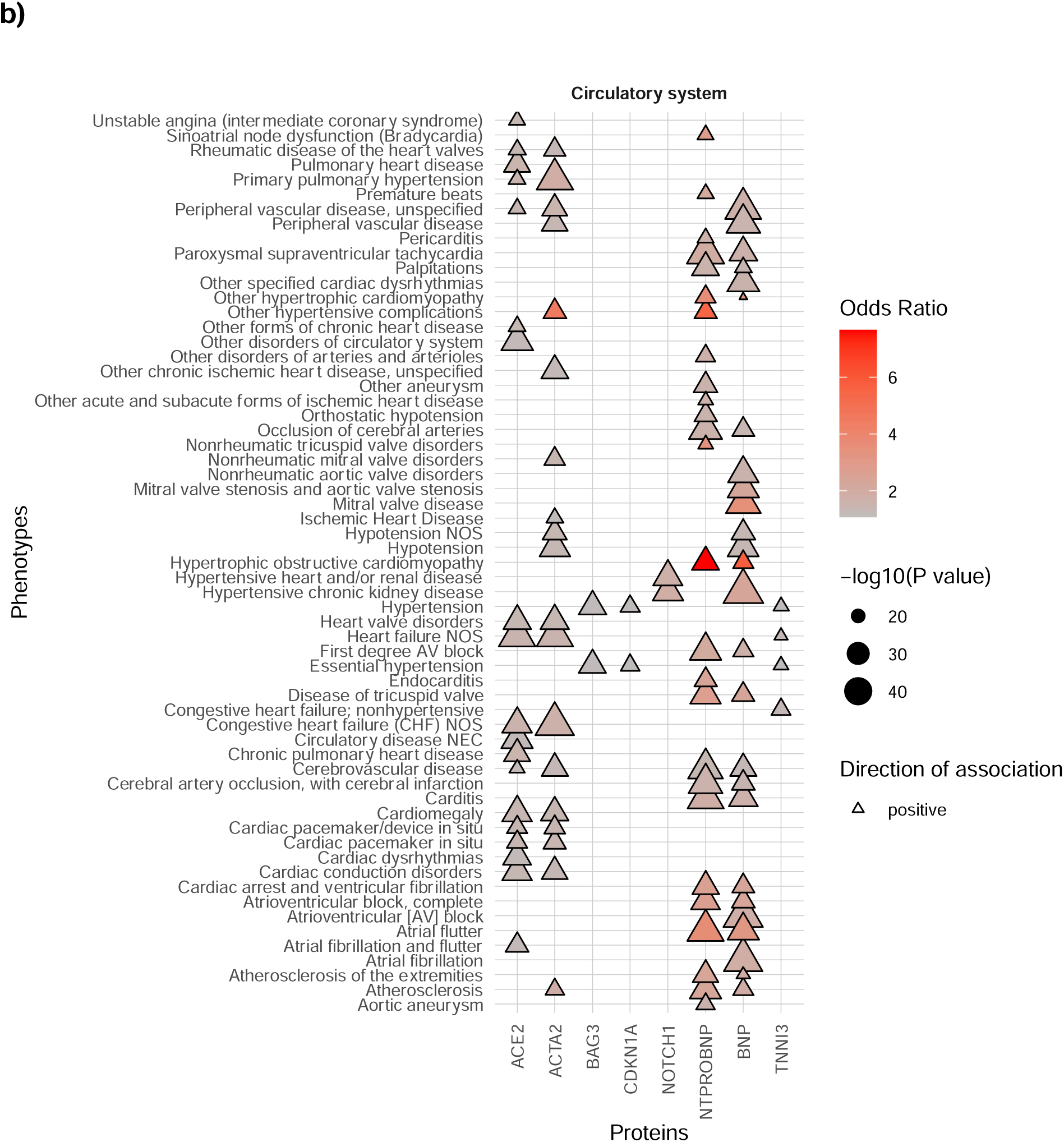
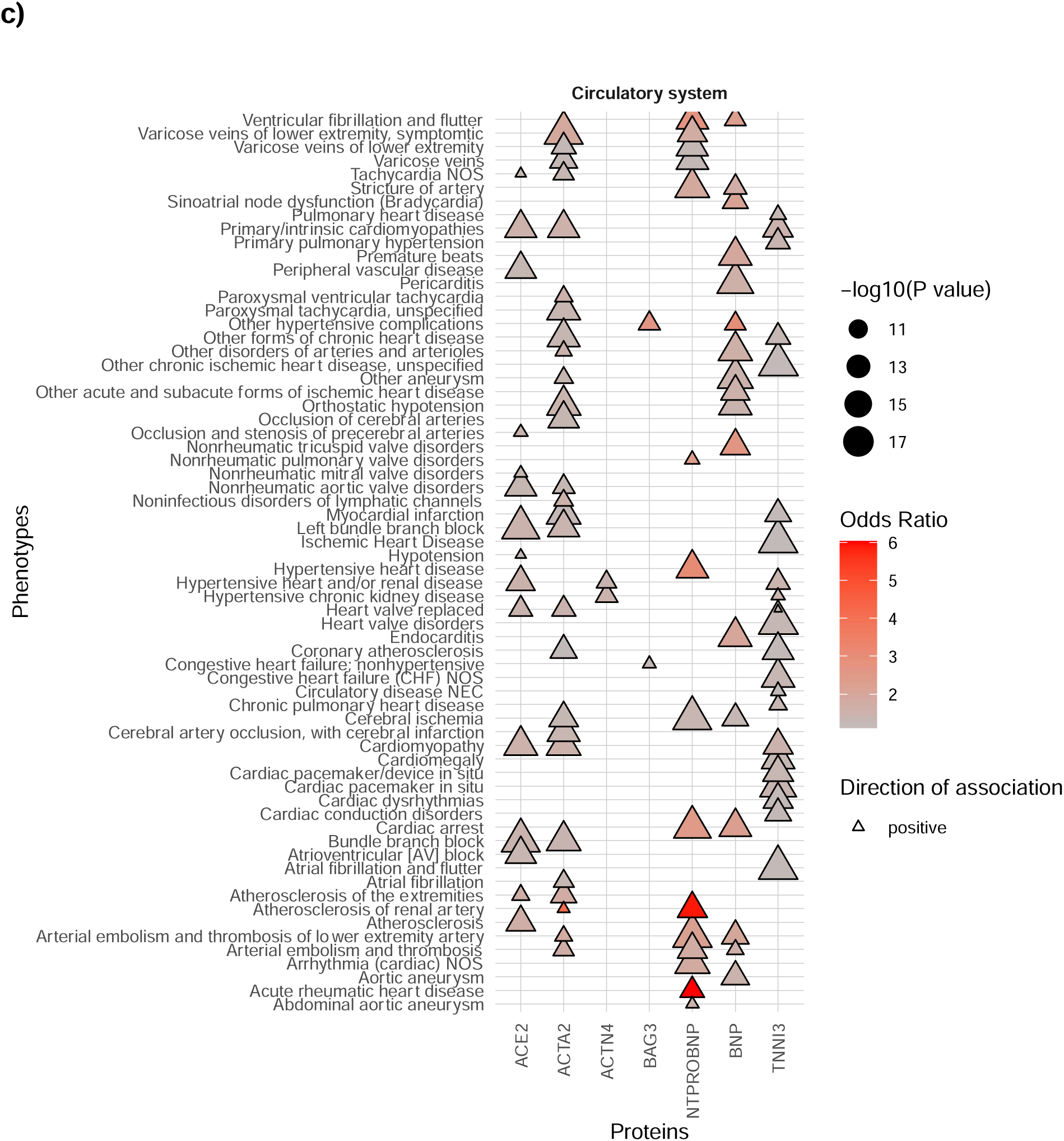
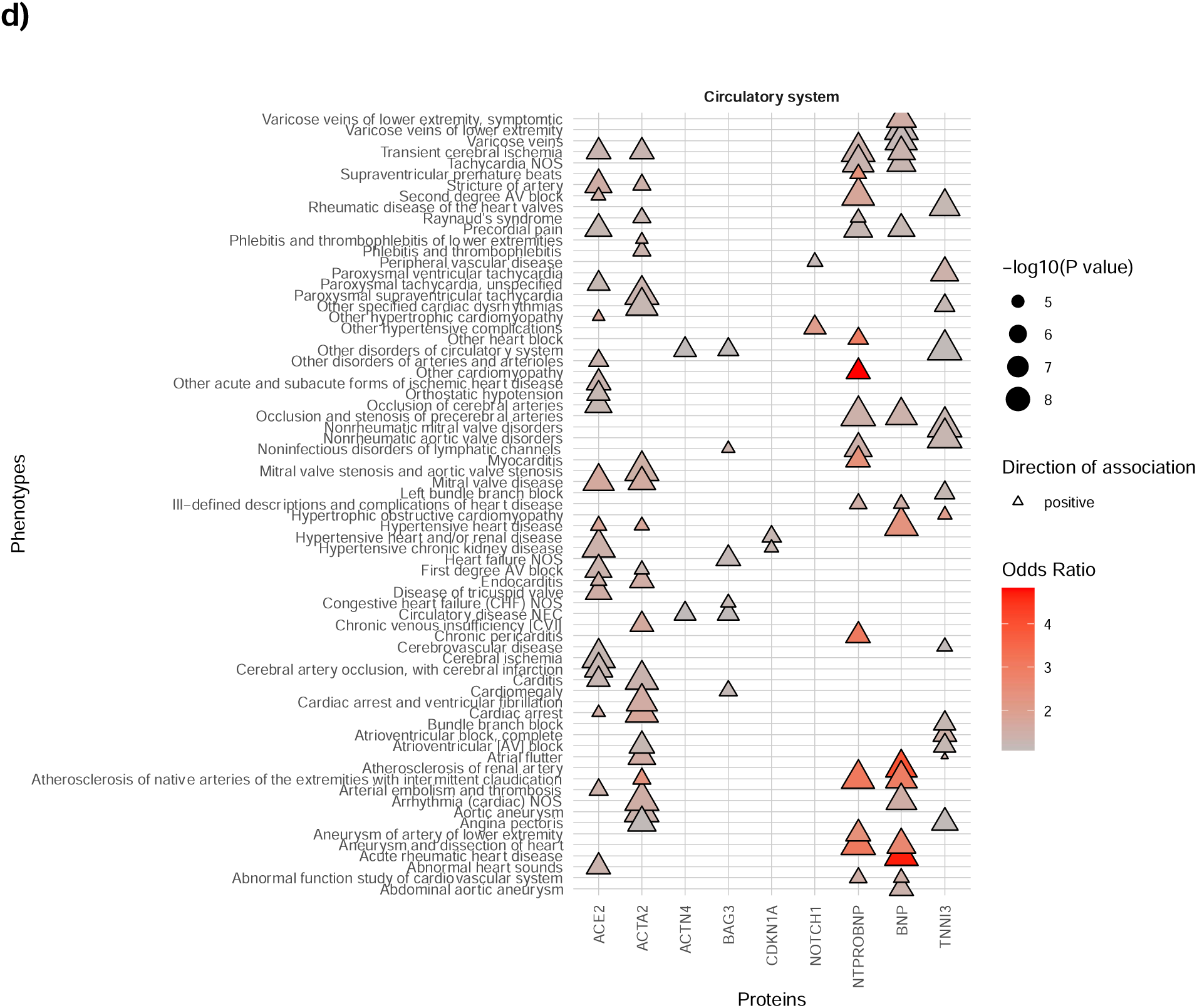
Phenome-wide association study results of the plasma protein levels with the circulatory system category of phenotypes. **a)-d)** results are separated into four plots for clarity. Phenotypes as phecodes are described on the y-axis and the protein traits on the x-axis. Each point denotes a significant PheWAS association with a Bonferroni correction for the number of analyzed phecodes. The shape and colour denote the direction of effect and odds ratio. See **Table S1** for the full PheWAS results.

